# Detecting and Adjusting for Hidden Biases due to Phenotype Misclassification in Genome-Wide Association Studies

**DOI:** 10.1101/2023.01.17.23284670

**Authors:** David Burstein, Gabriel E. Hoffman, Sonali Gupta, Suzannah De Almeida, Deepika Mathur, Sanan Venkatesh, Karen Therrien, Ayman H. Fanous, Tim B. Bigdeli, Philip D. Harvey, Panos Roussos, Georgios Voloudakis

## Abstract

With the advent of healthcare-based genotyped biobanks, genome-wide association studies (GWAS) leverage larger sample sizes, incorporate patients with diverse ancestries and introduce noisier phenotypic definitions. Yet the extent and impact of effect size dilution, from phenotypic misclassification or environmental confounders, on large-scale datasets is not currently well understood due to a lack of statistical methods to estimate relevant parameters from empirical data. Here, we develop a statistical method and scalable software, PheMED, <u>Phe</u>notypic <u>M</u>easurement of <u>E</u>ffective <u>D</u>ilution, that leverages GWAS summary statistics to quantify genome-wide effect size dilution across different phenotypic definitions and cohorts . We formulate conditions illustrating how the parameters estimated by PheMED relate to the negative and positive predictive value of the labeled phenotype, compared to ground truth. We apply our methodology to detect multiple instances of statistically significant dilution in real-world data. We then present multiple downstream applications, where dilution, irrespective of the exact cause, biases downstream GWAS replication and heritability analyses despite utilizing current best practices, and provide a dilution-aware meta-analysis approach that outperforms existing methods. Consequently, we anticipate that PheMED will be a valuable tool for researchers to flag potential data quality issues and harmonize differences in effect size distributions across GWAS.

## Introduction

Interpreting and integrating the results from different genome-wide association (GWA) studies can be challenging, as we anticipate different degrees of phenotypic data quality across studies and populations. In particular, phenotypic misclassification can significantly bias the effect size estimates from GWA studies, as the mislabeled phenotype inflates the similarity between the cases and controls, resulting in diluted effect sizes^1^. While we anticipate phenotypic misclassification when leveraging noisier phenotypic definitions from large population- and healthcare-based biobanks, these dilution-driven systematic biases can arise even when two phenotypes appear to share identical definitions. For example, in the context of healthcare disparities, prior literature has emphasized that African American patients face higher rates of misdiagnosis than European Americans^2,3^, which would lead to diluted effect sizes in African ancestry GWA studies, as genetically-inferred ancestry is correlated with the social construct of race. Consequently, researchers would greatly benefit from tools to estimate the level of phenotypic dilution across different cohorts, as including poor quality phenotypes can corrupt downstream replication and validation analyses.

While methodologies that address phenotypic dilution do exist, the utility of these tools is limited due to various methodological constraints. For example, they typically leverage individual level data^4–9^, possibly with knowledge of a gold standard phenotype^10^, or assume perfect specificity and require knowledge of the true but often unknown prevalence within the sample^1^. In contrast, we introduce a novel methodology, PheMED (<u>Phe</u>notypic <u>M</u>easurement for <u>E</u>ffective <u>D</u>ilution), to assess the relative dilution between GWA studies without the above limitations. Furthermore, since PheMED only uses summary statistics, our method is applicable even when researchers do not have access to individual level data. To develop the theory for this new tool, PheMED, we leverage the fact that phenotypic misclassification bias shrinks the estimated effect size of SNPs across all tests by the same multiplicative value^11^. And since a GWA study typically comprises more than 200,000 approximately independent tests of common genetic variants^12^, we can learn the effective dilution by comparing summary statistics across studies.

Although our motivation and theoretical derivation for our analyses comes from phenotype misclassification bias, we would like to emphasize that effect size dilution can be driven by other biases as well. For example, cohort prevalence differences in environmental confounders (e.g. carcinogen exposure in cancer or head trauma in neuropsychiatric traits), can also dilute the effects of genetic risk factors when predicting a given trait. Consequently, PheMED provides a composite measure of the *effective dilution*, which can capture other sources of bias, in addition to phenotype misclassification bias. In our work, we demonstrate the utility of leveraging an inclusive measure of effect size dilution to inform data-driven phenotyping as well as downstream polygenic risk score, meta and power analyses.

At this juncture, we outline our main contributions to the field. First, we formulate conditions illustrating how the effective dilution relates to the positive and negative predictive value of the labeled phenotype^11,13^ and extend the analysis even when a gold standard phenotype is unavailable. Since reported sample sizes obscure the number of correctly labeled cases and controls, we propose a dilution adjusted effective sample size, allowing researchers to better compare the contribution of different cohorts with varying levels of data quality. We demonstrate how effective dilution is distinct from genetic correlation, can confound heritability estimates and diminish power in GWA studies. Furthermore, we bolster the relevance of our theoretical results by applying our approach to multiple use-cases within the literature, such as cross-ancestry and cross-cohort analyses. We demonstrate that in many cases, directly comparing GWA studies for the same phenotype is problematic due to differences in rates of misclassification between study populations.

Since phenotypic misclassification dilutes the effect sizes in a study, meta-analysis using standard inverse-variance weights (IVW)^14^ incorrectly assumes that the effect sizes from different studies come from the same distribution. To address this issue, we propose a methodological extension of the IVW meta-analysis using dilution-adjusted weights (DAW), to adjust the weight of each study according to its effective dilution. We show that directly incorporating the estimated effective dilution into the statistical test can substantially increase performance and rescue the loss of power on real-world data. We perform simulations, indicating that DAW increases power in GWA studies without increasing the false positive rate. Consequently, we anticipate that our proposed methodology, PheMED, for measuring effective dilution will provide a valuable tool when comparing the results of similar phenotypes on account of data quality heterogeneity. This, in turn, will both aid in evaluating phenotyping strategies across populations and allow for dilution-aware meta-analyses.

## Results

### Estimating effective dilution from the data

When we misclassify cases as controls, the similarity between the two groups increases, resulting in diluted effect size estimates. We can measure the effective dilution, which we define as φ_*MED*_- the <u>Phe</u>notypic (φ) <u>M</u>easured *<u>E</u>ffective <u>D</u>ilution* (or relative effective dilution in short) - between the different studies, through a maximum likelihood approach across randomly selected approximately independent loci (see **Methods**).

Briefly, denote *𝒩* (*β* | *μ, σ*^2^ ) as the output of the normal probability distribution function for an observation β, given mean and variance parameters, μ and σ^2^. Mathematically, our log-likelihood function has the form,

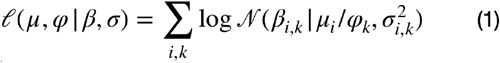

where i indicates the corresponding SNP, k indicates the corresponding study and β_*ik*_, σ_*ik*_, the estimated effect sizes and standard errors, are estimated values available in the summary statistics for each study. Intuitively, equation (1) tells us that we leverage the additional parameter φ_*k*_ to shrink (or grow) all of the true effect sizes of study k based on the quality of the phenotypic data. Under the general setting of k studies, we find the value for the φ_*MED*_ = ( φ_*k*_, …, φ_*k*_ ) that maximizes the likelihood in equation (1). To ensure uniqueness of the solution, we impose a normalization constraint on φ_*MED*_, where φ_1_ = 1, and the other values φ_2_ …, φ_*k*_ measure the relative effective dilution in reference to Study 1. We note that this framework allows for the possibility that no change in our effect sizes is required, since φ_*k*_ can equal 1. We can directly test the null hypothesis of no dilution, or φ_*k*_ = 1, using a bootstrap approach and report an approximate p-value (see Methods). Importantly, as demonstrated later in this work, incorporating additional studies decreases the uncertainty of the φ_*k*_ estimates and thus increases power to reject the null hypothesis (see Supplement). Consequently, PheMED provides us with a pipeline where we can input summary statistics and output effective dilution values, their p-values and confidence intervals for each study (**Fig. 1a**).

**Fig. 1.**
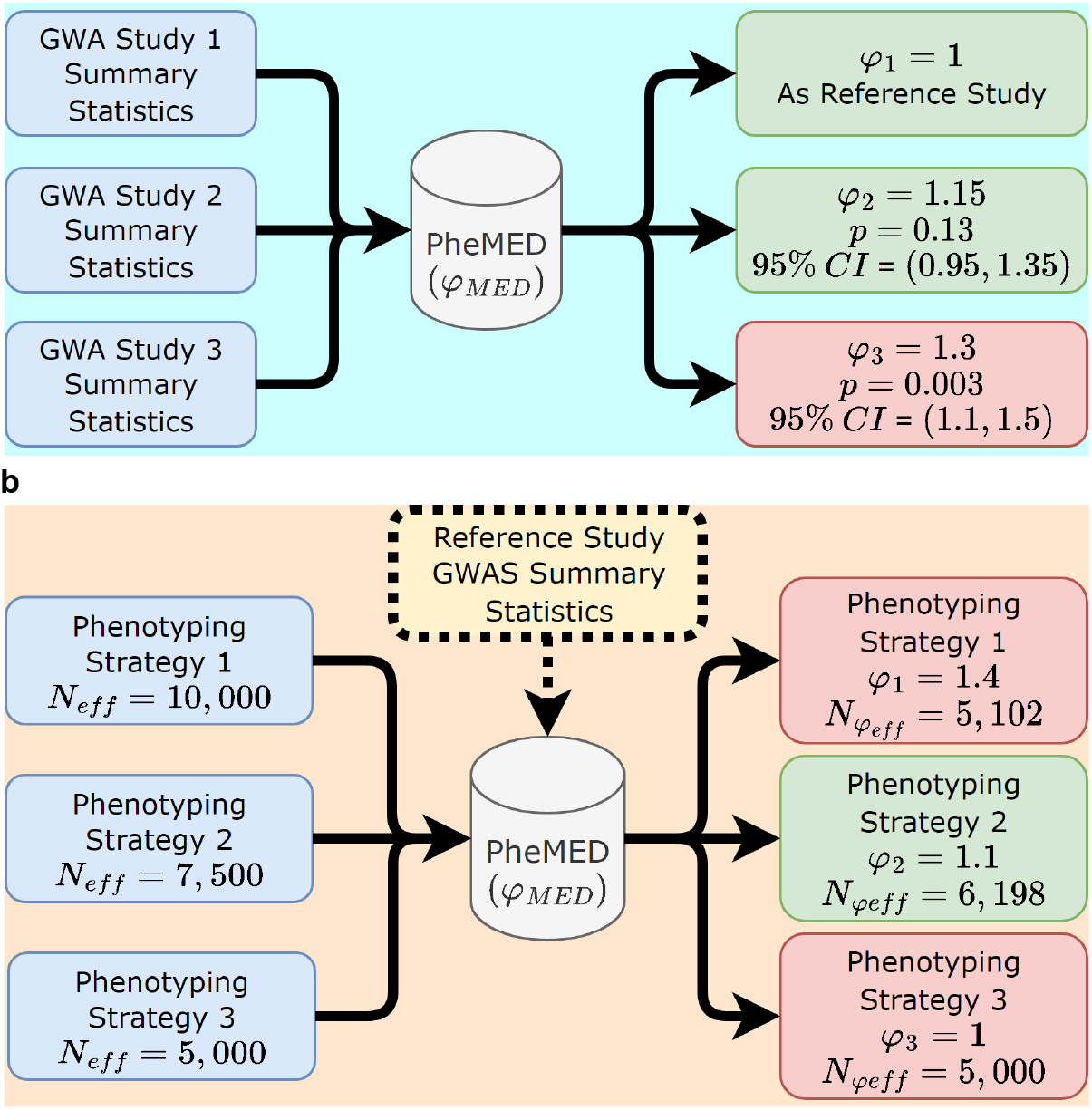
PheMED Pipeline. **a)** GWAS summary statistics for studies 1, 2 and 3 (where n_studies_ ≥ 2) were used as input for PheMED to estimate effective dilution values (φ), confidence intervals (CI) and corresponding p-values. In this example, study 3 exhibits statistically significant effective dilution (φ = 1. 3, *p* = 0. 003) compared to study 1. Study 1 was internally used as reference (φ_1_ = 1). **b)** In order to optimally increase power for GWAS, phenotyping strategies have to increase sample size (*N*_*eff*_ ) while containing phenotypic dilution (φ). PheMED provides researchers with a tool to estimate each study’s dilution-adjusted effective sample size 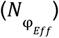 which can be used as an optimization parameter; higher 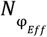 results in higher power. In this example, Phenotyping Strategy 2 strikes a good balance of flagging potential cases without overdiluting the phenotype. If there are overlapping samples among the different phenotyping strategies, PheMED has to be run separately for each phenotyping strategy against summary statistics from a non-overlapping sample (“Reference Study”).

In addition, we can leverage the estimated effective dilution (φ_*MED*_ ) to compute a dilution-adjusted effective sample size: 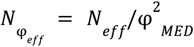 (See supplement for derivation). This notion of a dilution-adjusted sample size can then be used to compare different phenotyping strategies and inform the researcher regarding the strategy that yields the greatest dilution-adjusted effective sample size, 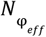 . We note that if the samples are overlapping, a separate non-overlapping sample must be used to estimate the dilution for each strategy (see **Methods, Fig 1b**). In the following sections, we discuss the theoretical implications of φ_*MED*_, and explain its connection to phenotypic misclassification in further detail.

### Interpreting the effective dilution

In the supplement, we provide a derivation relating the effect sizes, β _*diluted*,1_, β _*diluted*,2_ of two (possibly) diluted GWA studies. Specifically, we demonstrate that,

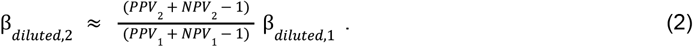

where PPV and NPV stand for the positive and negative predictive value of the labels under the phenotype. The numerator (or denominator), PPV + NPV - 1, found in equation (2) is often referred to the ‘markedness’ or Δp of the possibly imperfectly labeled phenotype in the corresponding study^15^. Observe that the multiplicative factor relating the two GWA studies, only depends on the positive and negative predictive values of the diluted phenotype in the respective studies. We note that the derivation in the supplement depends on the assumptions from Beesley et al.^1,13^, where effect sizes β are small and the mislabeling is independent of other confounders.

Let the factor relating β _*diluted*,1_, β_*diluted*,2_ be defined as

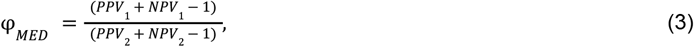

so that, β _*diluted*,2_ ≈ β _*diluted*,1_ /φ_*MED*_ . By this definition, increasing relative effective dilution corresponds to a smaller expected effect size estimate in the second study. Importantly, the dilution depends on the *relative* misclassification between the studies. Studies with large but equivalent misclassification do not result in dilution of effect size estimates.

In the special case where we have a gold standard phenotype, from equation (2) our algorithm estimates the markedness (Δp) of the diluted phenotype. In practice, however, two similar phenotypes may in fact be slightly different, as different samples could possess different degrees of environmental confounders or misclassified patients could have a different diagnosis and such misclassification might not be representative of a typical control patient. Alternatively, different studies could adjust for different confounders. Nevertheless, for the purposes of our work, we simply refer to the correction factor as φ_*MED*_ .

Furthermore, as evidenced by equations (2)-(3), since phenotypic misclassification can bias our effect size estimates and we often suspect different levels of phenotypic misclassification across different studies, we will demonstrate the importance of computing the effective dilution for a variety of use-cases even when the aforementioned assumptions may not be fully met.

### Heritability and co-heritability in the presence of dilution

Since the relative effective dilution φ_*MED*_, impacts the estimates of the effect sizes, β, in a GWAS, it will affect the heritability estimates. Intuitively, heritability refers to the variance explained by the risk allele predictors, and when our effect sizes are small, (e.g. our log odds ratios are near 0), the risk alleles capture very little information about our phenotype, which translates to low heritability values.

Mathematically, assuming standardized genotypes the observed SNP-heritability can be represented as a weighted sum of the effect sizes^16–18^ : 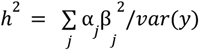, where the (possibly unknown) weights α in the numerator allow us to account for any linkage disequilibrium and the denominator measures the variance of the phenotype y.

Therefore diluting the effect sizes by a finite, positive factor of φ_*MED*_ according to 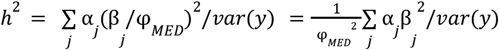, reduces the observed heritability by a factor of φ_*MED*_ ^2^.

Yet, surprisingly, dilution does not affect the estimated genetic correlation, *r*_*g*_, between studies for plausible values of φ_*MED*_ . Given the same assumptions, the genetic correlation between two traits is the correlation between the effect sizes^19^. Since *r*_*g*_ is based on correlation, which is scaling invariant, dilution does not affect *r*_*g*_ . More precisely, if we measured the correlation between effect sizes from randomly selected and approximately independent loci corresponding to two different GWASs, β_*GWAS*1_, β_*GWAS*2_, we observe that *r*_*g*_ = *cor*(β_*GWAS*1_, β_*GWAS*2_ ) = *cor*(β_*GWAS*1_, β_*GWAS*2_ /φ_*MED*_) . Thus estimates of genetic correlation are independent of the effective dilution (**Fig. S1**).

Importantly, when evaluating the similarity between summary statistics from two studies, *r*_*g*_ and φ_*MED*_capture complementary and statistically independent features of the data. *r*_*g*_ reflects the correlation in effect size estimates, while φ_*MED*_reflects the relative scaling between the effect size estimates.

### Simulation results

We evaluate this approach in the GWAS setting through simulation, consisting of 100,000 SNPs, where 90% of SNPs correspond to null effects and the remaining 10% of SNPs have very modest effect sizes. We specify different values for the PPV, NPV, expected size of true effects and sample sizes for the diluted and gold standard GWAS studies (See Methods for details). In particular, we find the maximum likelihood approach performs well in inferring the true effective dilution value, φ_*MED*_(**Fig. 2A**), and produces well-calibrated p-values under the null model of no dilution (**Fig. 2B, S2**) In the supplement, we consider the impact of substituting patients with related but different traits as cases and controls. In this scenario, we provide both mathematical analysis and simulations demonstrating that dilution provides a weighted measure of phenotypic data quality (markedness) based on the genetic similarity of the misclassified patients (**Fig S3, Supplementary Methods and Results)**

**Fig. 2.**
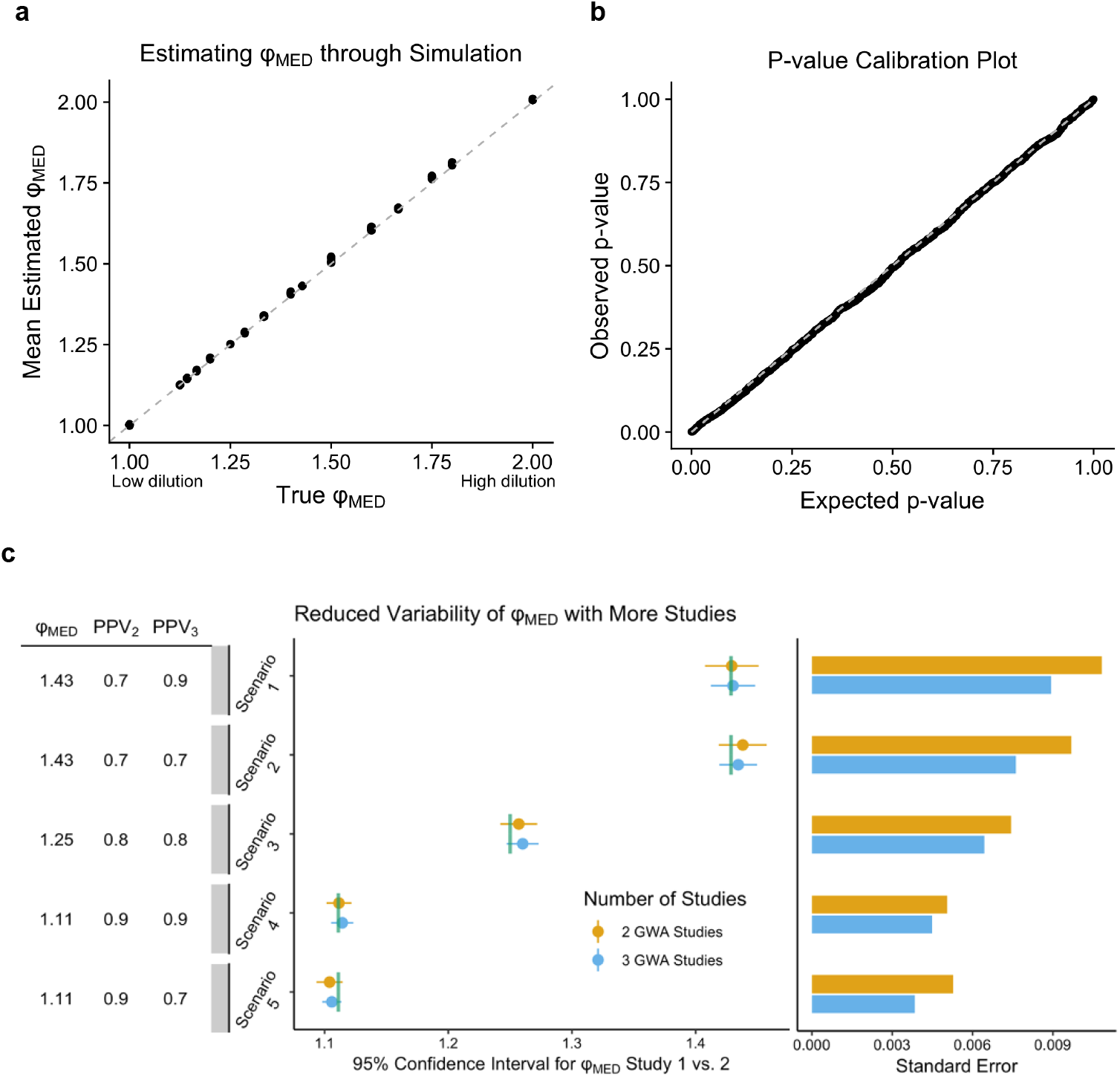
Measuring effective dilution in simulation. Panel **a** compares the estimated effective dilution φ_*MED*_with the true value in simulated GWA studies over 2,000 trials. The true effective dilution is defined as the ratio of the markedness values between the two GWA studies. The gray dashed line indicates when the estimated value for the effective dilution equals the true value. Standard errors of means for all parameter choices are too small to be visualized in this figure. Panel **b** compares the distribution of 2,000 computed p-values when there is no dilution between the two studies on the y-axis, with the expected uniform distribution of p-values on the x-axis, since the null hypothesis is true. The gray dashed line indicates that the expected p-value equals the observed p-value on the y-axis. Panel **c** describes a simulation study for computing 95% confidence intervals to estimate φ_*MED*_ for different GWA studies. One analysis, in orange, estimates φ between 2 GWA studies, whereas the other analysis, in blue, can also leverage additional information from a third GWA study to estimate φ_*MED*_ between the 2 aforementioned GWA studies. The vertical green line indicates the true value for φ_*MED*_ . The table of parameters on the left indicates the PPV for Studies 2 and 3. The bar plot to the right indicates the standard error in each simulation scenario. Study 1 is simulated so that it has PPV = 1 and all studies have NPV = 1. For ease in visualization, we only provide the confidence intervals for 50 of the 2,000 different simulations.

We also investigate the width of our confidence intervals as we increase the number of GWA studies in our pool. In particular, we find that the confidence intervals for measuring effective dilution narrow as we include more studies (**Fig. 2c, Table S1**). Thus, as we reduce our uncertainty regarding the true effect sizes, we attain greater precision regarding the range of plausible values for the effective dilution between phenotypes from different GWA studies in our simulations. Notably, for our simulation study, the standard errors for the mean effective dilution shrunk between 10-21% across the five simulation scenarios, *p* < 5. 74 × 10^−10^ for all simulation scenarios, one-sided chi-square test for the variance).

Finally, we provide simulations indicating that dilution is robust to population stratification and cryptic relatedness (**Fig. S4-S5, Table S2, Supplemental Methods**). These results highlight that PheMED measures a form of bias, independent of the biases that are measured through other summary statistics methodologies like LD Score Regression^20^. We note that differences in sample prevalence do impact the observed heritability, as sample prevalence influences the variance of the phenotype. Nevertheless, these differences in sample prevalence do not affect the dilution, as the log odds ratios are independent of the prevalence of the phenotype in the sample (**Fig. S6**).

### Applications: Use cases for effective dilution

We briefly describe three real-world use cases where we use our method to evaluate: *(a)* different phenotypic definitions within the same cohort, *(b)* cross-ancestry differences within the same cohort using the same phenotypic definition and *(c)* differences across discrete cohorts. For each of our use cases, we estimate the effective dilution between the different GWA studies with our proposed method PheMED.

#### Different phenotypic definitions within the same cohort

First, we consider effective dilution between two separate GWA studies using the presence of one (1; lenient) or at least two (2+; strict) phecode counts for bipolar disorder in the Million Veteran Program (MVP), leveraging non-overlapping patients (and controls) for each analysis. As expected from our prior work^21^, we observe significant effective dilution in the more lenient definition ( φ_*MED*_ = 1. 52, *p* = 1. 35 × 10^−13^, bootstrapped difference of means, see Methods for details) (**Fig. 3, Tables S3, S4**). In a similar thematic fashion, we measure the effective dilution between two GWA studies with different severity levels in the MVP, where the lenient case definition only requires the presence of at least two obesity phecodes, while the stringent case definition requires the presence of at least two morbid obesity phecodes, where patients have a BMI exceeding 35. Since these GWA studies came from overlapping samples, we estimate the effective dilution between the two phenotypes by estimating effective dilution of both against an independent study (see Methods). In this case as well, we identify significant effective dilution in the more lenient definition (φ_*MED*_ = 1. 16, *p* = 7. 30 × 10^−6^, bootstrapped difference of means; **Fig. 3, Tables S3**,**S4**).

**Fig. 3.**
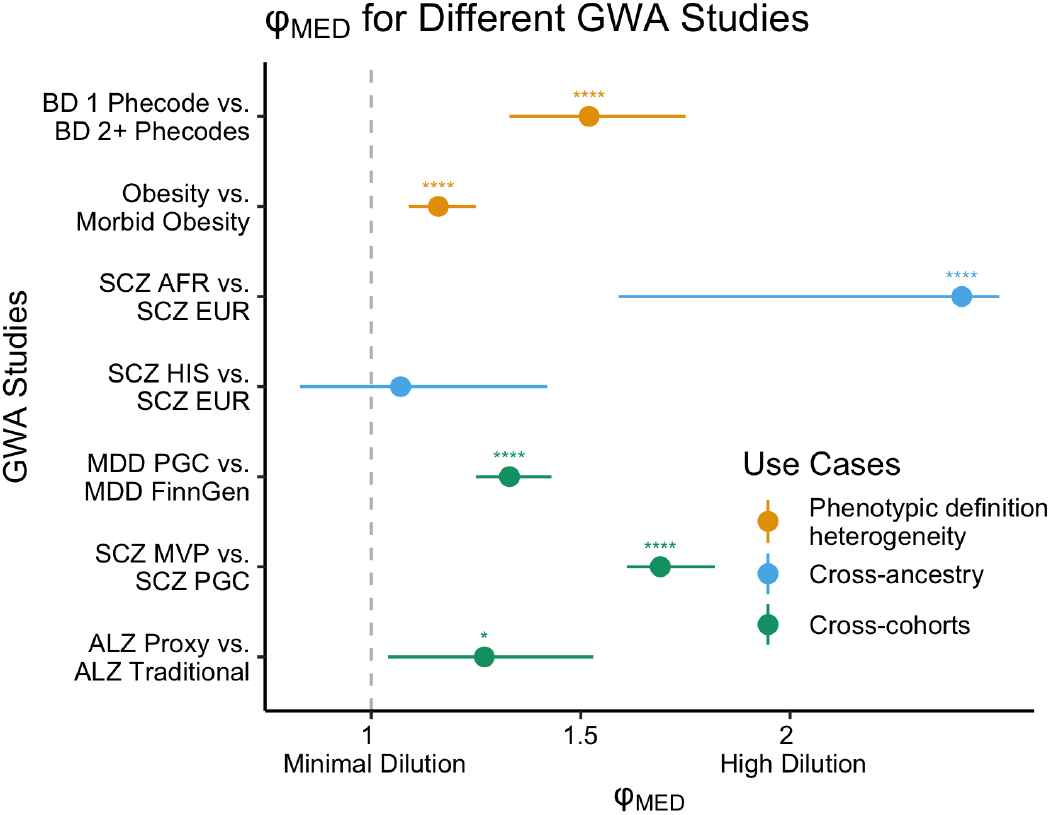
Effective dilution across different use cases. Examples of effective dilution (φ_*MED*_) for different use cases: different phenotypic definitions within the same cohort (orange; e.g. BD 1 Phecode vs. 2 Phecodes), cross-ancestry analyses (blue; e.g. SCZ AFR vs. EUR) and cross-cohort analyses (green; e.g. SCZ PGC vs. MVP). For a full list of values and confidence intervals see Table S2. \**p* < 0.05. \*\**p* < 0.01. \*\*\**p* < 0.001. \*\*\*\**p* < 0.0001. Note that the “ALZ Proxy” GWAS encompasses a mixture of both proxy cases from the UK Biobank as well as more conventionally defined cases; however, for notational simplicity, we denote this study as “ALZ Proxy”. In contrast the Alzheimer’s GWA Study from FinnGen only utilizes conventionally defined cases, and we denote this study as “ALZ Traditional”.

#### Cross-ancestry differences within the same cohort

We estimate effective dilution for schizophrenia (2+ phecodes) in individuals of African (AFR) and Hispanic (HIS) ancestry when compared to individuals of European (EUR) ancestry in the same cohort (MVP). We find significant effective dilution for AFR (φ_*MED*_ = 2. 41, *p* = 1. 59 × 10^−9^ ) but not HIS ( φ_*MED*_ = 1. 07, *p* =. 63; **Fig. 3, Tables S3, S4**) ancestry individuals. Notably, the finding of significant effective dilution for AFR ancestry patients, aligns with existing literature indicating that Black patients with mood disorders are more likely to be misdiagnosed with schizophrenia^2,3^; as we anticipate a correlation between the distinct concepts of genetically-inferred ancestry and race^22,23^. However, we caution that hidden confounders, such as socioeconomic status and other environmental factors, can bias heritability estimates of a trait and dilute effect sizes as well^24,25^.

#### Differences across independent cohorts

In addition to the aforementioned analyses, we consider effective dilution across different cohorts, leveraging an existing Major Depressive Disorder GWA study from FinnGen along with a multi-cohort meta-analyzed GWA study from the Psychiatric Genomic Consortium (PGC)^26^, which includes 127,552 self-reported cases from the UK Biobank (**Fig. 3, Tables S3, S4**). As we anticipate self-reported data to be less accurate than coded diagnoses in electronic medical records, we observe statistically significant effective dilution between the studies (φ_*MED*_ = 1. 33, *p* = 1. 34 × 10 ^−21^). We also compute effective dilution between an existing schizophrenia GWA study from the PGC^27,28^ and a GWA study from Release 4 of the MVP (**Fig. 3**, Tables S2, S3) where we attain a significant effective dilution value (φ_*MED*_ = 1. 69, *p* = 1. 05 × 10^−121^ ), supporting the existence of data quality heterogeneity between the different studies. And finally, we assess the effective dilution between two GWA studies for Alzheimer’s disease: *a)* a study which consists of a large percentage of proxy cases (individuals with family history of the disease) from the UK Biobank^29^ (denoted ALZ Proxy), and *b)* a study that doesn’t utilize a proxy case definition (FinnGen^30^; denoted ALZ Traditional). When comparing studies that incorporate different percentages of traditional cases, we identified a significant effective dilution value (φ_*MED*_= 1. 27, *p* = 0. 028), supporting the notion that proxy cases have lower positive predictive value than labels derived from a healthcare-based diagnostic code definition. Note that we anticipate the effective dilution between the proxy case phenotype from the UK Biobank and FinnGen to be higher than 1.27, as the meta-analyzed GWA study used above (ALZ Proxy) includes a substantial percentage of cases that were ascertained very rigorously (e.g. with definitive diagnoses using CERAD scores).

We wish to stress the perhaps surprisingly extreme values of effective dilution that were found above. To conceptualize this, suppose that our labeled phenotypes had a perfect negative predictive value and we meet the assumptions such that we can interpret effective dilution as a ratio of Δ*p* values between the different studies. To achieve effective dilution values of 2.41, that would suggest that our positive predictive value is less than half of the reference study. These findings highlight the utility of computing the effective dilution as a quality control metric when comparing GWA studies using different phenotypic definitions both within and across cohorts.

### Jointly measuring effective dilution over multiple GWA summary statistics increases precision of φ_*MED*_ estimation

Here, we illustrate the advantage of jointly measuring effective dilution across multiple studies on real-world data. Similar to our simulations from **Fig. 2c**, we also observe narrower confidence intervals when we compute effective dilution through a joint tri-ancestry analysis as opposed to separate bi-ancestry analyses. The width of our 95% confidence interval (95% CI) for estimating φ_*MED*_ from the AFR GWAS shrinks from (1.65, 5.26) to (1.59, 3.83). We observe similar, but less dramatic decrease in width for the 95% CI for estimating φ_*MED*_ from the HIS GWAS, where the confidence interval shrinks from (0.83, 1.63) to (0.82, 1.42), (**Fig. 4**).

**Fig. 4.**
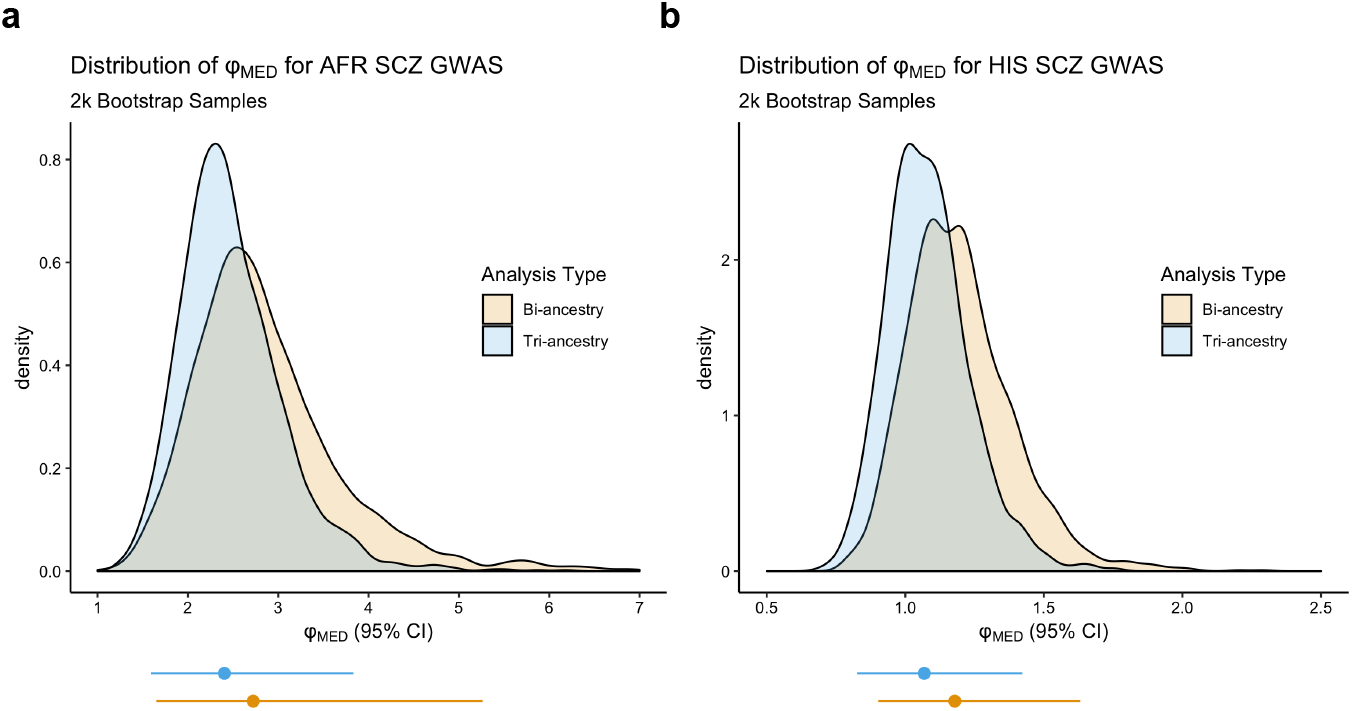
Reduced variability when jointly estimating effective dilution over multiple studies. Empirical bootstrap distribution of effective dilution for (a) African ancestry and (b) Hispanic ancestry for 2,000 circular bootstrap samples where the bi-ancestry analysis is highlighted in orange and the tri-ancestry analysis is highlighted in blue. 95% confidence intervals corresponding to these distributions are plotted below the x-axis. When adding a third ancestry into the analysis, Hispanic and African for (a) and (b) respectively, the confidence intervals for φ_*MED*_ shrink.

### Impact of dilution on heritability estimates

Since heritability provides us with a theoretical upper bound for the performance of leveraging polygenic risk scores to describe phenotypic variation^31,32^, computing accurate heritability estimates provides insight regarding the potential clinical relevance for generating polygenic risk scores in a clinical setting. As such, we follow up with heritability analysis across cohorts. We find that significant dilution can lead to incompatible SNP heritability (h^2^ _SNP_) estimates for the same trait (**Fig. 5)**, even when accounting for 95% confidence intervals (95% CI), as predicted by our theoretical derivation. This, in turn, affects heritability estimates for downstream inverse variance weighted (IVW) meta-analyses, resulting in loss of information regarding the “true” SNP effect sizes compared to optimally phenotyped cohorts (**Fig. 5**). Phenotypic misclassification can lead to extreme variability in h^2^ _SNP_ estimation and for many traits heritability estimates have been historically inconsistent (see Table **S5** for examples). We anticipate that systematic bias due to misclassification is playing at least a partial role in contributing to these inconsistencies as evidenced in our own schizophrenia and bipolar disorder examples (**Fig. 5**).

**Fig. 5.**
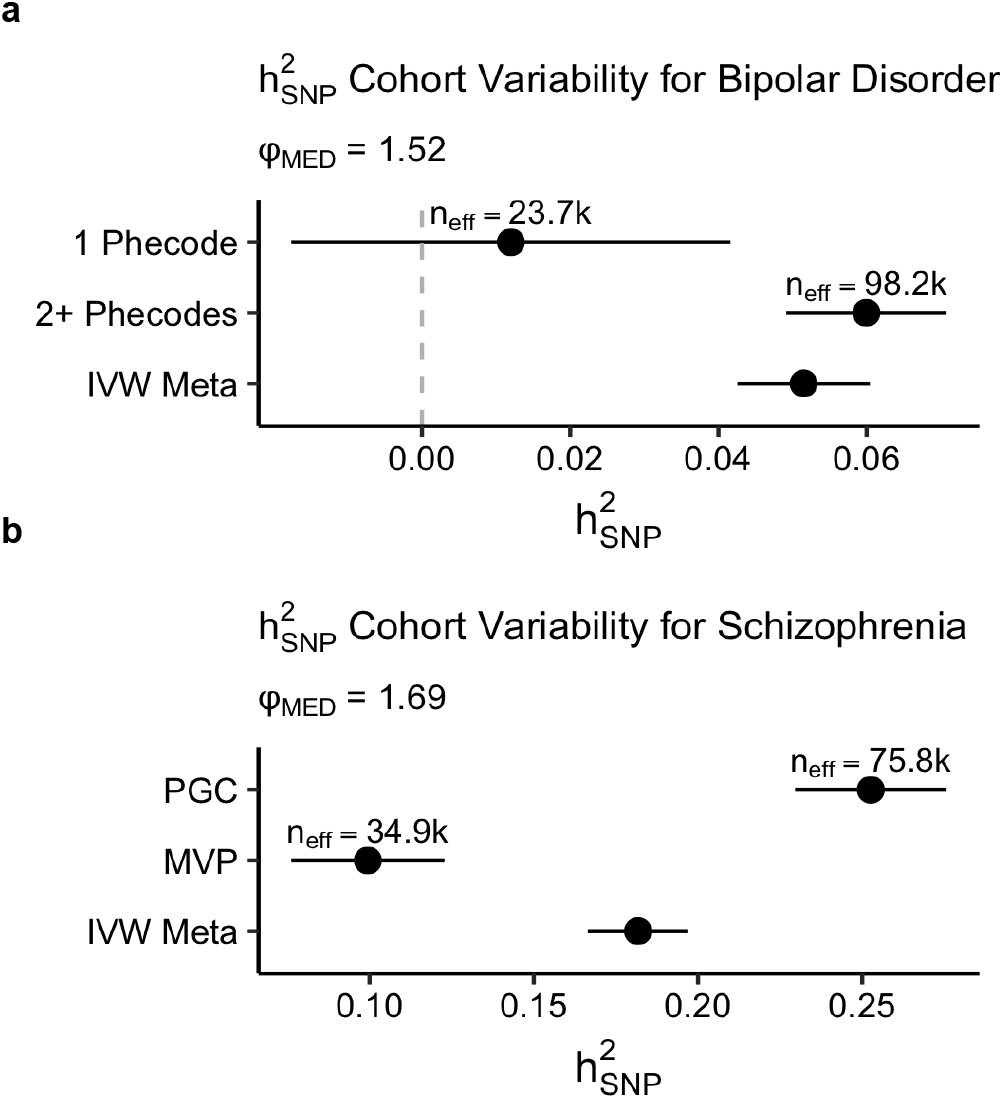
Effective dilution impacts heritability estimates. Confidence intervals for SNP-heritability across GWA studies for (**a**) two different definitions of Bipolar Disorder in the Million Veteran Program (MVP) and (**b**) Schizophrenia in the PGC and MVP cohorts. In both cases, using a diluted phenotype will yield incompatible (non-overlapping) confidence intervals for heritability and performing a meta-analysis of a diluted and non-diluted phenotype will inappropriately shrink the SNP-heritability. Assumed population prevalence for computing heritability on the liability scale was 2% for Bipolar Disorder and 1% for Schizophrenia. Effective sample sizes 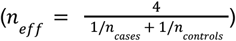 are provided for each study.

### Effective dilution is independent from and complimentary to genetic correlation

Subsequently, we run LD-score regression to compute the genetic correlation between our traits with significant effective dilution on our EUR ancestry cohorts. As displayed in **Fig. 6**, we observe that our schizophrenia and bipolar disorder studies can yield both high genetic correlation and significant values for effective dilution as well. Our computational results support the theoretical results derived earlier in our work, that a given trait can both have high genetic correlation and significant dilution with respect to another similar but more precise definition of the trait. In addition, estimates for genetic correlation from LDSC can yield implausible values for modest sample sizes, whereas we can attain robust estimates from effective dilution on the same sample size with PheMED. Consequently, we recommend that researchers measure both the effective dilution, through PheMED, and compute the genetic correlation between different GWA studies to assess the validity of comparing the results from different studies.

**Fig. 6.**
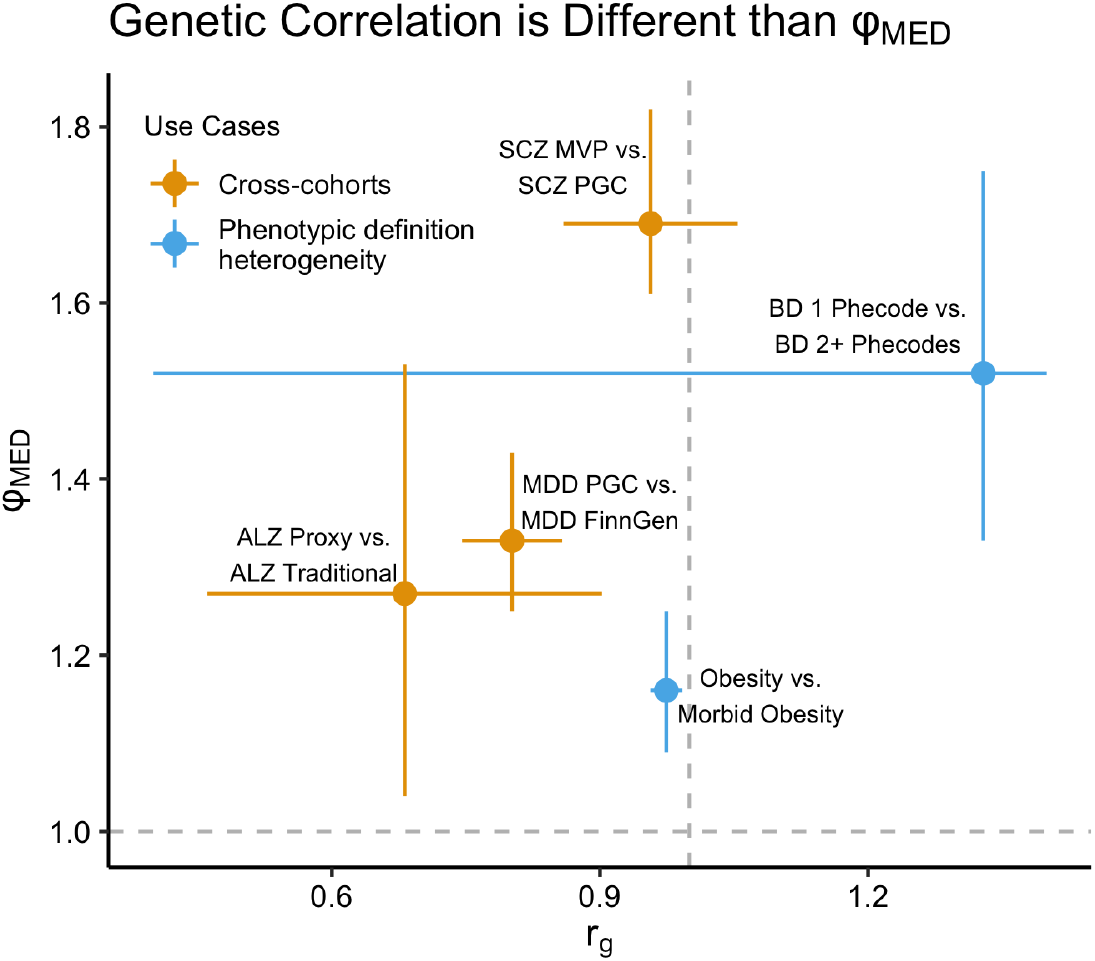
Genetic correlation (*r*_*g*_) does not correspond to effective dilution (φ_*MED*_). The x–axis plots the genetic correlation and 95% confidence intervals (95% CI), while the y-axis plots the effective dilution and 95% CI. Ideally, GWAS for the same trait should have genetic correlation near 1 and effective dilution near 1 (corresponding to horizontal and vertical dashed gray lines).

### Effective dilution can compromise validation and replication efforts

We also note that since effective dilution influences the effect size estimates, we anticipate that dilution can change power estimates for SNP replication in an independent sample. We provide a mathematical derivation illustrating how power is inversely related to dilution (**Supplementary Results**) and provide a visualization of the results in **Fig. 7a**. In particular, for replication of a single index SNP, we plot different power curves corresponding to 5% significance, where we fix the standard error to be 0.0125. We find that the SNP association effect size necessary for 80% power varies substantially as we change the effective dilution. For example, in the presence of no dilution, the effect size necessary for 80% power is 0.031. But with an effective dilution of 2, the effect size must be at least 0.062 to achieve the desired 80% power for replication. Naturally, our analysis can be extended to other significance levels as desired towards accurately estimating the power for replication.

**Fig. 7.**
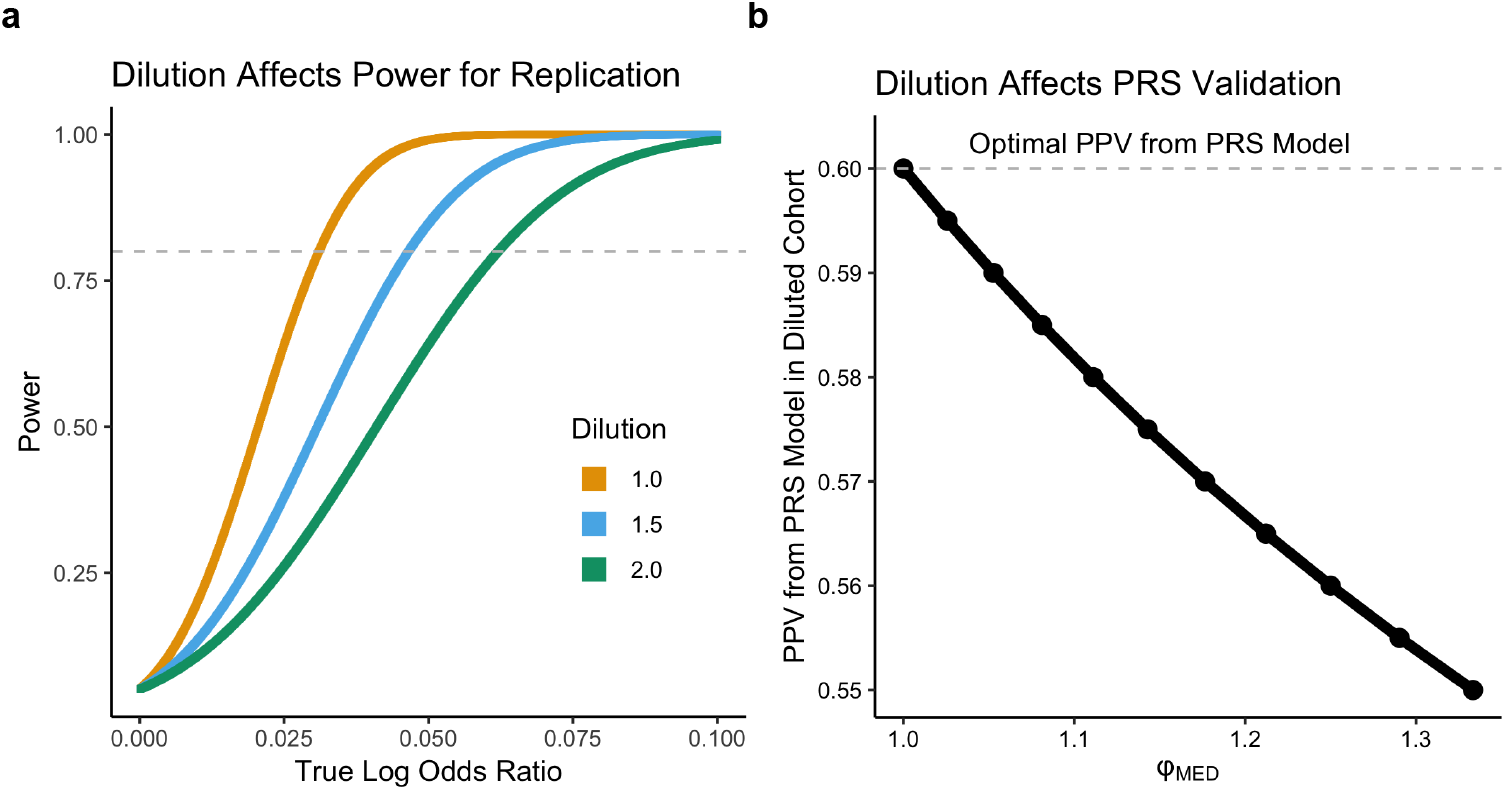
Dilution affects power for replication and PRS validation. **a**. We plot power curves for replicating SNPs with different true log odds ratios (x-axis) with different effective dilution values in different colors. We set the standard error to be fixed at 0.0125 and defined the significance threshold to be at 5% using a one-sided Z test. The gray dashed line signifies 80% power. To achieve 80% power with no effective dilution, we require the true effect to be 0.031. In contrast, if we have an effective dilution value of 2, the true effect would need to be 0.062. **b**. We plot how effective dilution, φ_*MED*_, can bias the observed PPV from a polygenic risk score model compared to a validation cohort with no dilution. The x-axis denotes φ_*MED*_ . The y-axis indicates the PPV from the polygenic risk score model in the diluted cohort and the dashed gray line identifies the PPV from the polygenic risk score model under no dilution. For simplicity, we fix the NPV of the labeled phenotype at 1 and allow the PPV of the labeled phenotype to vary.

Phenotype dilution can also affect the performance of polygenic risk scores (PRS). After training a PRS model on a perfectly encoded phenotype, dilution in the validation dataset can substantially reduce the PPV and NPV of the predictor (**Supplementary Results**). In **Fig. 7b**, we plot the observed PPV from a PRS model as a function of φ_*MED*_, where the optimal *PPV* _*PRS,Optimal*_ = *NPV* _*PRS,Optimal*_ = 0. 60. As expected, when there is no dilution, *PPV* _*PRS,Observed*_ = *PPV* _*PRS,True*_. However, even for modest levels of dilution, where φ _*MED*_ = 1.33, we find that our observed PPV from the PRS model, *PPV* _*PRS,Observed*_ = 0. 55, a 5 point decrease from *PPV* _*PRS,Optimal*_ .

### Accounting for effective dilution in meta-analysis

When different studies possess effect-size heterogeneity due to phenotypic misclassification bias, the assumptions of the standard fixed-effect meta-analysis using inverse variance weights (IVW) or Stouffer’s z-score method with sample-size weights (weighted Z) are violated. Consequently, just as we can use our knowledge of φ_*MED*_ to compute more robust power computations and PRS validation metrics, we can also employ φ_*MED*_ to meta-analyze diluted phenotypes. Here, we propose the dilution adjusted weights (DAW) meta-analysis methodology, an extension of the fixed effects meta-analysis framework, where we account for effect size heterogeneity by adjusting the weights of each study according to their effective dilution (see **Methods** for details). We compare the performance of our new methodology against four other meta-analysis techniques, including IVW^14^, Weighted Z^14^, random effects meta-analysis (REMA)^33^ and MTAG^34^.

We first compare the number of FDR-significant hits for five different use-cases and find that DAW consistently identifies more FDR-significant loci than competing meta-analysis methodologies (**Fig. 8, Methods**). Furthermore, we find that DAW is consistently competitive or outperforms competing methodologies when comparing the number of significant hits that are validated through an independent cohort as well (**Tables S6-S12**). We then perform a simulation analysis, where we both demonstrate that DAW controls the Type I error rate and flags more significant hits than competing meta-analysis methodologies (**Fig. S7, S8**). We perform additional simulations on real world genomic data from the Million Veteran Program, comparing MTAG with PheMED/DAW. Notably, we find that while PheMED/DAW can recover the true dilution values even when within-ancestry population stratification was not adequately adjusted for (**Table S2; Fig. S4, S5**), MTAG fails to recover the correct dilution values due to methodological limitations in accommodating GWASs with modest sample sizes, which diminishes the power when performing meta-analyses (**Fig. S9-S12, Supplementary Methods, Supplementary Results)**. We also apply our meta-analysis methodology to transcriptome-wide association studies and DAW increases power in that setting as well (**Fig. S13, Methods**). Finally, we stress that DAW corrects dilution at the GWA summary statistics level; notably, since publicly available GWA summary statistics often reflect an aggregation of multiple cohorts and varying phenotype definitions, researchers ideally should generate separate GWA summary statistics for different phenotypic definitions and run PheMED to assess the data quality of the phenotypes, as illustrated earlier in **Fig 1b**.

**Fig. 8.**
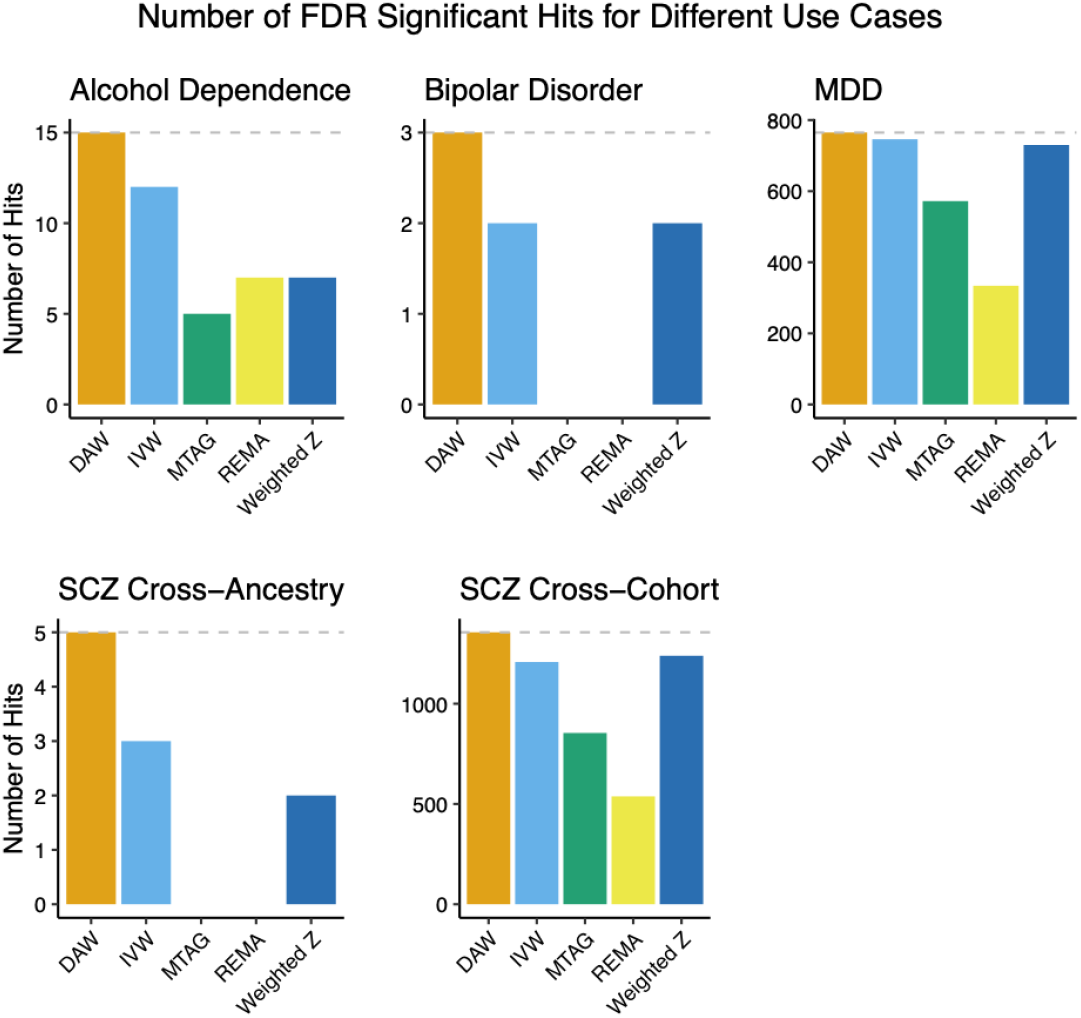
Comparing dilution adjusted weights meta-analysis (DAW) with competing methodologies. Number of FDR Significant Hits (y-axis) found by different meta-analysis methodologies (x-axis) for different use-cases (facets). The gray dashed line indicates the maximum number of FDR significant hits identified by the best performing methodology for each use case. To assess the number of hits, only approximately LD-independent SNPs were used (see methods for details on the clumping procedure). Note that for SCZ Cross-Ancestry, MTAG was not performed as a cross-ancestry analysis violates the assumptions of the model.

## Discussion

Whenever we compare phenotypes from different data sources, we should anticipate data quality heterogeneity. In particular, we recommend caution when leveraging large samples from biobank data, as the true effective sample size can be much smaller, depending on the level of dilution from the data acquisition process. Consequently, we develop a new method, PheMED, to detect genome-wide effect size dilution across GWA studies for the same trait, using only summary statistics from the GWA study. The main underlying assumption for our approach is that common SNP-trait association effect sizes are comparable across populations and ancestries in well-controlled studies at the genome level, although there are exceptions, e.g. ancestry-specific associations at specific loci^35^. We stress that PheMED is unique from other heterogeneity tests, where our test specifically looks for genome-wide dilution across all effect sizes in a GWAS, and does not require knowledge of genome-wide significant loci from other cohorts to assess for dilution-driven heterogeneity^36^. We showcase several real-world scenarios where our methodology benchmarks phenotypic quality governed by potential sources of misclassification errors, such as different phenotypic definitions and healthcare disparities within the same cohort, as well as cross-cohort heterogeneity. In particular, we find instances of statistically significant dilution in all of the aforementioned use-cases, suggesting different levels of data quality across different GWA studies. We demonstrate that our methodology produces narrower confidence intervals for the effective dilution when jointly estimating the effective dilution across multiple studies that share the same phenotype. We refer to this as ‘transitive inference’. Furthermore, we introduce the dilution-adjusted effective sample size to help researchers identify the optimal phenotyping strategy that will produce the greatest power in detecting statistically significant results.

Currently, when justifying the inclusion of a cohort as part of a meta-analysis or establishing consistency between a newer study and a prior analysis, the current accepted practice is to verify that there is a high genetic correlation between the two studies^37–39^. We illustrate that the aforementioned quality check is insufficient, as two GWA studies can simultaneously share both high genetic correlation and strong levels of effective dilution, resulting in diluted effect sizes and heritability estimates from the meta-analysis. We note that prior work has emphasized that cross-study heterogeneity, due to imperfect genetic correlation, can affect heritability estimates^40^. In contrast, our results indicate that heritability can be affected even with perfect genetic correlation on account of phenotypic misclassification.

We emphasize that measuring the effective dilution can also aid researchers in assessing performance of polygenic risk scores and estimating power for replication on external cohorts, as misclassified phenotypes can bias these metrics. Previous work has described how phenotypic misclassification can reduce power^41,42^, but employing such results in practice requires direct knowledge of unknown misclassification probabilities. Nevertheless, all of these aforementioned studies underscore the potential utility of leveraging our methodology to infer the effective dilution of odds ratios from the summary statistics to compute accurate heritability and power computations for GWA studies.

At this juncture, we review the limitations in using PheMED. First, interpreting the effective dilution as a ratio of markedness values (NPV + PPV - 1) can be problematic if the phenotypic misclassification depends on other confounders, if the studies address different phenotypes, adjust for different confounders, or if there are differences in unadjusted environmental effects between the cohorts that impact the heritability of the trait. In such scenarios, PheMED provides an aggregated estimate of dilution across multiple potential sources of bias, analogous to how the intercept in LD Score Regression aggregates the impact of cryptic relatedness, model misspecification and population stratification into a single metric for measuring inflation. We also note that when jointly modeling multiple studies, at least one pair of studies must have no sample overlap. In spite of these limitations, we can relate effective dilution estimates from PheMED to a meaningful metric (ratio of markedness) when interpreting the practical significance of different values for the effective dilution. Consequently, we present a deterministic method for computing the effective dilution to assess if GWA studies are indeed comparable to one another for heritability, meta and replication analyses. And in contrast to prior methods in the literature, estimating the effective dilution through PheMED does not depend on a gold standard GWA study^10^, does not require us to assume values for unknown performance metrics for a phenotype in a given GWA study^1^, and only uses summary statistics to detect effective phenotypic dilution.

Finally, we wish to highlight the utility of leveraging PheMED estimates for meta-analysis, through our newly proposed dilution-adjusted weights (DAW) algorithm. First, our approach allows us to overcome limitations associated with violations of the principal assumptions of existing methods. For example, fixed effects meta-analysis techniques such as inverse variance weights (IVW) and Stouffer’s z-score method (weighted Z) assume that the population effect sizes are fixed across studies. In practice when meta-analyzing diluted phenotypes, the effect size will depend on the quality of the data. Furthermore, even meta-analysis techniques that aim to account for effect size heterogeneity, like random effects, apply the same effect size shinkage across all studies. In truth, the level of shrinkage for high quality studies should be smaller than for low quality studies. A couple of common real-world examples of these violations include: *a)* the equal treatment of phenotypes based on self-reported data vs. multiple coded diagnoses in a patient’s electronic health record (EHR), and *b)* disregarding the potential for algorithmic bias through non-random healthcare disparities cryptically embedded in EHR for e.g. AFR ancestry individuals^43–46^. When leveraging single ancestry studies, we also considered a multi-trait meta-analysis methodology, MTAG^34^, since different studies for the same trait can be considered as different correlated traits on account of measurement error. Specifically when compared with MTAG, DAW allows for transitive inference which increases the precision of our approach with an increasing number of studies, and affords more flexibility by enabling the utilization of summary statistics with low heritability and/or sample sizes because it is not built on top of LD Score Regression. Consequently, we demonstrate both on real-world data and through simulation that DAW meta-analysis consistently achieves higher power for detecting true effects compared to all of the aforementioned methodologies without increasing the false positive rate in simulations. Therefore, we maintain that the DAW meta-analysis framework is a much needed step towards making our analyses more robust against the aforementioned implicit biases that compromise data quality across studies.

In consideration of our findings, we anticipate that PheMED will serve as a valuable approach for integrating GWA studies with phenotypic quality heterogeneity to guide algorithmic phenotyping and correct for diluted phenotypes that can further bias downstream analyses.

## Methods

### Modeling effective dilution

We assume that the standard errors (provided in the GWA summary statistics) are fixed parameters. Without loss of generality, we standardize our GWA studies by stipulating that φ_1_ = 1, so that all effect sizes are measured in reference to the first study. For ease in interpretation on real-world data, reference studies are selected such that either the reference study has a more rigorous phenotypic definition or is expected to have less phenotypic noise. Our approach allows us to solve the general problem: given N GWA summary statistics, setting the first study in the list to be the reference study, produce N - 1 effective dilution values, where the i^th^ value relates the effective dilution of the i^th^ study relative to a reference study.

To achieve this goal, we optimize the log-likelihood as the effect sizes come from approximately normal distributions.

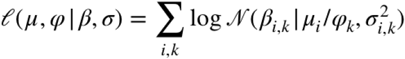

The basic rationale behind this maximum likelihood formulation is that we are applying a study-specific transformation to the overall study means μ_*i*_, for the ith effect (SNP). Here, φ_*k*_ represents how much the true mean for the effect size of the i^th^ SNP shrinks or grows based on the relative phenotypic misclassification rates for study k. For a derivation that phenotypic misclassification is a multiplicative transformation of our means, we refer the reader to the supplement.

Computationally, we express μ as a function of φ, β, σ (as the estimated means from the data are the result of an inverse-variance weights meta-analysis after we know how to transform the GWA summary statistics with φ). We optimize both of these functions by using the Nedler-Mead algorithm as implemented in SciPy^47^, initializing 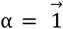.

### Data Processing: Identifying Approximately Independent Loci

To estimate dilution, we perform random clumping, where we identify approximately independent loci by randomly assigning a p-value from a uniform distribution to each SNP so that the loci do not contain any information regarding the effect sizes for any of the GWA studies. From these randomly assigned p-values, we clumped lead SNPs in PLINK^48^ using the European 1k Genomes reference panel^49^. In particular, we required that all SNPs in the same clump must have an R^2^ of at least 0.5 with the lead SNP and be at most 250 kbp away from the lead SNP. We emphasize that our methodology does not require strict independence as we leverage blocked bootstrapping (described later), which is robust to any remaining spatial dependency in our analysis. Furthermore, to ensure robustness for GWA studies that leverage additional ancestries, we performed additional clumping with the surviving SNPs on the African ancestry 1k Genomes reference panel to ensure that the loci were approximately independent across ancestries. For measuring effective dilution in the multi-ancestry schizophrenia GWA studies, we also repeated this procedure for Hispanic ancestry patients as well.

### Estimating Effective Dilution from Studies with Sample Overlap using an Independent Study

Here, we show that we can still estimate the effective dilution between the two studies with sample overlap provided we have access to a third independent study, as performed on our obesity GWA studies with overlapping samples. In particular, suppose that studies 1 and 2 have possible sample overlap, while study 3 has no overlap between studies 1 and 2.

Using the maximum likelihood formulation (5) from the text, we can estimate the effective dilution between studies 1 and 3,

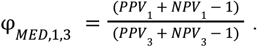

Similarly, we can use the maximum likelihood formulation to estimate the effective dilution between studies 2 and 3,

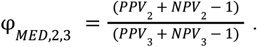

By dividing these two quantities, we attain an estimate for the effective dilution between studies 1 and 2, as

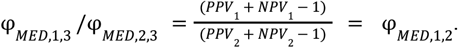

As noted in the text, estimating φ_*MED*_ separately for each pair of studies will result in greater uncertainty regarding the true estimate for the effective dilution. Consequently, when our samples have minimal sample overlap, we recommend using joint estimation. However, if samples from independent data are unattainable, this approach provides us with a robust tool to estimate effective dilution even in the presence of sample overlap.

### Evaluating hypothesis test, computing confidence intervals and p-values

To compute confidence intervals for the effective dilution, φ, we create 2,000 circular blocked bootstrap samples, and construct empirical 95% confidence intervals for φ. We employ an automated procedure to compute the size of the blocks so that our confidence intervals are robust to the spatial dependence between nearby SNPs^50^. For computing p-values, we considered a combination of three different approaches. From the 2,000 bootstrap samples, in the event we had at least 10 bootstrap samples with effective dilution values greater than 1 and 10 bootstrap samples with effective dilution values less than 1, we could invoke the central limit theorem to estimate the p-value directly from the bootstrap samples. For small p-values, (e.g. p < .01) we considered two additional techniques.

First, we can test if the empirical distribution for the effective dilution from the 2,000 bootstrap samples resembles an approximate normal distribution using the Shapiro-Wilk test, D’Agostino’s K^2^ Test, and the Anderson-Darling Test. Prior to performing the aforementioned tests, if the mean estimated effective dilution parameter exceeded 1 we transformed the effective dilution distribution by taking its reciprocal for convenience, so that the null hypothesis (of no dilution) would always be greater than the observed estimate. Provided that the p-values indicated insufficient evidence to reject the normal distribution assumption for all tests (p > .05), we then used the method of moments to fit a normal distribution to the bootstrapped distribution of the effective dilution and computed a two-sided p-value to ascertain how likely we would observe such an extreme observation for dilution assuming that our null hypothesis of no effective dilution was actually true.

Alternatively, we leverage Extreme Value Theory to fit the tail of our empirical bootstrap distribution to a General Pareto Distribution^51^. We fit the parameters of the General Pareto Distribution using maximum likelihood estimation^52^ and can compute estimated p-values along with confidence intervals for these p-values using a similar approach that has been previously described in the literature^51^. In particular, we computed confidence intervals for our p-values by exploiting the asymptotic bivariate normality of the maximum likelihood estimator. After estimating our covariance matrix, we simulated 10,000 bivariate normal variables to compute expected values for the aforementioned p-values. For the p-values listed in the main table that are significant at the .05 level, we selected the larger p-value of the two methodologies, with the exception that if we had strong evidence that the empirical distribution did not follow a normal distribution, we selected the p-value generated from Extreme Value Theory.

### Meta-analysis

With the known effective dilution weights, we apply the following transformation so that effects across studies share the same mean. That is, if 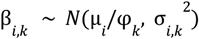, then 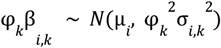, so that the observed i^th^ effect now shares the same mean across all studies k. We can then perform an inverse-variance weights meta-analysis using the observed effect size 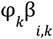 and estimated variance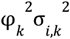. Note that while we estimate φ_*k*_ using only approximately independent loci, we can apply the proposed weighting to all SNPs in the meta-analysis. In the Supplementary Results section, we describe an extension of Dilution-Adjusted Weights, called Transformed Effects Meta-Analysis to infer both linear and non-linear hidden transformations that can bias the summary statistics produced from GWA studies.

When benchmarking the meta-analysis methodologies on use-cases, we ran MTAG using the code from the MTAG GitHub^34^, where we specified options indicating perfect genetic correlation and no sample overlap. We leveraged the MetaFor package for the Random Effects Meta-Analysis^33^. To implement the Weighted Z methodology, we utilized the effective sample size for weighting as recommended in METAL^14^. For assessing the number of significant hits, we treated the extended MHC locus, chr6 25-33 Mbp as one locus. For a theoretical discussion outlining various assumptions of different methodologies, we refer the reader to the Supplement.

### Simulations

To validate our theoretical derivation in **Fig. 2a**, for each simulated GWA study, we aggregated patients into four groups: true positive phenotypes, false positive phenotypes, false negative phenotypes and true negative phenotypes. For each independent SNP, we modeled the total allele counts for each of these four groups as a Binomial random variable (which depends on the number of patients in the group, the true status of the phenotype, and the conditional probability of possessing the effect allele given the true status of the phenotype). Subsequently, we computed the log odds ratio between the labeled phenotypes. For simplicity, within each GWA study we set the number of labeled cases and controls to be equal to each other. We denoted the study with higher markedness as the “gold” study and the study with lower markedness as the “diluted” study. Possible values for different parameters are listed as follows: PPV_gold_ = {0.8,0.9,1}, NPV_gold_ = {0.8,0.9,1}, PPV_diluted_ = {0.6,0.7,0.8,0.9}, NPV_diluted_ = {0.6,0.7,0.8,0.9}, N_controls_,N_cases_ = {20,000}. For each study, we generated 100,000 independent SNPs, of which 10% of the SNPs corresponded to true effects. For both studies, we specified a common heritability parameter that corresponds to the typical effect size of a true effect, h^2^ = {0.025}, which is described in greater detail below. We model the minor allele frequencies for controls, Pr(Minor Allele|True Control) for each SNP using independent identically distributed uniform random variables bounded from 0.05 to 0.5.

We then generate probabilities, Pr(Minor Allele|True Case) for each SNP, and define the *Pr*(*Minor Allele*|*True Case*) = *Pr*(*Minor Allele*|*True Control*) + β_*_ /*H*, where *H* = 1/*Pr*(*Minor Allele*|*True Control*) + 1/(1 − *Pr*(*Minor Allele*|*True Control*)) and β_*_ ∼ *N*(0, *h*^2^ ).

We leveraged a similar approach in **Fig. 2c**, to assess the reduction in variance when jointly analyzing the effective dilution across multiple studies.

For the p-value calibration simulation in **Fig. 2b**, where there is no dilution between the two GWA studies, we generated summary statistics for 100,000 independent SNPs, of which 10% of them correspond to true effects. For SNPs with true effects (log odds ratios), β ∼ *N*(0, 0. 02 ^2^).

Alternatively for null effects β = 0. We then modeled the reported GWA estimates for β_*GWAS*_ and SE such that 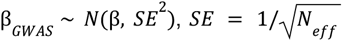,and SE is treated as a known quantity, where *N*_*eff*_ =20,000.

We construct simulated pairs of GWA study summary statistics 2,000 times and for each pair of GWA studies we estimate the effective dilution and compute a p-value by creating 2,000 bootstrap samples on the SNPs in the simulated GWA summary statistics. Specifically we estimate the effective dilution for each bootstrap sample and compute the p-value as,

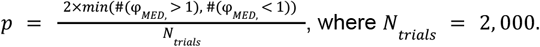

For the meta-analysis simulations, to generate the distribution of effect sizes for SNPs in an undiluted study, 90% of SNPs had a true effect size of 0, whereas for the other 10% of SNPs β ∼ *N*(0, *h*^2^), for some parameter *h*^2^, represented as the inverse of the sample size parameter in the simulation .Since phenotypic misclassification bias is an approximately linear transformation, each study has a dilution factor and true effect sizes for that study were divided by the dilution factor. With the true study-specific effect sizes, we could then simulate the observed effect size and standard error by adding an independent normal random variable with mean 0 and variance equal to the square of the standard error to attain our simulated GWA studies. In simulation, we meta-analyzed 5 GWA studies where each study had an effective sample size N = [500, 2,500] and effective dilution = [1, 1.5, 2]. For simulation, we used a method of moments approach to compute the covariance matrix as detailed in the MTAG supplement. On both simulation and real-world data examples, we set the correlation between different studies to equal 1. In particular for simulation, we utilized the version of MTAG that uses the SNP standard errors (as opposed to sample size) when assigning weights for each study. For simulations, we used a method of moments approach to perform random effects meta-analysis^53^ .

### GWAS Data Sources

For data sources excluding the Schizophrenia (“SCZ AFR”, “SCZ EUR”, “SCZ HIS”) and Bipolar Disorder (e.g. “BD 1 Phecode”, “BD 2+ Phecodes”) GWA studies from the Million Veteran Program (MVP), we leveraged existing GWA summary statistics. Cases as defined from the MVP Schizophrenia GWAS were required to have 2 or more schizophrenia phecodes, 295.1. The Alzheimer’s disease GWA study (“ALZ Traditional”) and the Depression GWA Study (“MDD FinnGen”) come from Release 7 of FinnGen^30^, where we selected the “wide definition” for Alzheimer’s disease based on diagnoses as listed through cause of death, hospital discharge, insurance reimbursement, and medication reimbursement. Controls were excluded for dementia, memory loss, severe traumatic brain injury, neurological diseases, and psychiatric diseases. For details see, https://risteys.finngen.fi/endpoints/G6_AD_WIDE_EXMORE.

GWA summary statistics for the Alzheimer’s disease study leveraging predominantly proxy cases from the UKBiobank (“ALZ Proxy”) are publicly available through the GWAS catalog https://www.ebi.ac.uk/gwas/studies/GCST90027158^29^. For details on the depression phenotype, see https://risteys.finngen.fi/endpoints/F5_DEPRESSIO

Other GWA studies used in this work for Major Depressive Disorder (“MDD PGC”) and Schizophrenia (“SCZ PGC”) are available through the Psychiatric Genomic Consortium^26,54,55^. For the obesity (“Obesity”) and morbid obesity (“Morbid Obesity”) phenotypes, corresponding to PheCodes 278.1 and 278.11, we leveraged a beta release of the gwPheWAS project from August 15th, 2022, which encompasses GWA studies in MVP for over 1,700 phenotypes. In particular to compute the effective dilution between the European ancestry GWA studies for obesity and morbid obesity, as they had overlapping samples, we utilized an African ancestry obesity GWA study from the gwPheWAS project as well. All GWA studies from the gwPheWAS project were adjusted for age, sex, and the first 10 population specific principal components.

To benchmark the meta-analysis methodologies for depression, we leveraged the East Asian ancestry GWAS from the Psychiatric Genomics Consortium^56^ for validation and the FinnGen and European ancestry Psychiatric Genomics Consortium GWAS^57^ for the meta-analysis. For BD, we validated the meta-analysis of our bipolar disorder (BD) GWA studies from the MVP with an existing BD GWAS from the Psychiatric Genomics Consortium^37^. For alcohol dependence, we leveraged Release 7 of FinnGen to attain an Alcohol Dependence GWAS along with European and African ancestry Alcohol Dependence GWA studies came from the Psychiatric Genomic Consortium^58^, where we used the African ancestry GWAS for validation. For details on the FinnGen alcohol dependence phenotype, see https://risteys.finngen.fi/endpoints/F5_ALCOHOL_DEPENDENCE. Finally, we validated the cross-ancestry schizophrenia GWA studies from the MVP with the European ancestry schizophrenia GWAS from the Psychiatric Genomic Consortium^59^. For sample sizes for each of the aforementioned cohorts, see Table S12.

#### Genotyping, quality control and imputation

MVP Genotyping, initial quality control, and imputation is handled by the MVP Data Team as previously described^60,61^. For completeness, we provide a brief overview of the methodologies employed. This study leveraged release 4 of the MVP, which contains genotyping data from blood samples for more than 650,000 individuals. The MVP 1.0 custom Genotyping array was used for genotyping, where the APT software 2.11.3 was used for genotype calling. Genotype quality control metrics encompass: sex check, rare heterozygous adjustment, plate normalization and sample labeling. The MVP Data Team performed ancestry-specific principal component analysis using PLINK 2.0^62^ and computed relatedness (kinship coefficient) with KING 2.1^63^. Ancestry was computed using HARE^64^ (genetically inferred ancestry leveraging the top 30 principal components and ADMIXTURE^65^) and SHAPEIT4^66^ version 4.1.3 was used to phase genotypes. Pre-imputation QC filtered out variants that had significant missingness (> 20%), were monomorphic, or had substantial variation in frequency across batches. The MVP Data Team imputed genotypes with the reference panel from 1,000 Genomes (except for African individuals, where the Sanger Institute African Genome Resources Panel^67^ using Minimac4^68^ was used for imputation). We then performed follow-up quality control filters where we only included samples with sample call rates exceeding 98.5%, required that that the sample heterozygosity rates deviate at most four standard deviations from the samples’ heterozygosity rate mean and filtered out related samples/samples with cryptic relationship by leveraging a cut-off value of .0884 for the kinship coefficient (first we removed samples with multiple relationships; subsequently, we filtered out samples with the highest missingness rates from the remaining relationship pairs). We performed variant-level filtering retaining SNPs such that the MAF exceeded 0.005, the effective minor allele count was at least 30, the variant call rates exceeded 95%, the Hardy-Weinberg equilibrium test p-value exceeded 5 × 10^-8^, and the imputation R^2^ exceeded 0.4. For binary traits, we leveraged logistic regression for performing the GWAS analysis with PLINK 2.0. We performed separate analyses on different genetic ancestries and adjusted for confounders, including sex, age and the top 20 principal components.

### Heritability

We estimated SNP heritability and genetic correlation using the LDSC package^20,69^. Heritability for case-control phenotypes were computed on the liability scale. For Bipolar Disorder, assumed population prevalence was 2% and for Schizophrenia, assumed population prevalence was 1%. To compute the heritability and genetic correlation estimates, the 1k Genomes Project - European Project was used to construct the reference panel.

### TWAS Data Sources and Pipeline

From the MVP, we leveraged individual level imputed gene expression data to perform transcriptome-wide association studies (TWAS) on European ancestry patients with BD 1 Phecode (n_cases_ = 6,408; n_controls_ = 78,626), BD 2+ Phecodes (n_cases_ = 26,560; n_controls_ = 325,892), SCZ 1 Phecode (n_cases_ = 2,110; n_controls_ = 81,405), and SCZ 2 + Phecodes (n_cases_ = 8,924; n_controls_ = 344,293). Genetically regulated gene expression (GReX) was imputed for the dorso-lateral prefrontal cortex (DLPFC, brain) homogenate for each individual by leveraging an EpiXcan^70^ transcriptomic model based on the PsychENCODE cohort^71,72^ trained as previously described^73^.

In order to estimate individually imputed GReX, we first perform QC of MVP genotypes. We utilize the same sample-level QC as described above (see “Genotyping, quality control and imputation”) and filter variants based on MAF and variant imputation (MAF >= 0.01, population specific R^2^ >= 0.6). We also limit variants to biallelic SNPs. Next, we convert genotypes to dosages for use in linear combinations with transcriptomic model gene-SNP weights. We substitute missing genotypes with twice the SNP MAF (representing the population average). This replaces traditional filtering of SNPs by missingness to maximize genotypic information in GReX estimation. To ensure confident GReX estimation, we limit genes to those with a 0.01 or greater prediction performance R^2^. Finally, we perform TWAS with estimated GReX by utilizing a logistic regression (logit link function) with covariates for sex, age, and top ten principal components.

For validation, we used TWAS summary statistics generated from the same transcriptomic imputation model applied to the summary statistics from PGC for BD^38^ and SCZ^74^ with the S-PrediXcan approach^75^. Similar to the GWAS setting, we created randomly assigned p-values for each gene, and clumped lead genes, where we required that all genes in the same clump must have an R^2^ of at least 0.5 with the lead gene and either overlap with the lead gene or be at most 250 kbp away from the lead gene. We excluded patients that had exactly 1 SCZ phecode and 1 BD phecode from the analysis.

## Code Availability

The PheMED GitHub Repo can be downloaded from https://github.com/DiseaseNeuroGenomics/PheMED

## Data Availability

Summary statistics will become available on dbGaP when paper is accepted.

## Ethical Compliance Statement

The authors declare no competing interests. Dr. Harvey reported personal fees from Alkermes, Karuna Pharma, Merck Pharma, Minerva Pharma, and Sunovion Pharma outside the submitted work; and royalties from WCG Verasci outside the submitted work. No other disclosures were reported. The study protocols in the present study, which involved secondary analysis of existing data collected through the primary Million Veteran Program study, were reviewed and approved by the Department of Veterans Affairs Central Institutional Review Board.

## Acknowledgements

This research is based on data from the Million Veteran Program, Office of Research and Development, Veterans Health Administration, and was supported by award #MVP006. This publication does not represent the views of the Department of Veteran Affairs or the United States Government. This study was also supported by the National Institutes of Health (NIH), Bethesda, MD under award numbers K08MH122911 and R01AG078657 (Georgios Voloudakis), R01MH125246, R01AG065582, R01AG067025, R01AG050986, U01MH116442, R01MH109677, and by the Veterans Affairs Merit grants I01BX004189 (Panos Roussos). This study has also been funded in part by the Brain & Behavior Research Foundation via the 2020 NARSAD Young Investigator Grant #29350 (Georgios Voloudakis). We acknowledge the VA Million Veteran Program (MVP) and the VA-DOE genome-wide PheWAS core analytic team for generating the corresponding summary statistics that were used in this manuscript. We want to acknowledge the participants and investigators of the FinnGen study. We acknowledge the Psychiatric Genomics Consortium for the corresponding summary statistics that were used in this manuscript. This work was supported in part through the computational resources and staff expertise provided by Scientific Computing at the Icahn School of Medicine at Mount Sinai.

## Bibliography

1. Beesley, L. J. & Mukherjee, B. Statistical inference for association studies using electronic health records: handling both selection bias and outcome misclassification. Biometrics 78, 214–226 (2022).

2. Akinhanmi, M. O. et al. Racial disparities in bipolar disorder treatment and research: a call to action. Bipolar Disord. 20, 506–514 (2018).

3. Gara, M. A., Minsky, S., Silverstein, S. M., Miskimen, T. & Strakowski, S. M. A naturalistic study of racial disparities in diagnoses at an outpatient behavioral health clinic. Psychiatr. Serv. 70, 130–134 (2019).

4. Nagelkerke, N. & Fidler, V. Estimating a logistic discrimination functions when one of the training samples is subject to misclassification: A maximum likelihood approach. PLoS ONE 10, e0140718 (2015).

5. Zhang, Q. & Yi, G. Y. Genetic association studies with bivariate mixed responses subject to measurement error and misclassification. Stat. Med. 39, 3700–3719 (2020).

6. Rekaya, R., Smith, S., Hay, E. H., Farhat, N. & Aggrey, S. E. Analysis of binary responses with outcome-specific misclassification probability in genome-wide association studies. Appl. Clin. Genet. 9, 169–177 (2016).

7. Gilbert, R. et al. Misclassification of outcome in case-control studies: Methods for sensitivity analysis. Stat. Methods Med. Res. 25, 2377–2393 (2016).

8. Ling, A., Hay, E. H., Aggrey, S. E. & Rekaya, R. A Bayesian approach for analysis of ordered categorical responses subject to misclassification. PLoS ONE 13, e0208433 (2018).

9. Smith, S., Hay, E. H., Farhat, N. & Rekaya, R. Genome wide association studies in presence of misclassified binary responses. BMC Genet. 14, 124 (2013).

10. Tong, J. et al. An augmented estimation procedure for EHR-based association studies accounting for differential misclassification. J. Am. Med. Inform. Assoc. 27, 244–253 (2020).

11. Duffy, S. W. et al. A simple model for potential use with a misclassified binary outcome in epidemiology. J. Epidemiol. Community Health 58, 712–717 (2004).

12. Gao, X., Becker, L. C., Becker, D. M., Starmer, J. D. & Province, M. A. Avoiding the high Bonferroni penalty in genome-wide association studies. Genet. Epidemiol. 34, 100–105 (2010).

13. Beesley, L. J., Fritsche, L. G. & Mukherjee, B. An analytic framework for exploring sampling and observation process biases in genome and phenome-wide association studies using electronic health records. Stat. Med. 39, 1965–1979 (2020).

14. Willer, C. J., Li, Y. & Abecasis, G. R. METAL: fast and efficient meta-analysis of genomewide association scans. Bioinformatics 26, 2190–2191 (2010).

15. Powers, D. Evaluation: From precision, recall and fmeasure to roc, informedness, markedness and correlation. (2007).

16. Speed, D., Kaphle, A. & Balding, D. J. SNP-based heritability and selection analyses: Improved models and new results. Bioessays 44, e2100170 (2022).

17. Zaitlen, N. & Kraft, P. Heritability in the genome-wide association era. Hum. Genet. 131, 1655–1664 (2012).

18. Lynch, M. & Walsh, B. Genetics and analysis of quantitative traits. (Sinauer, 1998).

19. Vattikuti, S., Guo, J. & Chow, C. C. Heritability and genetic correlations explained by common SNPs for metabolic syndrome traits. PLoS Genet. 8, e1002637 (2012).

20. Bulik-Sullivan, B. et al. LD Score regression distinguishes confounding from polygenicity in genome-wide association studies. Nat. Genet. 47, 291–295 (2015).

21. Bigdeli, T. B. et al. Penetrance and pleiotropy of polygenic risk scores for schizophrenia, bipolar disorder, and depression among adults in the US veterans affairs health care system. JAMA Psychiatry 79, 1092–1101 (2022).

22. Bryc, K., Durand, E. Y., Macpherson, J. M., Reich, D. & Mountain, J. L. The genetic ancestry of African Americans, Latinos, and European Americans across the United States. Am. J. Hum. Genet. 96, 37–53 (2015).

23. Yudell, M., Roberts, D., DeSalle, R. & Tishkoff, S. SCIENCE AND SOCIETY. Taking race out of human genetics. Science 351, 564–565 (2016).

24. Vilhjálmsson, B. J. & Nordborg, M. The nature of confounding in genome-wide association studies. Nat. Rev. Genet. 14, 1–2 (2013).

25. Mostafavi, H. et al. Variable prediction accuracy of polygenic scores within an ancestry group. eLife 9, (2020).

26. Howard, D. M. et al. Genome-wide meta-analysis of depression identifies 102 independent variants and highlights the importance of the prefrontal brain regions. Nat. Neurosci. 22, 343–352 (2019).

27. Schizophrenia Working Group of the Psychiatric Genomics Consortium. Biological insights from 108 schizophrenia-associated genetic loci. Nature 511, 421–427 (2014).

28. Lam, M. et al. Comparative genetic architectures of schizophrenia in East Asian and European populations. Nat. Genet. 51, 1670–1678 (2019).

29. Bellenguez, C. et al. New insights into the genetic etiology of Alzheimer’s disease and related dementias. Nat. Genet. 54, 412–436 (2022).

30. Kurki, M. I. et al. FinnGen: Unique genetic insights from combining isolated population and national health register data. medRxiv (2022) doi:10.1101/2022.03.03.22271360.

31. Daetwyler, H. D., Villanueva, B. & Woolliams, J. A. Accuracy of predicting the genetic risk of disease using a genome-wide approach. PLoS ONE 3, e3395 (2008).

32. Liang, Y. et al. Polygenic transcriptome risk scores (PTRS) can improve portability of polygenic risk scores across ancestries. Genome Biol. 23, 23 (2022).

33. Viechtbauer, W. Conducting Meta-Analyses in R with the metafor Package. J. Stat. Softw. 36, (2010).

34. Turley, P. et al. Multi-trait analysis of genome-wide association summary statistics using MTAG. Nat. Genet. 50, 229–237 (2018).

35. Gorman, B. R. et al. Distinctive cross-ancestry genetic architecture for age-related macular degeneration. medRxiv (2022) doi:10.1101/2022.08.16.22278855.

36. Yuan, J. et al. Leveraging correlations between variants in polygenic risk scores to detect heterogeneity in GWAS cohorts. PLoS Genet. 16, e1009015 (2020).

37. Stahl, E. A. et al. Genome-wide association study identifies 30 loci associated with bipolar disorder. Nat. Genet. 51, 793–803 (2019).

38. Mullins, N. et al. Genome-wide association study of more than 40,000 bipolar disorder cases provides new insights into the underlying biology. Nat. Genet. 53, 817–829 (2021).

39. Watson, H. J. et al. Genome-wide association study identifies eight risk loci and implicates metabo-psychiatric origins for anorexia nervosa. Nat. Genet. 51, 1207–1214 (2019).

40. de Vlaming, R. et al. Meta-GWAS Accuracy and Power (MetaGAP) Calculator Shows that Hiding Heritability Is Partially Due to Imperfect Genetic Correlations across Studies. PLoS Genet. 13, e1006495 (2017).

41. Manchia, M. et al. The impact of phenotypic and genetic heterogeneity on results of genome wide association studies of complex diseases. PLoS ONE 8, e76295 (2013).

42. Edwards, B. J., Haynes, C., Levenstien, M. A., Finch, S. J. & Gordon, D. Power and sample size calculations in the presence of phenotype errors for case/control genetic association studies. BMC Genet. 6, 18 (2005).

43. Verma, S. & Rubin, J. Fairness definitions explained. in Proceedings of the International Workshop on Software Fairness 1–7 (ACM, 2018). doi:10.1145/3194770.3194776.

44. Mehrabi, N., Morstatter, F., Saxena, N., Lerman, K. & Galstyan, A. A survey on bias and fairness in machine learning. ACM Comput. Surv. 54, 1–35 (2021).

45. Dueñas, H. R., Seah, C., Johnson, J. S. & Huckins, L. M. Implicit bias of encoded variables: frameworks for addressing structured bias in EHR-GWAS data. Hum. Mol. Genet. 29, R33–R41 (2020).

46. Panch, T., Mattie, H. & Atun, R. Artificial intelligence and algorithmic bias: implications for health systems. J. Glob. Health 9, 010318 (2019).

47. Virtanen, P. et al. SciPy 1.0: fundamental algorithms for scientific computing in Python. Nat. Methods 17, 261–272 (2020).

48. Purcell, S. et al. PLINK: a tool set for whole-genome association and population-based linkage analyses. Am. J. Hum. Genet. 81, 559–575 (2007).

49. 1000 Genomes Project Consortium et al. An integrated map of genetic variation from 1,092 human genomes. Nature 491, 56–65 (2012).

50. Politis, D. N. & White, H. Automatic Block-Length Selection for the Dependent Bootstrap. Econom. Rev. 23, 53–70 (2004).

51. Knijnenburg, T. A., Wessels, L. F. A., Reinders, M. J. T. & Shmulevich, I. Fewer permutations, more accurate P-values. Bioinformatics 25, i161–8 (2009).

52. Grimshaw, S. D. Computing maximum likelihood estimates for the generalized pareto distribution. Technometrics 35, 185 (1993).

53. Higgins, J. P. T., Thompson, S. G. & Spiegelhalter, D. J. A re-evaluation of random-effects meta-analysis. J R Stat Soc Ser A Stat Soc 172, 137–159 (2009).

54. Bipolar Disorder and Schizophrenia Working Group of the Psychiatric Genomics Consortium. Electronic address: & Bipolar Disorder and Schizophrenia Working Group of the Psychiatric Genomics Consortium. Genomic dissection of bipolar disorder and schizophrenia, including 28 subphenotypes. Cell 173, 1705-1715.e16 (2018).

55. Lam, M. et al. RICOPILI: rapid imputation for consortias pipeline. Bioinformatics 36, 930–933 (2020).

56. Giannakopoulou, O. et al. The Genetic Architecture of Depression in Individuals of East Asian Ancestry: A Genome-Wide Association Study. JAMA Psychiatry 78, 1258–1269 (2021).

57. Lawrence, N. S. et al. Subcortical and ventral prefrontal cortical neural responses to facial expressions distinguish patients with bipolar disorder and major depression. Biol. Psychiatry 55, 578–587 (2004).

58. Walters, R. K. et al. Transancestral GWAS of alcohol dependence reveals common genetic underpinnings with psychiatric disorders. Nat. Neurosci. 21, 1656–1669 (2018).

59. Pardiñas, A. F. et al. Common schizophrenia alleles are enriched in mutation-intolerant genes and in regions under strong background selection. Nat. Genet. 50, 381–389 (2018).

60. Hunter-Zinck, H. et al. Genotyping array design and data quality control in the million veteran program. Am. J. Hum. Genet. 106, 535–548 (2020).

61. Kang, J. et al. Genome-wide association study of treatment resistant depression highlights shared biology with metabolic traits. medRxiv (2022) doi:10.1101/2022.08.10.22278630.

62. Chang, C. C. et al. Second-generation PLINK: rising to the challenge of larger and richer datasets. Gigascience 4, 7 (2015).

63. Manichaikul, A. et al. Robust relationship inference in genome-wide association studies. Bioinformatics 26, 2867–2873 (2010).

64. Fang, H. et al. Harmonizing Genetic Ancestry and Self-identified Race/Ethnicity in Genome-wide Association Studies. Am. J. Hum. Genet. 105, 763–772 (2019).

65. Alexander, D. H., Novembre, J. & Lange, K. Fast model-based estimation of ancestry in unrelated individuals. Genome Res. 19, 1655–1664 (2009).

66. Delaneau, O., Zagury, J.-F., Robinson, M. R., Marchini, J. L. & Dermitzakis, E. T. Accurate, scalable and integrative haplotype estimation. Nat. Commun. 10, 5436 (2019).

67. Gurdasani, D. et al. The African Genome Variation Project shapes medical genetics in Africa. Nature 517, 327–332 (2015).

68. Das, S. et al. Next-generation genotype imputation service and methods. Nat. Genet. 48, 1284–1287 (2016).

69. Bulik-Sullivan, B. et al. An atlas of genetic correlations across human diseases and traits. Nat. Genet. 47, 1236–1241 (2015).

70. Zhang, W. et al. Integrative transcriptome imputation reveals tissue-specific and shared biological mechanisms mediating susceptibility to complex traits. Nat. Commun. 10, 3834 (2019).

71. Wang, D. et al. Comprehensive functional genomic resource and integrative model for the human brain. Science 362, (2018).

72. Gandal, M. J. et al. Transcriptome-wide isoform-level dysregulation in ASD, schizophrenia, and bipolar disorder. Science 362, (2018).

73. Fullard, J. F. et al. Single-nucleus transcriptome analysis of human brain immune response in patients with severe COVID-19. Genome Med. 13, 118 (2021).

74. Trubetskoy, V. et al. Mapping genomic loci implicates genes and synaptic biology in schizophrenia. Nature 604, 502–508 (2022).

75. Barbeira, A. N. et al. Exploring the phenotypic consequences of tissue specific gene expression variation inferred from GWAS summary statistics. Nat. Commun. 9, 1825 (2018).

