## Supplementary Material for "Detecting and Adjusting for Hidden Biases due to Phenotype Misclassification in Genome-Wide Association Studies"

### Supplementary Appendix


**VA Million Veteran Program Core Acknowledgement** **1**
**Supplementary Tables** **7**
**Supplementary Figures** **20**
**Supplementary Results** **34**
Phenotypic misclassification shrinks our means, an overview 34
Mathematical derivation that phenotype misclassification shrinks the means. 35
Supplementary Derivation on Dilution Adjusted Effective Sample Size 39
Joint Estimate of Effective Dilution Increases Power 40
Derivation for Estimating Power with Dilution 40
Derivation Illustrating how Dilution Affects Performance Metrics in Polygenic Risk Score Validation 41
Overview of Transformed Effects Meta-Analysis (TEMA), an Extension of Dilution Weights Meta-Analysis 42
Comparison of DAW with other meta-analysis methods 43
**Supplementary Methods** **45**
Multi-trait dilution simulation 45
Impact of ascertainment bias/sampling prevalence on dilution, simulation 46
Measuring impact of population stratification on dilution with schizophrenia GWAS's 46
Measuring impact of population stratification on simulated traits with real-world genomic data 46
Measuring impact of cryptic relatedness on simulated traits with real-world genomic data 46
Assessing performance of MTAG and PheMED/dilution adjusted weights meta-analysis on real-world genomic data 47



#### VA Million Veteran Program Core Acknowledgement

MVP Program Office

- Program Director - Sumitra Muralidhar, Ph.D.
US Department of Veterans Affairs, 810 Vermont Avenue NW, Washington, DC 20420
- Associate Director, Scientific Programs - Jennifer Moser, Ph.D.
US Department of Veterans Affairs, 810 Vermont Avenue NW, Washington, DC 20420
- Associate Director, Cohort Management & Public Relations - Jennifer E. Deen, B.S.
US Department of Veterans Affairs, 810 Vermont Avenue NW, Washington, DC 20420

MVP Executive Committee

- Co-Chair: J. Michael Gaziano, M.D., M.P.H.

VA Boston Healthcare System, 150 S. Huntington Avenue, Boston, MA 02130
- Co-Chair: Sumitra Muralidhar, Ph.D.
US Department of Veterans Affairs, 810 Vermont Avenue NW, Washington, DC 20420
- Jean Beckham, Ph.D.
Durham VA Medical Center, 508 Fulton Street, Durham, NC 27705
- Kyong-Mi Chang, M.D.
Philadelphia VA Medical Center, 3900 Woodland Avenue, Philadelphia, PA 19104
- Philip S. Tsao, Ph.D.
VA Palo Alto Health Care System, 3801 Miranda Avenue, Palo Alto, CA 94304
- Shiuh-Wen Luoh, M.D., Ph.D.
VA Portland Health Care System, 3710 SW US Veterans Hospital Rd, Portland, OR 97239
US Department of Veterans Affairs, 810 Vermont Avenue NW, Washington, DC 20420
- Juan P. Casas, M.D., Ph.D., Ex-Officio
VA Boston Healthcare System, 150 S. Huntington Avenue, Boston, MA 02130

MVP Principal Investigators

- J. Michael Gaziano, M.D., M.P.H.
VA Boston Healthcare System, 150 S. Huntington Avenue, Boston, MA 02130
- Philip S. Tsao, Ph.D.
VA Palo Alto Health Care System, 3801 Miranda Avenue, Palo Alto, CA 94304

MVP Operations

- MVP Executive Director – Juan P. Casas, M.D., Ph.D.
VA Boston Healthcare System, 150 S. Huntington Avenue, Boston, MA 02130
- Director of Regulatory Affairs – Lori Churby, B.S.
VA Palo Alto Health Care System, 3801 Miranda Avenue, Palo Alto, CA 94304
- MVP Cohort Management Director – Stacey B. Whitbourne, Ph.D.
VA Boston Healthcare System, 150 S. Huntington Avenue, Boston, MA 02130
- MVP Recruitment/Enrollment Director - Jessica V. Brewer, M.P.H.
VA Boston Healthcare System, 150 S. Huntington Avenue, Boston, MA 02130
- Director, VA Central Biorepository, Boston – Mary T. Brophy M.D., M.P.H.
VA Boston Healthcare System, 150 S. Huntington Avenue, Boston, MA 02130
- Executive Director for MVP Biorepositories - Luis E. Selva, Ph.D.
VA Boston Healthcare System, 150 S. Huntington Avenue, Boston, MA 02130
- MVP Informatics, Boston – Shahpoor (Alex) Shayan, M.S.
VA Boston Healthcare System, 150 S. Huntington Avenue, Boston, MA 02130
- Director, MVP Data Operations/Analytics, Boston – Kelly Cho, M.P.H., Ph.D.
VA Boston Healthcare System, 150 S. Huntington Avenue, Boston, MA 02130
- Director, Center for Computational and Data Science (C-DACS) & Genomics Core – Saiju Pyarajan Ph.D.
VA Boston Healthcare System, 150 S. Huntington Avenue, Boston, MA 02130

- Director, Molecular Data Core – Philip S. Tsao, Ph.D.
VA Palo Alto Health Care System, 3801 Miranda Avenue, Palo Alto, CA 94304
- Director, Phenomics Data Core – Kelly Cho, M.P.H, Ph.D.
VA Boston Healthcare System, 150 S. Huntington Avenue, Boston, MA 02130
- Director, VA Informatics and Computing Infrastructure (VINCI) – Scott L. DuVall, Ph.D.
VA Salt Lake City Health Care System, 500 Foothill Drive, Salt Lake City, UT 84148
- MVP Coordinating Centers
    - o Cooperative Studies Program Clinical Research Pharmacy Coordinating Center, Albuquerque – Todd Connor, Pharm.D.; Dean P. Argyres, B.S., M.S.
    New Mexico VA Health Care System, 1501 San Pedro Drive SE, Albuquerque, NM 87108
    - o Genomics Coordinating Center, Palo Alto – Philip S. Tsao, Ph.D.
    VA Palo Alto Health Care System, 3801 Miranda Avenue, Palo Alto, CA 94304
    - o MVP Boston Coordinating Center, Boston - J. Michael Gaziano, M.D., M.P.H.
    VA Boston Healthcare System, 150 S. Huntington Avenue, Boston, MA 02130
    - o MVP Information Center, Canandaigua – Brady Stephens, M.S.
    Canandaigua VA Medical Center, 400 Fort Hill Avenue, Canandaigua, NY 14424

Current MVP Local Site Investigators

- Atlanta VA Medical Center (Peter Wilson, M.D.)
1670 Clairmont Road, Decatur, GA 30033

- Bay Pines VA Healthcare System (Rachel McArdle, Ph.D.)
10,000 Bay Pines Blvd Bay Pines, FL 33744
- Birmingham VA Medical Center (Louis Dellitalia, M.D.)
700 S. 19th Street, Birmingham AL 35233
- Central Western Massachusetts Healthcare System (Kristin Mattocks, Ph.D., M.P.H.)
421 North Main Street, Leeds, MA 01053
- Cincinnati VA Medical Center (John Harley, M.D., Ph.D.)
3200 Vine Street, Cincinnati, OH 45220
- Clement J. Zablocki VA Medical Center (Jeffrey Whittle, M.D., M.P.H.)
5000 West National Avenue, Milwaukee, WI 53295
- VA Northeast Ohio Healthcare System (Frank Jacono, M.D.)
10701 East Boulevard, Cleveland, OH 44106
- Durham VA Medical Center (Jean Beckham, Ph.D.)
508 Fulton Street, Durham, NC 27705
- Edith Nourse Rogers Memorial Veterans Hospital (John Wells., Ph.D.)
200 Springs Road, Bedford, MA 01730
- Edward Hines, Jr. VA Medical Center (Salvador Gutierrez, M.D.)
5000 South 5th Avenue, Hines, IL 60141
- Veterans Health Care System of the Ozarks (Kathrina Alexander, M.D.)

1100 North College Avenue, Fayetteville, AR 72703
- Fargo VA Health Care System (Kimberly Hammer, Ph.D.)
2101 N. Elm, Fargo, ND 58102
- VA Health Care Upstate New York (James Norton, Ph.D.)
113 Holland Avenue, Albany, NY 12208
- New Mexico VA Health Care System (Gerardo Villareal, M.D.)
1501 San Pedro Drive, S.E. Albuquerque, NM 87108
- VA Boston Healthcare System (Scott Kinlay, M.B.B.S., Ph.D.)
150 S. Huntington Avenue, Boston, MA 02130
- VA Western New York Healthcare System (Junzhe Xu, M.D.)
3495 Bailey Avenue, Buffalo, NY 14215-1199
- Ralph H. Johnson VA Medical Center (Mark Hamner, M.D.)
109 Bee Street, Mental Health Research, Charleston, SC 29401
- Columbia VA Health Care System (Roy Mathew, M.D.)
6439 Garners Ferry Road, Columbia, SC 29209
- VA North Texas Health Care System (Sujata Bhushan, M.D.)
4500 S. Lancaster Road, Dallas, TX 75216
- Hampton VA Medical Center (Pran Iruvanti, D.O., Ph.D.)
100 Emancipation Drive, Hampton, VA 23667
- Richmond VA Medical Center (Michael Godschalk, M.D.)
1201 Broad Rock Blvd., Richmond, VA 23249
- Iowa City VA Health Care System (Zuhair Ballas, M.D.)
601 Highway 6 West, Iowa City, IA 52246-2208

- Eastern Oklahoma VA Health Care System (River Smith, Ph.D.)
1011 Honor Heights Drive, Muskogee, OK 74401
- James A. Haley Veterans' Hospital (Stephen Mastorides, M.D.)
13000 Bruce B. Downs Blvd, Tampa, FL 33612
- James H. Quillen VA Medical Center (Jonathan Moorman, M.D., Ph.D.)
Corner of Lamont & Veterans Way, Mountain Home, TN 37684
- John D. Dingell VA Medical Center (Saib Gappy, M.D.)
4646 John R Street, Detroit, MI 48201
- Louisville VA Medical Center (Jon Klein, M.D., Ph.D.)
800 Zorn Avenue, Louisville, KY 40206
- Manchester VA Medical Center (Nora Ratcliffe, M.D.)
718 Smyth Road, Manchester, NH 03104
- Miami VA Health Care System (Ana Palacio, M.D., M.P.H.)
1201 NW 16th Street, 11 GRC, Miami FL 33125
- Michael E. DeBakey VA Medical Center (Olaoluwa Okusaga, M.D.)
2002 Holcombe Blvd, Houston, TX 77030
- Minneapolis VA Health Care System (Maureen Murdoch, M.D., M.P.H.)
One Veterans Drive, Minneapolis, MN 55417
- N. FL/S. GA Veterans Health System (Peruvemba Sriram, M.D.)
1601 SW Archer Road, Gainesville, FL 32608

- Northport VA Medical Center (Shing Shing Yeh, Ph.D., M.D.)
79 Middleville Road, Northport, NY 11768
- Overton Brooks VA Medical Center (Neeraj Tandon, M.D.)
510 East Stoner Ave, Shreveport, LA 71101
- Philadelphia VA Medical Center (Darshana Jhala, M.D.)
3900 Woodland Avenue, Philadelphia, PA 19104
- Phoenix VA Health Care System (Samuel Aguayo, M.D.)
650 E. Indian School Road, Phoenix, AZ 85012
- Portland VA Medical Center (David Cohen, M.D.)
3710 SW U.S. Veterans Hospital Road, Portland, OR 97239
- Providence VA Medical Center (Satish Sharma, M.D.)
830 Chalkstone Avenue, Providence, RI 02908
- Richard Roudebush VA Medical Center (Suthat Liangpunsakul, M.D., M.P.H.)
1481 West 10th Street, Indianapolis, IN 46202
- Salem VA Medical Center (Kris Ann Oursler, M.D.)
1970 Roanoke Blvd, Salem, VA 24153
- San Francisco VA Health Care System (Mary Whooley, M.D.)
4150 Clement Street, San Francisco, CA 94121
- South Texas Veterans Health Care System (Sunil Ahuja, M.D.)
7400 Merton Minter Boulevard, San Antonio, TX 78229
- Southeast Louisiana Veterans Health Care System (Joseph Constans, Ph.D.)
2400 Canal Street, New Orleans, LA 70119
- Southern Arizona VA Health Care System (Paul Meyer, M.D., Ph.D.)
3601 S 6th Avenue, Tucson, AZ 85723
- Sioux Falls VA Health Care System (Jennifer Greco, M.D.)
2501 W 22nd Street, Sioux Falls, SD 57105
- St. Louis VA Health Care System (Michael Rauchman, M.D.)
915 North Grand Blvd, St. Louis, MO 63106
- Syracuse VA Medical Center (Richard Servatius, Ph.D.)
800 Irving Avenue, Syracuse, NY 13210
- VA Eastern Kansas Health Care System (Melinda Gaddy, Ph.D.)
4101 S 4th Street Trafficway, Leavenworth, KS 66048
- VA Greater Los Angeles Health Care System (Agnes Wallbom, M.D., M.S.)
11301 Wilshire Blvd, Los Angeles, CA 90073
- VA Long Beach Healthcare System (Timothy Morgan, M.D.)
5901 East 7th Street Long Beach, CA 90822
- VA Maine Healthcare System (Todd Stapley, D.O.)
1 VA Center, Augusta, ME 04330
- VA New York Harbor Healthcare System (Peter Liang, M.D., M.P.H.)
423 East 23rd Street, New York, NY 10010
- VA Pacific Islands Health Care System (Daryl Fujii, Ph.D.)
459 Patterson Rd, Honolulu, HI 96819
- VA Palo Alto Health Care System (Philip Tsao, Ph.D.)
3801 Miranda Avenue, Palo Alto, CA 94304-1290

- VA Pittsburgh Health Care System (Patrick Strollo, Jr., M.D.)
University Drive, Pittsburgh, PA 15240
- VA Puget Sound Health Care System (Edward Boyko, M.D.)
1660 S. Columbian Way, Seattle, WA 98108-1597
- VA Salt Lake City Health Care System (Jessica Walsh, M.D.)
500 Foothill Drive, Salt Lake City, UT 84148
- VA San Diego Healthcare System (Samir Gupta, M.D., M.S.C.S.)
3350 La Jolla Village Drive, San Diego, CA 92161
- VA Sierra Nevada Health Care System (Mostaqul Huq, Pharm.D., Ph.D.)
975 Kirman Avenue, Reno, NV 89502
- VA Southern Nevada Healthcare System (Joseph Fayad, M.D.)
6900 North Pecos Road, North Las Vegas, NV 89086
- VA Tennessee Valley Healthcare System (Adriana Hung, M.D., M.P.H.)
1310 24th Avenue, South Nashville, TN 37212
- Washington DC VA Medical Center (Jack Lichy, M.D., Ph.D.)
50 Irving St, Washington, D. C. 20422
- W.G. (Bill) Hefner VA Medical Center (Robin Hurley, M.D.)
1601 Brenner Ave, Salisbury, NC 28144
- White River Junction VA Medical Center (Brooks Robey, M.D.)
163 Veterans Drive, White River Junction, VT 05009
- William S. Middleton Memorial Veterans Hospital (Prakash Balasubramanian, M.D.)
2500 Overlook Terrace, Madison, WI 53705

#### Supplementary Tables

| Simulation | PPV Study 1 | PPV Study 2 | PPV Extra Study | NPV for All Studies | Sample Size for All Studies (Number of cases = Number of controls) |
|---|---|---|---|---|---|
| Scenario 1 | 1 | 0.7 | 0.9 | 1 | 20,000 |
| Scenario 2 | 1 | 0.7 | 0.7 | 1 | 20,000 |
| Scenario 3 | 1 | 0.8 | 0.8 | 1 | 20,000 |
| Scenario 4 | 1 | 0.9 | 0.9 | 1 | 20,000 |
| Scenario 5 | 1 | 0.9 | 0.7 | 1 | 20,000 |

**Table S1 | Simulation Scenarios for Fig 2c.** We provide the PPV, NPV and sample sizes used to generate the GWA studies in each of the five simulation scenarios. In Fig 2c we plot the confidence intervals for the effective dilution between Study 2 and Study 1 (which is used as the reference).

| Number of PCs | Dilution against Independent Sample | Dilution P-value | Difference in Dilution from Using 20 PCs |
|---|---|---|---|
| 5 | 1.030 | 0.648 | +0.006 |
| 10 | 1.025 | 0.700 | +0.001 |
| 15 | 1.027 | 0.682 | +0.003 |
| 20 | 1.024 | 0.716 | N/A |

**Table S2 | Assessing impact of population stratification on dilution between two schizophrenia GWAS's.** We measure the dilution when comparing two schizophrenia GWAS's from the Million Veteran Program, where we randomly allocate cases and controls to a different group and perform GWAS's on the different groups. For one of the groups, we adjust for a fixed number of PC's (20). For the other group, we vary the number of PC's that we adjust (n = 5, 10, 15, 20).

| Trait 1 | Trait 2 (reference) | PheMED | | | LDSC | | |
|---|---|---|---|---|---|---|---|
| | | Effective Dilution ($\varphi_{MED}$) | 95% CI | P-value | $r_g$ | SE | P-value |
| BD 1 Phecode | BD 2+ Phecodes | **1.52** | (1.33,1.75) | **$1.35\times10^{-13}$** | 1.329 | 0.855 | $6.00\times10^{-2}$ ($r_g$ out of bounds) |
| Obesity | Morbid Obesity | **1.16** | (1.09,1.24) | **$7.30\times10^{-6}$** | 0.9742 | 0.009 | $< 10^{-300}$ |
| SCZ MVP AFR | SCZ MVP EUR | **2.41** | (1.59,3.83) | **$1.59\times10^{-9}$** | N/A | N/A | N/A |
| SCZ MVP HIS | SCZ MVP EUR | 1.07 | (0.83,1.42) | 0.63 | N/A | N/A | N/A |
| MDD PGC | MDD FinnGen | **1.33** | (1.25,1.43) | **$1.34\times10^{-21}$** | 0.8016 | 0.028 | $2.89\times10^{-186}$ |
| SCZ MVP | SCZ PGC | **1.69** | (1.61,1.82) | **$1.05\times10^{-121}$** | 0.9566 | 0.049 | $1.55\times10^{-85}$ |
| ALZ Proxy Cases UKBB | ALZ Traditional Cases FinnGen | **1.27** | (1.04,1.53) | **$2.80\times10^{-2}$** | 0.6814 | 0.110 | $6.65\times10^{-10}$ |

**Table S3 | Ascertaining Statistical Significance of Effective Dilution and comparison with genetic correlation.** We generate 2,000 circular blocked bootstrap samples to construct 95% CI for the true value for the effective dilution. Statistically significant effective dilution values are highlighted in bold.

| Trait 1 | Trait 2 | Effective Dilution | P-Value Bootstrap Extreme Value Theory Mean P-Value (10k Simulations) | P-Value Bootstrap Normal Approximation | Passed Normality Checks |
|---|---|---|---|---|---|
| SCZ MVP | SCZ PGC | 1.69 | 0 | $1.05\times10^{-121}$ | Yes |
| BD 1 Phecode | BD 2 Phecodes | 1.52 | $2.49\times10^{-14}$ | $1.35\times10^{-13}$ | Yes |
| Obesity | Morbid Obesity | 1.16 | $1.24\times10^{-16}$ | $7.30\times10^{-6}$ | Yes |
| SCZ MVP AFR | SCZ MVP EUR | 2.41 | $1.59\times10^{-9}$ | $3.82\times10^{-10}$ | No |
| MDD PGC | MDD FinnGen | 1.33 | 0 | $1.34\times10^{-21}$ | Yes |

**Table S4 | P-values for different methodologies where estimated p-value < 0.1.** Here, we list the outputs for computing p-values across different use-cases, when the estimated p-value is < 0.1.

| Study 1 | PMID | $h^2_{SNP}$ | 95% CI | Study 2 | PMID | $h^2_{SNP}$ | 95% CI |
|---|---|---|---|---|---|---|---|
| SCZ EUR [1] (CLOZUK) [2] | 31740837<br>29483656 | 0.24<br>0.29 | 0.20-0.28<br>0.27-0.31 | SCZ AFR [3] | 33169155 | 0.09 | 0.00-0.19 |
| AN 2018 [4] | 28494655 | 0.20 | 0.16-0.24 | AN 2020 [5] | 31308545 | 0.11 | 0.09-0.13 |
| BD PGC1 [6] | 34002096 | 0.23 | 0.18-0.28 | BD External [6]I | 34002096 | 0.10 | 0.07-0.13 |

**Table S5 | Historic challenges in estimating trait SNP heritability.** We present confidence intervals for heritability for three different phenotypes (Schizophrenia, Anorexia and Bipolar Disorder) as reported in the literature. In particular, we observe that confidence intervals for heritability do not overlap and are inconsistent. As illustrated in Fig 5, meta-analyzing many GWA studies, corresponding to diluted phenotypes, can result in smaller heritability estimates. Perhaps unsurprisingly, more recent studies, which generally leverage more patients and incorporate more inclusive phenotypic definitions, yield smaller heritability estimates compared to their predecessors.

| Definition of Significant Hit | Discovery Cohort Significance | Validation Cohort Significance | Table |
|---|---|---|---|
| Metric 1 | Bonferroni Significant | Not validated | S7 |
| Metric 2 | Bonferroni Significant | FDR Significant | S8 |
| Metric 3 | Bonferroni Significant | Nominal Significance | S9 |
| Metric 4 | FDR Significant | Not validated | S10 |
| Metric 5 | FDR Significant | FDR Significant | S11 |
| Metric 6 | FDR Significant | Nominal Significance | S12 |

**Table S6 | Overview of Tables S6-S11.** Describes the metrics used in Tables S5-S10 to benchmark the meta-analysis methodologies

| Technique/Study | Alcohol Dependence | Bipolar Disorder | Major Depressive Disorder | Schizophrenia Cross-Ancestry | Schizophrenia Cross-Cohort |
|---|---|---|---|---|---|
| DAW | 2 | 1 | 76 | 2 | 140 |
| IVW | 1 | 0 | 81 | 1 | 133 |
| MTAG | 2 | 0 | 72 | NA | 105 |
| REMA | 1 | 0 | 43 | 0 | 74 |
| Weighted Z | 1 | 0 | 81 | 2 | 135 |

**Table S7 | Number of significant hits for different meta-analysis methodologies, Metric 1.** Rows indicate the corresponding meta-analysis methodology used, columns correspond to the use-case and each cell identifies the hits from that meta-analysis according to Metric 1, where hits need to be Bonferroni significant on the discovery cohort. See Table S5 for all metric definitions.

| Technique/Study | Alcohol Dependence | Bipolar Disorder | Major Depressive Disorder | Schizophrenia Cross-Ancestry | Schizophrenia Cross-Cohort |
|---|---|---|---|---|---|
| DAW | 1 | 1 | 1 | 2 | 40 |
| IVW | 1 | 0 | 1 | 1 | 38 |
| MTAG | 1 | 0 | 0 | NA | 31 |
| REMA | 1 | 0 | 1 | 0 | 24 |
| Weighted Z | 1 | 0 | 1 | 1 | 38 |

**Table S8 | Number of validated hits for different meta-analysis methodologies, Metric 2.** Rows indicate the corresponding meta-analysis methodology used, columns correspond to the use-case and each cell identifies the hits from that meta-analysis according to Metric 2, where hits need to be Bonferroni significant on the discovery cohort and FDR significant on the validation cohort. See Table S5 for all metric definitions.

| Technique/Study | Alcohol Dependence | Bipolar Disorder | Major Depressive Disorder | Schizophrenia Cross-Ancestry | Schizophrenia Cross-Cohort |
|---|---|---|---|---|---|
| DAW | 1 | 1 | 12 | 2 | 55 |
| IVW | 1 | 0 | 12 | 1 | 50 |
| MTAG | 1 | 0 | 10 | NA | 42 |
| REMA | 1 | 0 | 7 | 0 | 31 |
| Weighted Z | 1 | 0 | 12 | 1 | 50 |

**Table S9 | Number of significant hits for different meta-analysis methodologies, Metric 3.** Rows indicate the corresponding meta-analysis methodology used, columns correspond to the use-case and each cell identifies the hits from that meta-analysis according to Metric 3, where hits need to be Bonferroni significant on the discovery cohort and nominally significant on the validation cohort. See Table S5 for all metric definitions.

| Technique/Study | Alcohol Dependence | Bipolar Disorder | Major Depressive Disorder | Schizophrenia Cross-Ancestry | Schizophrenia Cross-Cohort |
|---|---|---|---|---|---|
| DAW | 15 | 3 | 765 | 5 | 1,356 |
| IVW | 12 | 2 | 746 | 3 | 1,208 |
| MTAG | 5 | 0 | 572 | NA | 854 |
| REMA | 7 | 0 | 334 | 0 | 538 |
| Weighted Z | 7 | 2 | 730 | 2 | 1,239 |

**Table S10 | Number of significant hits for different meta-analysis methodologies, Metric 4.** Rows indicate the corresponding meta-analysis methodology used, columns correspond to the use-case and each cell identifies the hits from that meta-analysis according to Metric 4, where hits need to be FDR significant on the discovery cohort. See Table S5 for all metric definitions.

| Technique/Study | Alcohol Dependence | Bipolar Disorder | Major Depressive Disorder | Schizophrenia Cross-Ancestry | Schizophrenia Cross-Cohort |
|---|---|---|---|---|---|
| DAW | 0 | 1 | 1 | 5 | 93 |
| IVW | 0 | 1 | 0 | 2 | 80 |
| MTAG | 1 | 0 | 0 | NA | 81 |
| REMA | 0 | 0 | 0 | 0 | 47 |
| Weighted Z | 0 | 1 | 0 | 1 | 81 |

**Table S11 | Number of significant hits for different meta-analysis methodologies, Metric 5.** Rows indicate the corresponding meta-analysis methodology used, columns correspond to the use-case and each cell identifies the hits from that meta-analysis according to Metric 5, where hits need to be FDR significant on both the discovery and validation cohort. See Table S5 for all metric definitions.

| Technique/Study | Alcohol Dependence | Bipolar Disorder | Major Depressive Disorder | Schizophrenia Cross-Ancestry | Schizophrenia Cross-Cohort |
|---|---|---|---|---|---|
| DAW | 3 | 1 | 54 | 5 | 257 |
| IVW | 2 | 1 | 53 | 2 | 226 |
| MTAG | 1 | 0 | 43 | NA | 169 |
| REMA | 2 | 0 | 30 | 0 | 109 |
| Weighted Z | 2 | 1 | 53 | 1 | 232 |

**Table S12 | Number of significant hits for different meta-analysis methodologies, Metric 6.** Rows indicate the corresponding meta-analysis methodology used, columns correspond to the use-case and each cell identifies the hits from that meta-analysis according to Metric 6, where hits need to be FDR significant on the discovery cohort and nominally significant on the validation cohort. See Table S5 for all metric definitions.

| Trait | Source | Cases | Controls | Ancestry |
|---|---|---|---|---|
| Alcohol Dependence | FinnGen | 8,137 | 290,126 | EUR |
| Alzheimer's Disease | FinnGen | 9,271 | 150,169 | EUR |
| Major Depressive Disorder | FinnGen | 33,812 | 271,380 | EUR |
| Schizophrenia | PGC | 33,640 | 43,456 | EUR |
| Major Depressive Disorder | PGC | 170,756 | 329,443 | EUR |
| Major Depressive Disorder | PGC | 13,893 | 85,914 | EAS |
| Alcohol Dependence | PGC | 8,485 | 20,272 | EUR |
| Bipolar Disorder | PGC | 20,352 | 31,358 | EUR |
| Alcohol Dependence | PGC | 2,991 | 2,808 | AFR |
| Alzheimer's Disease | Bellenguez | 85,934 | 401,577 | EUR |
| Obesity | gwPheWAS (MVP) | 172,191 | 248,150 | EUR |
| Morbid Obesity | gwPheWAS (MVP) | 46,629 | 391,905 | EUR |
| Obesity | gwPheWAS (MVP) | 52,255 | 58,643 | AFR |
| Bipolar Disorder (1 Phecode) | MVP | 6,408 | 78,626 | EUR |
| Bipolar Disorder (2+ Phecodes) | MVP | 26,560 | 325,892 | EUR |
| Schizophrenia | MVP | 8,924 | 425,698 | EUR |
| Schizophrenia | MVP | 6,778 | 103,738 | AFR |
| Schizophrenia | MVP | 1,657 | 49,657 | HIS |

**Table S13 | Sample sizes for different GWA Studies.** Each row indicates the sample size (number of cases and controls) for the different GWA studies used in this work.

#### Supplementary Figures

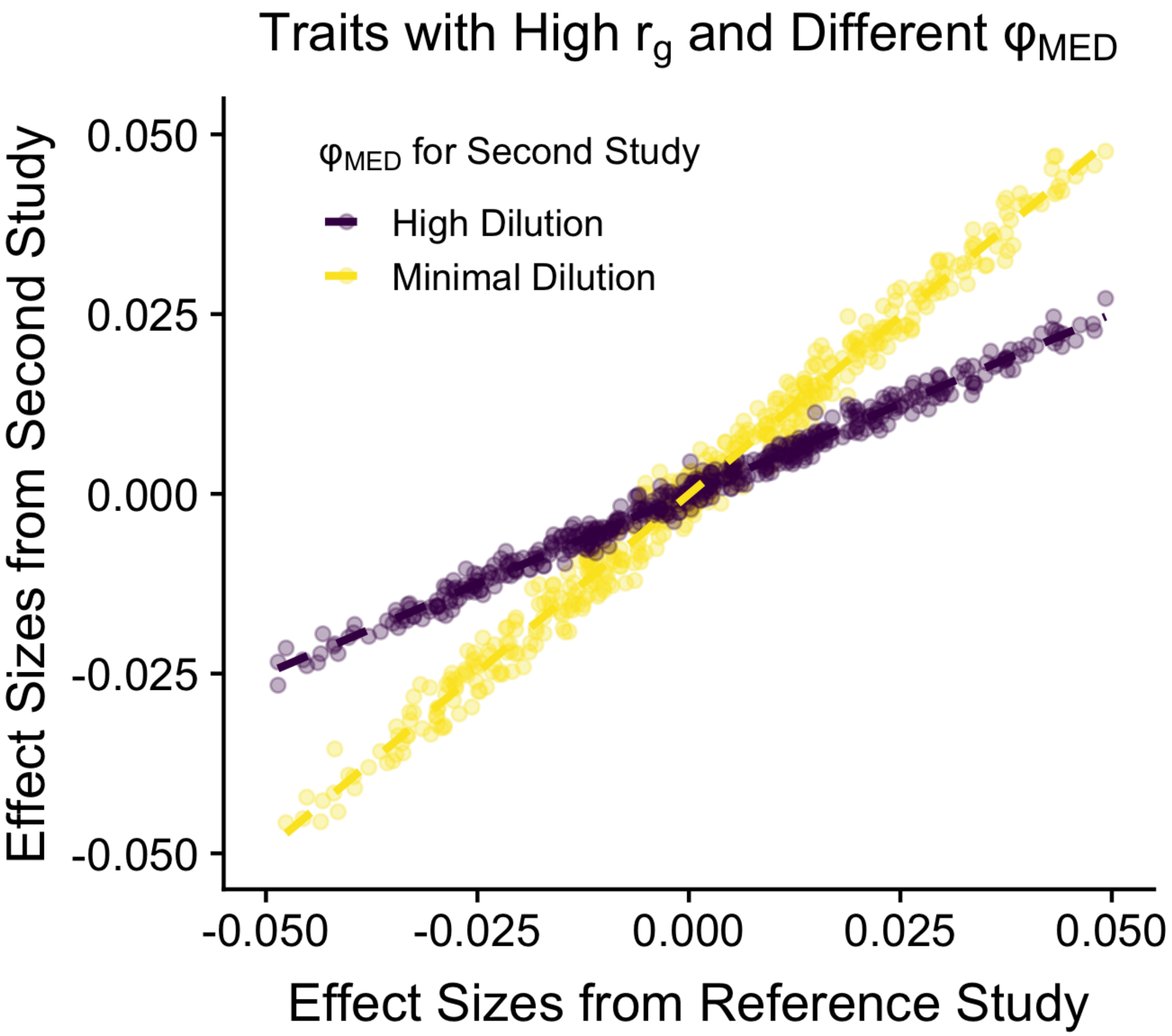


**Figure S1 | Illustrative example where two different GWA studies can have high genetic correlation ($r_g$) with a reference study but different levels of effective dilution ($\varphi_{MED}$).** Note that both studies have high genetic correlation due to the quality of the linear fit in predicting the effect sizes from the second study with the effect sizes of the reference study. Observe that the effect sizes between the reference study and the second study, highlighted in yellow, have comparable effect sizes, hence there is minimal dilution. In contrast, the effect sizes from the purple second study are much smaller than the effect sizes from the reference study, indicating high levels of dilution.

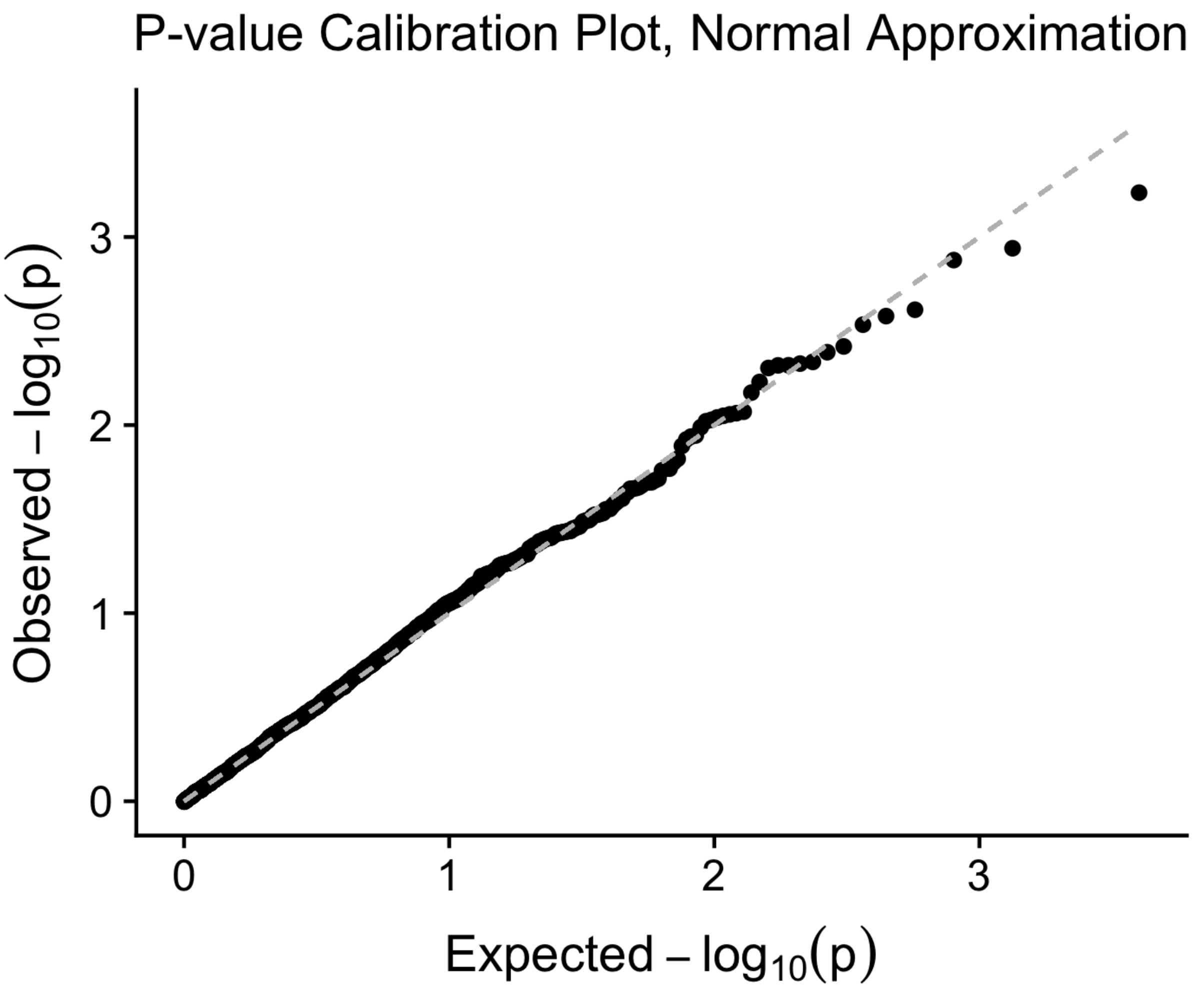


**Figure S2 | P-value calibration plot on the log scale for the p-values outputted by PheMED, using a normal approximation for the effective dilution ($\varphi_{MED}$) test statistics.** The x-axis displays the expected p-values on the log scale, while the y-axis plots the observed p-values on the log scale. Ideally, the observed p-values should follow a uniform distribution and be aligned with the expected p-values, as indicated by the dashed line indicating equality. This simulation suggests that our normal approximation may be conservative towards the tails of the distribution.

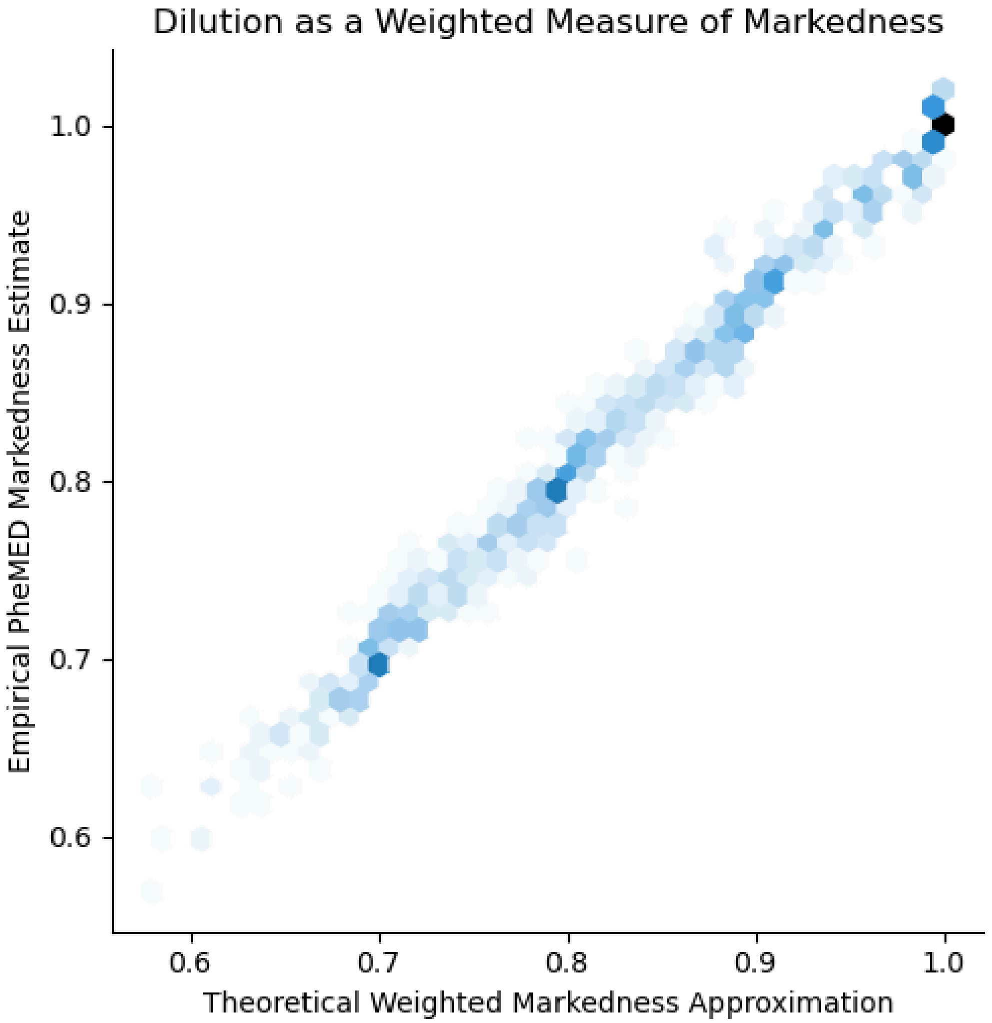


**Figure S3 | Dilution as a weighted measure of markedness.** Here, we provide a two dimensional hexagonal binning density plot from a simulation study to understand what happens when we substitute cases with a genetically correlated, but different trait (e.g. schizophrenia cases with bipolar disorder). The x-axis indicates the theoretical weighted markedness approximation, and the y-axis indicates the empirical PheMED estimate. The color corresponds to the frequency of observations in the corresponding region. The theoretical weighted markedness approximation comes from the derivation in the subsection in the Supplementary Results, the mathematical derivation that phenotype misclassification shrinks the means. The correlation from our theoretical approximation and the empirical estimates is .993. Details on the simulation can be found in the Supplementary Methods section.

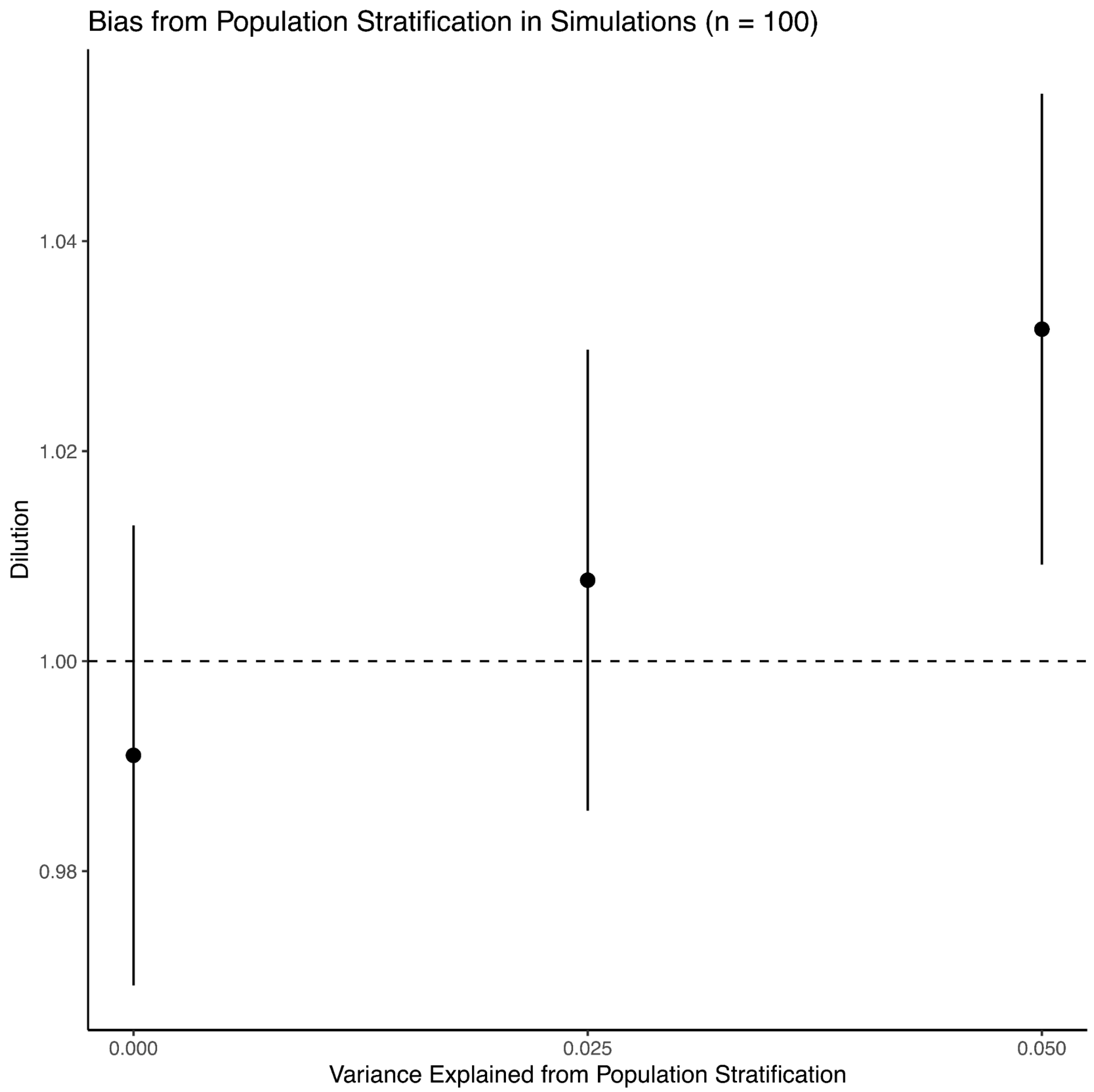


**Figure S4 | Negligible impact of population stratification on dilution.** Here, we provide the results from a simulation study to understand how population stratification can impact the dilution. The x-axis indicates the variance explained from population stratification, where we plot 95% confidence intervals for the dilution on the y-axis. Details on the simulation can be found in the Supplementary Methods section. To assess the potential for population stratification to bias our effective dilution estimates, we simulated a polygenic trait with 25% prevalence and 25% heritability (on the liability scale). We then considered a range of plausible values from 0% to 5% for the percentage of phenotypic variance explained by the 11th principal component (PC), where we measure the dilution between two GWAS's, one that correctly adjusts for all PCs and the other GWAS that does not adjust for the 11th PC. We specifically selected the 11th principal component, as GWAS's often adjust for the first 10 principal components, see for example [7–9]**.**

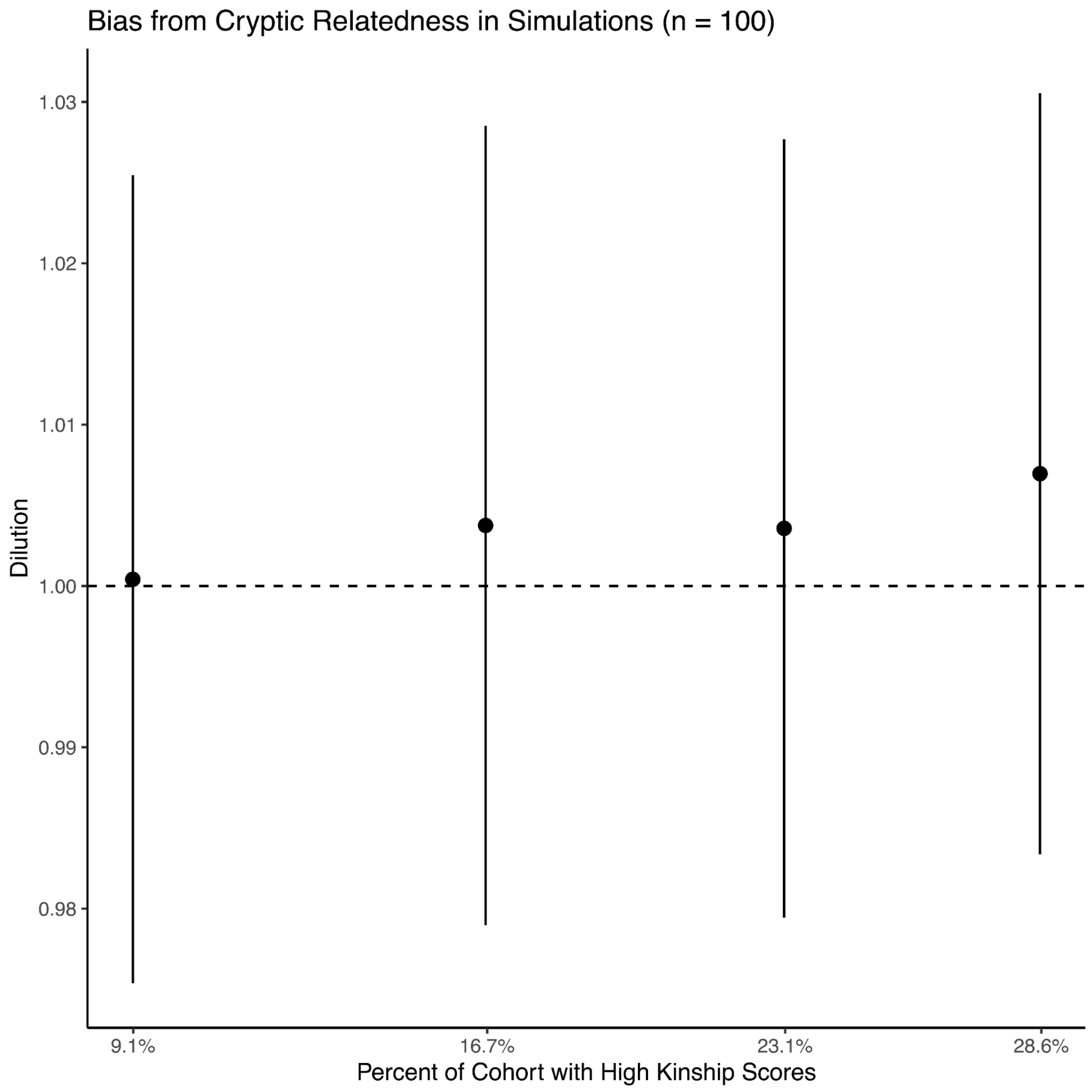


**Figure S5 | Negligible impact of cryptic relatedness on dilution.** Here, we provide the results from a simulation study to understand how cryptic relatedness can impact the dilution. Details on the simulation can be found in the Supplementary Methods section. The x-axis indicates the percentage of the cohort with high kinship scores in the more inclusive analysis, and we plot 95% confidence intervals for the dilution, corresponding to the y-axis. To assess the potential for cryptic relatedness to bias our effective dilution estimates, we simulated a polygenic trait with 25% prevalence and 25% heritability (on the liability scale). We then measured the dilution between two GWAS studies, where one study appropriately filters out patients with high kinship scores, whereas the other cohort retains a percentage of the patients in the analysis. This result is consistent with prior results, as cryptic relatedness does not systematically bias SNP effect sizes across the board [10].

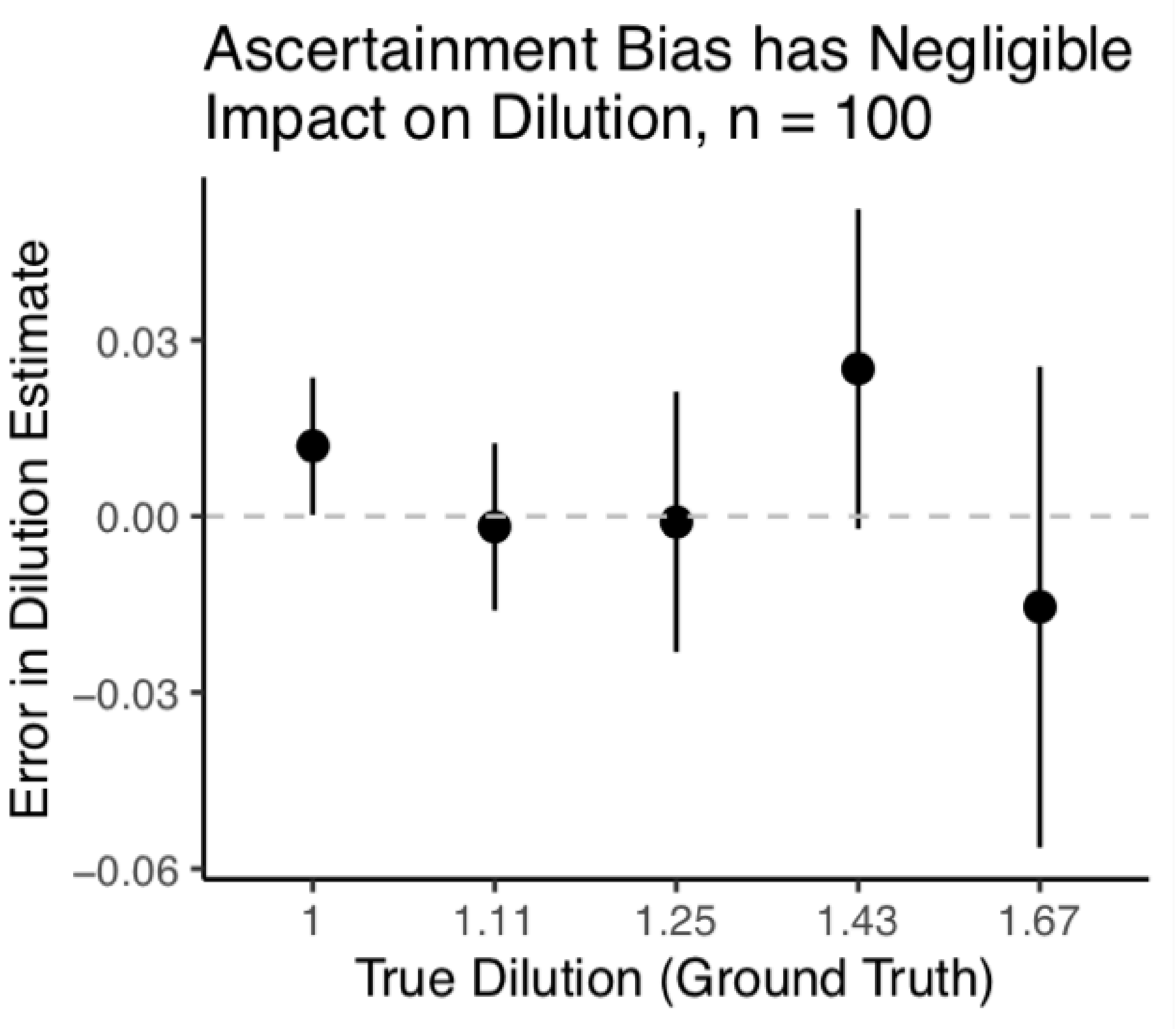


**Figure S6 | Negligible impact of ascertainment bias (sample prevalence) on dilution.** Here, we provide the results from a simulation study to understand how sample prevalence (or ascertainment bias) can impact the dilution. The x-axis indicates the true dilution value. We plot 95% confidence intervals for the error in our dilution estimates corresponding to the y-axis. Details on the simulation can be found in the Supplementary Methods section. To assess the potential for sample prevalence to bias our effective dilution estimates, we simulated a polygenic trait with 25% prevalence, where we measured the dilution between a GWAS that included patients based on the true prevalence, and an ascertained GWAS where the sample prevalence of cases was 50%. We then measured the dilution for different positive predictive values of the labeled phenotype. We note that the wider confidence intervals for higher dilution values are expected due to the log-concavity of the maximum likelihood curve for extreme dilution values.

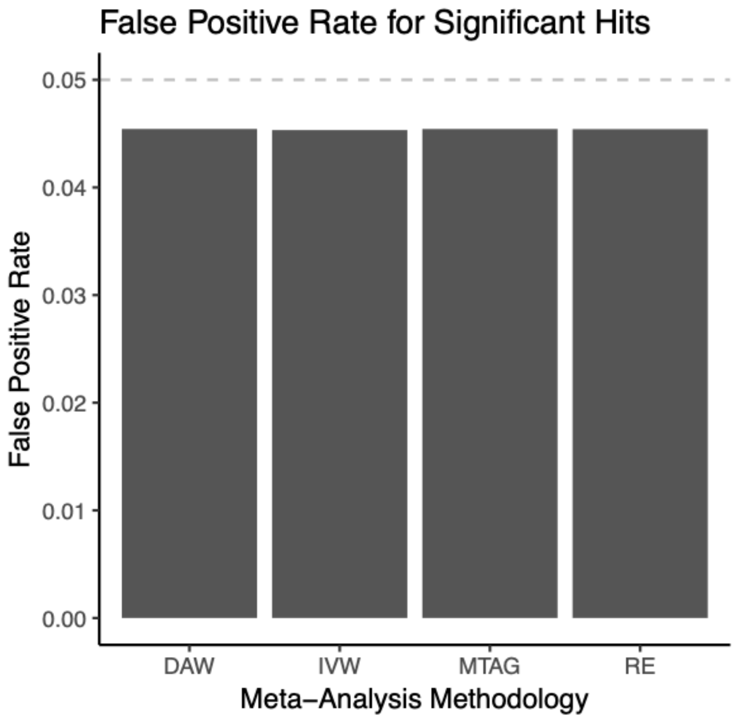


**Figure S7 | Simulation study for the false positive rate for significant hits across meta-analysis methodologies, including Inverse Variance Weights (IVW), MTAG and Random Effects (RE).** The y-axis plots the false positive rate, whereas the x-axis indicates the meta-analysis methodology.

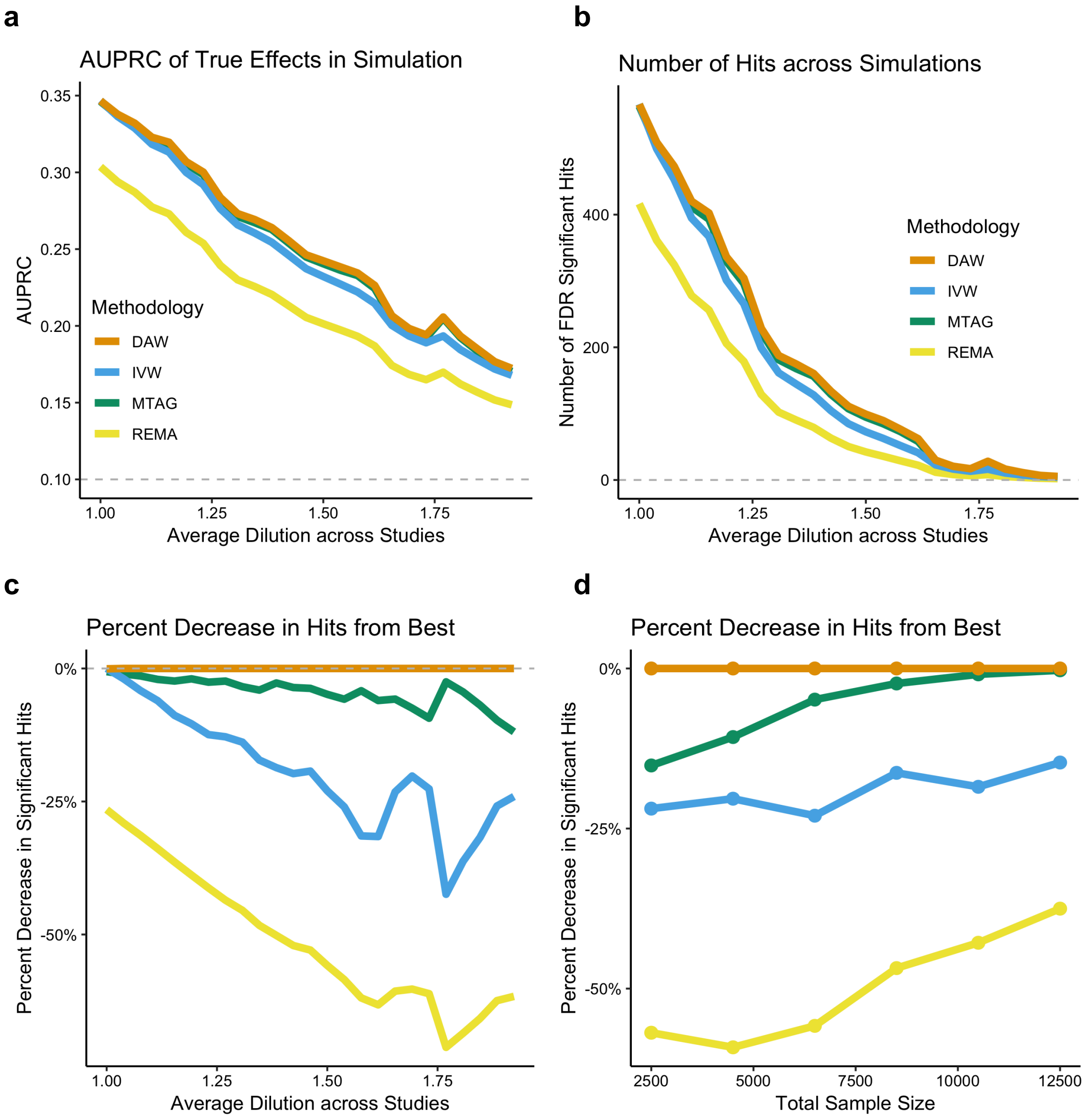


**Figure S8 | Performance of different GWAS meta-analytic approaches.** a: Plotting the area under the precision recall curve for detecting true effects (SNPs), where SNPs are ranked by their p-value. The x-axis denotes the average dilution across studies in the simulation. The dashed line at y = 0.10 denotes performance of a random classifier. Different colors correspond to different methodologies. b: Depicts how the average dilution across studies (x-axis) impacts the number of FDR-significant hits for each methodology (y-axis). c: Depicts how the average dilution across studies (x-axis) impacts the relative percent decrease in FDR significant hits compared to the top performing methodology (y-axis). For panels a-c, the total sample size across studies is fixed at 6,500. d: Illustrates how the total sample size (x-axis) influences the percent decrease in FDR significant hits across different methodologies when the average effective dilution is approximately 1.5.

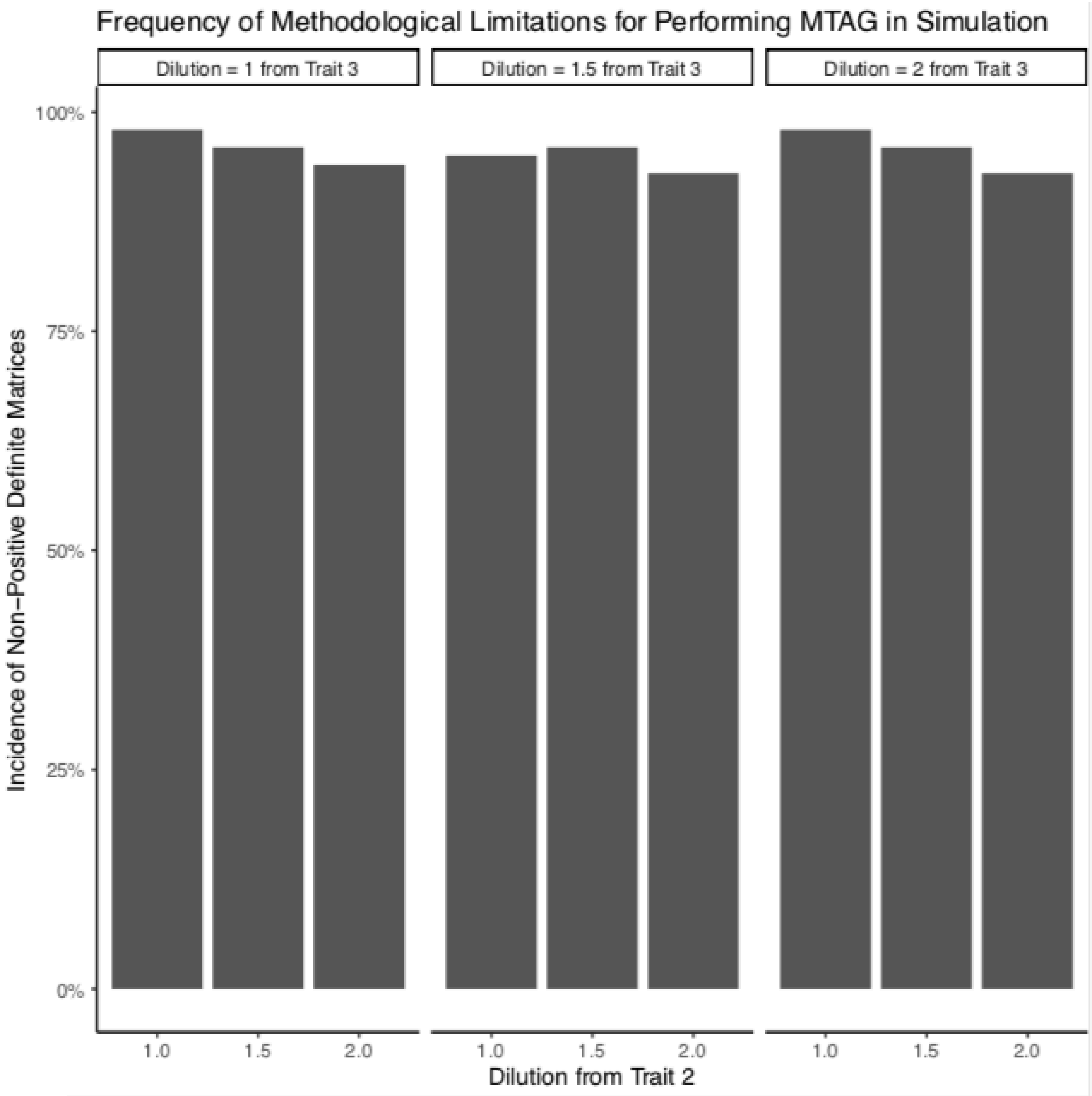


**Figure S9 | Methodological limitations when performing MTAG meta-analyses on GWAS's with modest sample sizes.** Here, we provide the incidence of non-positive definite matrices from a simulation study on real-world genomic data from the Million Veteran Program as outputted by MTAG [11], to compare performance between MTAG and PheMED/Dilution Adjusted Weights Meta-Analysis. Details on the simulation can be found in the Supplementary Methods section. The x-axis indicates the dilution corresponding to the trait from the second GWAS. The y-axis identifies the incidence of non-positive definite matrices, as computed by MTAG. Each cluster in the bar chart corresponds to a different dilution value for the trait corresponding to the third GWAS. (The dilution of the first GWAS, by construction of the simulation, is 1).

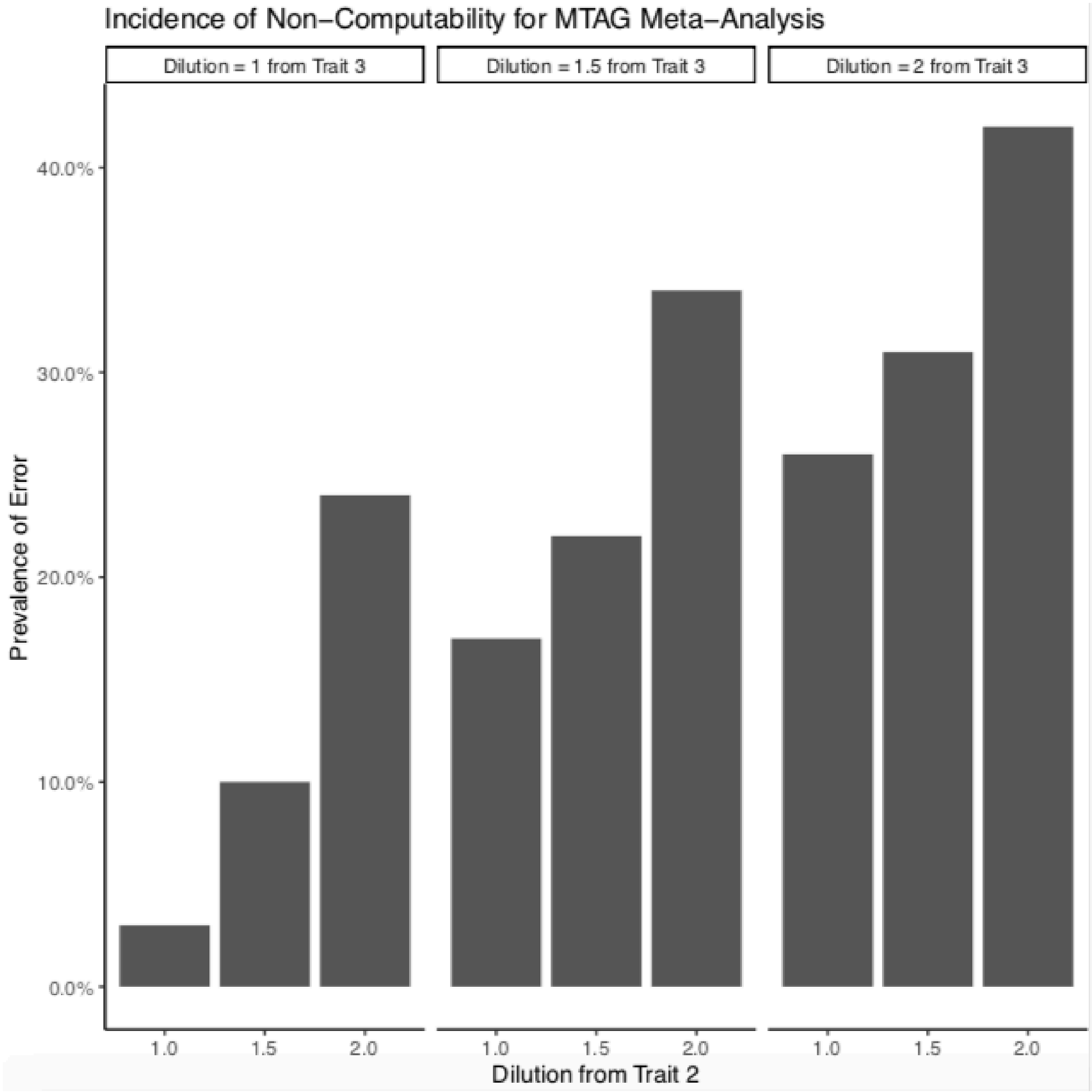


**Figure S10 | Incidence of non-computability for MTAG meta-analyses on GWAS's with modest sample sizes.** Here, we provide the incidence of non-computability for MTAG meta-analyses from a simulation study on real-world genomic data from the Million Veteran Program as outputted by MTAG[11]. The x-axis indicates the dilution corresponding to the trait from the second GWAS. The y-axis identifies the prevalence of noncomputability errors from MTAG. Each cluster in the bar chart corresponds to a different dilution value for the trait corresponding to the third GWAS. Details on the simulation can be found in the Supplementary Methods section. MTAG attempts to perform a matrix adjustment to ensure the covariance matrix is positive definite. However even with this matrix adjustment, MTAG generated missing values in the meta-analysis outputs for up to 42% of the simulations when both traits 2 and 3 were highly diluted (dilution = 2) and over 24% when one of the traits was highly diluted.

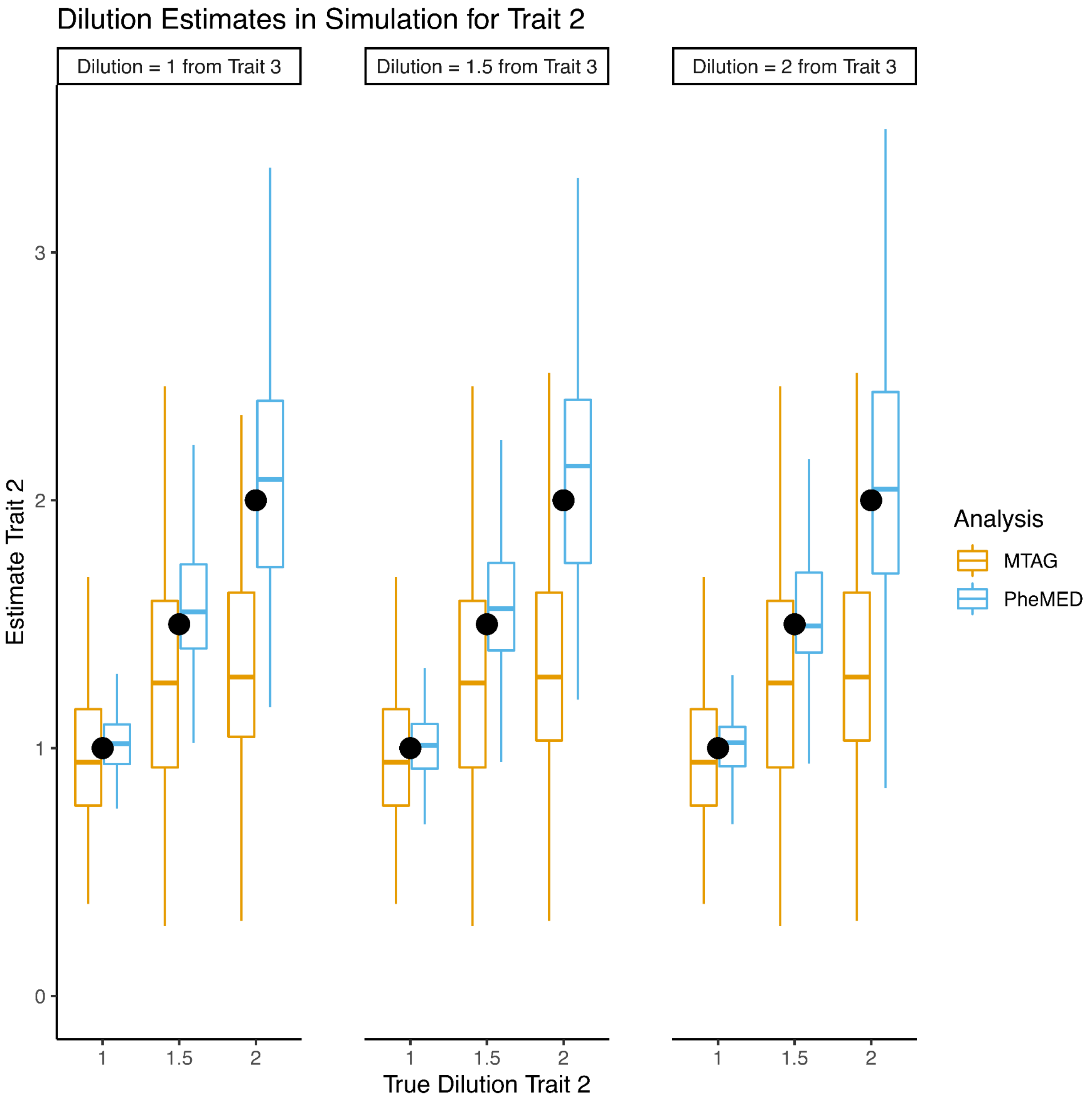


**Figure S11a | Measuring dilution with MTAG and PheMED.** Here, we compare the performance of MTAG and PheMED on measuring the dilution from a simulation study on real-world genomic data from the Million Veteran Program. Details on the simulation can be found in the Supplementary Methods section. The x-axis indicates the dilution corresponding to the trait from the second GWAS. The y-axis identifies the estimated dilution value and the color of the boxplot indicates the methodology used to measure the dilution. Each cluster corresponds to a different dilution value for the trait corresponding to the third GWAS. The black dot highlights the true dilution value. For 10 of the 18 dilution estimates (figures S11a/b), MTAG generated dilution estimates that were significantly more biased than PheMED (fdr significant at the .05 level, 10,000 bootstrap samples). Across the other 8 dilution estimates, PheMED had greater precision than MTAG in estimating the dilution (fdr significant at the .05 level, 10,000 bootstrap samples).

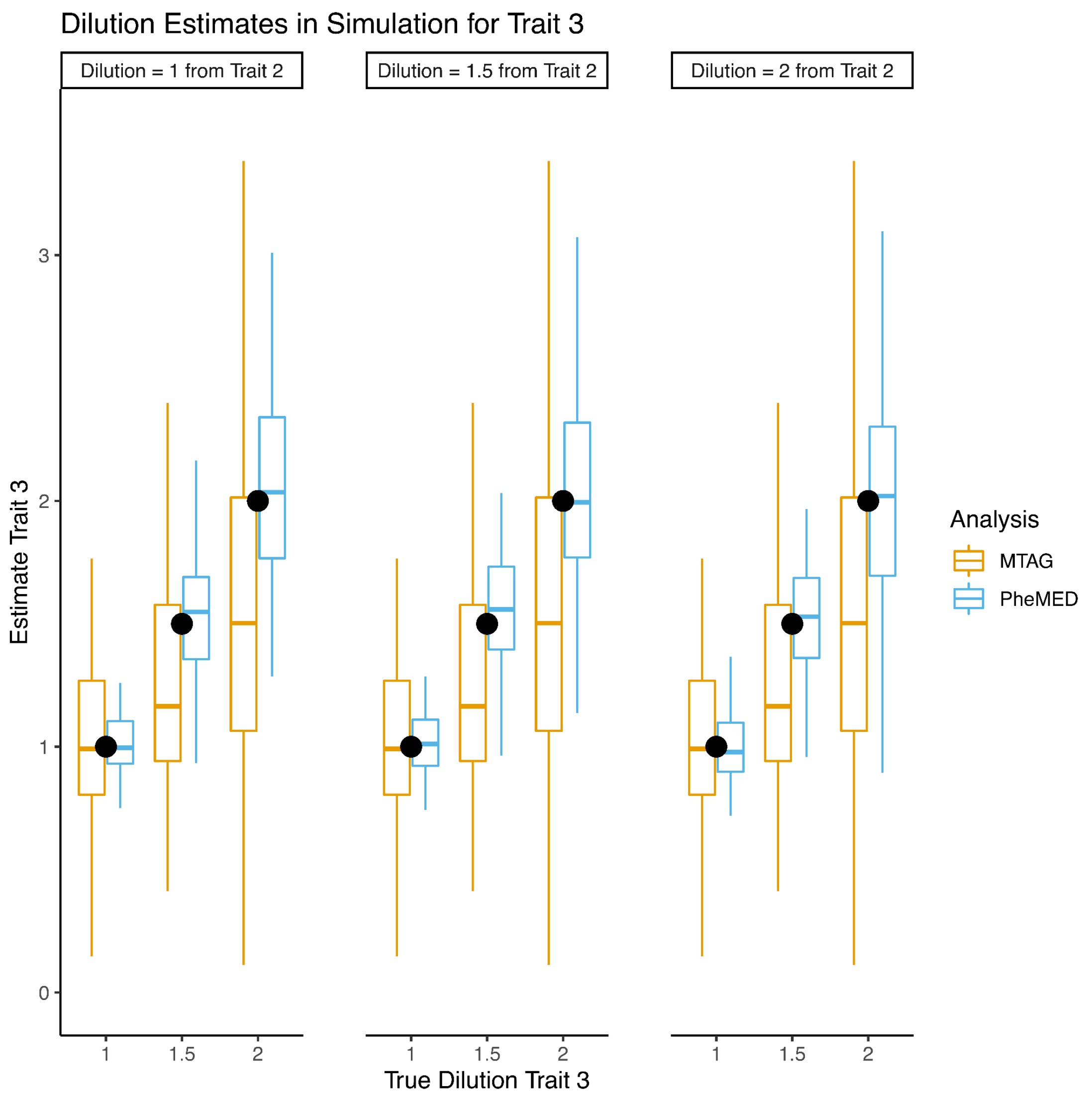


**Figure S11b | Measuring dilution with MTAG and PheMED.** Here, we compare the performance of MTAG and PheMED on measuring the dilution from a simulation study on real-world genomic data from the Million Veteran Program. The x-axis indicates the dilution corresponding to the trait from the third GWAS. The y-axis identifies the estimated dilution value and the color of the boxplot indicates the methodology used to measure the dilution.The black dot highlights the true dilution value. Each cluster corresponds to a different dilution value for the trait corresponding to the third GWAS. Details on the simulation can be found in the Supplementary Methods section. For 10 of the 18 dilution estimates (figures S11a/b), MTAG generated dilution estimates that were significantly more biased than PheMED (fdr significant at the .05 level, 10,000 bootstrap samples). Across the other 8 dilution estimates, PheMED had greater precision than MTAG in estimating the dilution (fdr significant at the .05 level, 10,000 bootstrap samples).

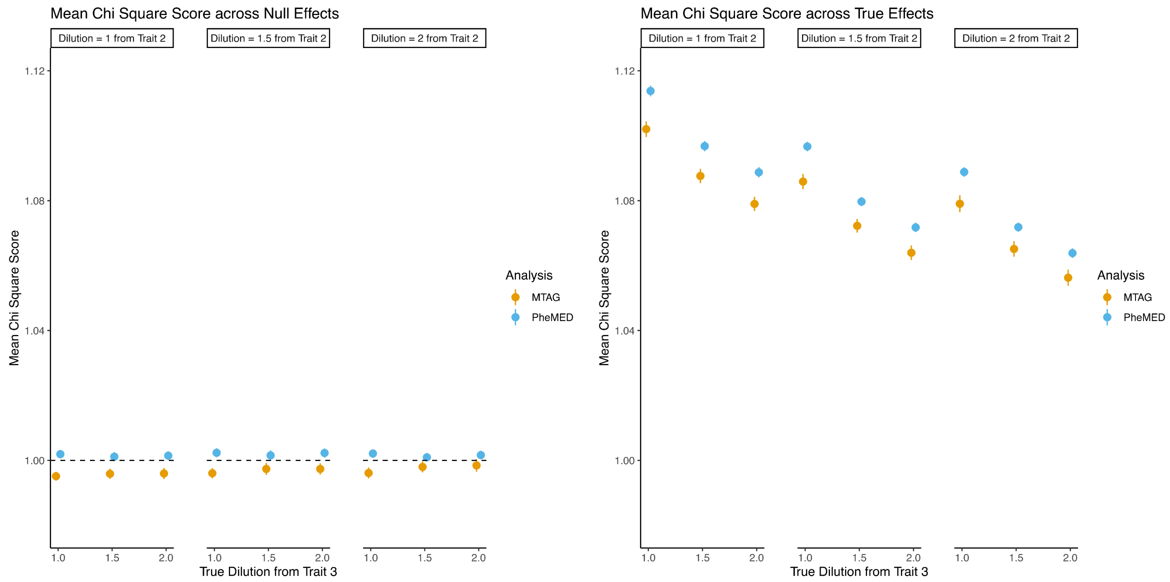


**Figure S12 | Measuring dilution with MTAG and PheMED.** Here, we compare the performance of MTAG and PheMED/Dilution Adjusted Weights Meta-Analysis on measuring the dilution from a simulation study on real-world genomic data from the Million Veteran Program. On the left, we provide 95% confidence intervals for the inflation of the chi square statistic across null effects (y-axis) and on the right we provide 95% confidence intervals for the mean chi square statistic across true effects (y-axis). The x-axis indicates the true dilution value for the trait from the third GWAS. The cluster (subplot) indicates the dilution of the trait from the second GWAS. The color corresponds to the meta-analysis methodology. See the Supplementary Methods section for details.

Number of FDR Significant Hits for Different TWAS Use Cases

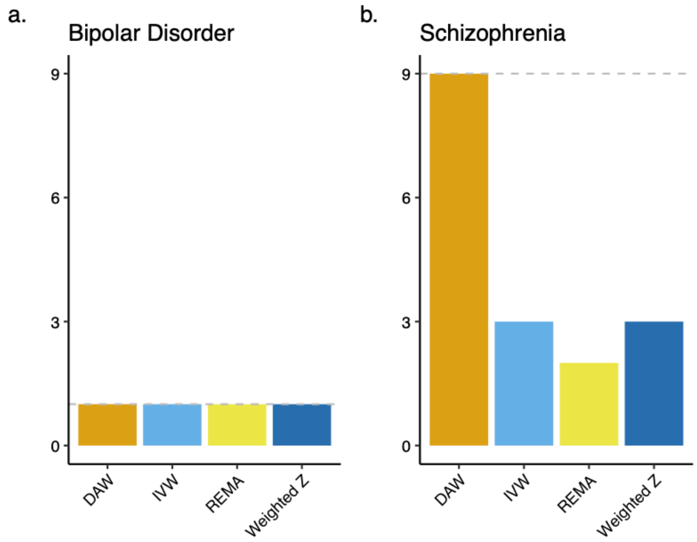


**Figure S13 | Number of FDR Significant Hits (y-axis) found by different meta-analysis methodologies (x-axis) for different TWAS use-cases.** The gray dashed line indicates the maximum number of FDR significant hits across meta-analysis methodologies.

#### Supplementary Results

##### Phenotypic misclassification shrinks our means, an overview

Consider the log odds ratios from two different logistic regression analyses, $\beta_{gold}$, $\beta_{diluted}$, where $\beta_{gold}$ corresponds to the analysis that uses a perfectly encoded phenotype, while $\beta_{diluted}$ corresponds to the diluted phenotype, where some patients may be misclassified. Then we have the following linear approximation[12,13] relating $\beta_{gold}$ and $\beta_{diluted}$:

$$\beta_{diluted} \approx (PPV + NPV - 1)\,\beta_{gold} \qquad (1)$$

where PPV and NPV stand for the positive and negative predictive value of the labels under the diluted phenotype. For a self-contained derivation, we refer the reader to the next section. The dilution factor, PPV + NPV - 1, found in equation (1) is often referred to the 'markedness' or Δp of the possibly imperfectly labeled phenotype[14]. Observe that the multiplicative factor relating $\beta_{gold}$ to $\beta_{diluted}$, only depends on the positive and negative predictive values of the diluted phenotype. We note that the derivation provided by Beesley et al.[13,15] assumes that the effect sizes $\beta_{gold}$ are small and the mislabeling is independent of other confounders. In our work, we can circumvent the use of a gold-standard phenotype by comparing the effective dilution between two possibly diluted phenotypes. For example equation (1) implies that we can relate two different studies 1 and 2, to the same gold-standard study so that

$$\beta_{diluted,1} \approx (PPV_1 + NPV_1 - 1)\,\beta_{gold}$$
$$\beta_{diluted,2} \approx (PPV_2 + NPV_2 - 1)\,\beta_{gold}\,.$$

Therefore, even if we do not have access to a gold-standard study, we can still connect the two diluted phenotypes, as the above relationships imply that

$$\beta_{diluted,2} \approx \frac{(PPV_2 + NPV_2 - 1)}{(PPV_1 + NPV_1 - 1)}\,\beta_{diluted,1}\,. \qquad (2)$$

Let the factor be defined as

$$\varphi_{MED} = \frac{(PPV_1 + NPV_1 - 1)}{(PPV_2 + NPV_2 - 1)}\,,$$

so that

$$\beta_{diluted,2} \approx \beta_{diluted,1}/\varphi_{MED} \qquad (3)$$

#### Mathematical derivation that phenotype misclassification shrinks the means.

Here we provide mathematical derivations illustrating how phenotype misclassification bias dilutes the effect sizes from logs odds ratios. We first consider the impact of phenotype misclassification bias on the difference of means of two groups in Lemma 1. Subsequently in Lemmas 2-3, , we illustrate how changes in the difference of means relates to log odds ratios. And finally in Lemma 4, we extend our results, to consider the impact of phenotype misclassification on the difference of means where there are multiple groups/traits.

First, we introduce some notation. Denote PPV, NPV to be the positive predictive value and negative predictive value of the labeled trait. Furthermore, define $p_{Case}$ to be the true mean probability of observing a SNP across correctly labeled cases and $p_{controls}$ to be the true mean probability of observing a SNP across correctly labeled controls. Then under phenotype misclassification, we are interested in the quantities $p_{Labeled\ Case}$ and $p_{Labeled\ Controls.}$

**Lemma 1:** $p_{Labeled\ Cases} - p_{Labeled\ Controls} = (PPV + NPV - 1) \cdot (p_{cases} - p_{controls})$

**Proof.** From the above definitions we have that,

$$p_{Labeled\ Case} = PPV \cdot p_{Case} + (1 - PPV) \cdot p_{Controls}$$
$$p_{Labeled\ Controls} = NPV \cdot p_{Controls} + (1 - NPV) \cdot p_{Case},$$

where we assume that the probability of observing a SNP is independent of the labeled case-control status when conditioning on the individual's true case-control status. When we take the difference between the two groups, we get that:
$p_{Labeled\ Cases} - p_{Labeled\ Controls} = (PPV - 1 + NPV) \cdot p_{case} + (1 - PPV - NPV) \cdot p_{controls} = (PPV + NPV - 1) \cdot (p_{cases} - p_{controls})$ . □

**Remark 1:** Hence the difference of means from the labeled phenotype gets shrunk by the markedness.

We now connect the difference of means in probability to the log odds ratio with the following result. But first, we introduce additional notation.

Let $p_{MAF}$ be the minor allele frequency

**Lemma 2:** Assume that the log odds ratio is small, or more formally that $\frac{p_{Labeled\ Cases} - p_{Labeled\ Controls}}{p_{MAF}}$ is small. Then the log odds ratio, $\beta \approx \frac{1}{p_{MAF}(1 - p_{MAF})} (p_{Labeled\ Cases} - p_{Labeled\ Controls})$.

**Remark 2:** By combining Lemmas 1 and 2, we get that the log odds ratios also shrink by the markedness as well.

**Proof:** $\beta = log(\frac{p_{Labeled\ Cases}/(1-p_{Labeled\ Cases})}{p_{Labeled\ Controls}/(1-p_{Labeled\ Controls})}) = log(\frac{p_{Labeled\ Cases}}{p_{Labeled\ Controls}}) - log(\frac{1-p_{Labeled\ Cases}}{1-p_{Labeled\ Controls}}) =$
$log(\frac{p_{Labeled\ Controls}}{p_{Labeled\ Controls}} + \frac{p_{Labeled\ Cases}-p_{Labeled\ Controls}}{p_{Labeled\ Controls}}) - log(\frac{1-p_{Labeled\ Controls}}{1-p_{Labeled\ Controls}} - \frac{p_{Labeled\ Cases}-p_{Labeled\ Controls}}{1-p_{Labeled\ Controls}}) =$
$log(1 + \frac{p_{Labeled\ Cases}-p_{Labeled\ Controls}}{p_{Labeled\ Controls}}) - log(1 - \frac{p_{Labeled\ Cases}-p_{Labeled\ Controls}}{1-p_{Labeled\ Controls}}) =$
$log(1 + \frac{p_{Labeled\ Cases}-p_{Labeled\ Controls}}{p_{MAF}+(p_{Labeled\ Controls}-p_{MAF})}) - log(1 - \frac{p_{Labeled\ Cases}-p_{Labeled\ Controls}}{1-p_{MAF}-(p_{Labeled\ Controls}-p_{MAF})}),$
where $p_{MAF}$ is the minor allele frequency. We now assume that the effect sizes (log odds ratios) are small. In particular, we assume that $\frac{p_{Labeled\ Cases}-p_{Labeled\ Controls}}{p_{MAF}}$ (We will later show in Lemma 3 that if $\frac{p_{Labeled\ Cases}-p_{Labeled\ Controls}}{p_{MAF}}$ is small, then both $\frac{p_{Labeled\ Controls}-p_{MAF}}{p_{MAF}}$ and $\frac{p_{Labeled\ Controls}-p_{MAF}}{1-p_{MAF}}$ are small).
When this is the case, we can then take a first order Taylor Series expansion and get the following approximation,

$$\beta \approx \frac{p_{Labeled\ Cases}-p_{Labeled\ Controls}}{p_{MAF}} + \frac{p_{Labeled\ Cases}-p_{Labeled\ Controls}}{1-p_{MAF}} = \frac{1}{p_{MAF}(1-p_{MAF})}(p_{Labeled\ Cases} - p_{Labeled\ Controls})$$

□

**Remark 3:** To justify the assumption in the proof, we now prove the following Lemma

**Lemma 3.** If $\frac{p_{Labeled\ Cases}-p_{Labeled\ Controls}}{p_{MAF}}$ is small, then both $\frac{p_{Labeled\ Controls}-p_{MAF}}{p_{MAF}}$ and $\frac{p_{Labeled\ Controls}-p_{MAF}}{1-p_{MAF}}$ are small as well. More rigorously,

$$\frac{|p_{Labeled\ Controls}-p_{MAF}|}{1-p_{MAF}} \leq \frac{|p_{Labeled\ Controls}-p_{MAF}|}{p_{MAF}} \leq \frac{|p_{Labeled\ Cases}-p_{Labeled\ Controls}|}{p_{MAF}}.$$

**Proof.** Trivially, by definition of the minor allele frequency, the first part of the result is true as

$$\frac{|p_{Labeled\ Controls}-p_{MAF}|}{1-p_{MAF}} \leq \frac{|p_{Labeled\ Controls}-p_{MAF}|}{p_{MAF}}.$$

Finally, if we show that $\frac{|p_{Labeled\ Controls}-p_{MAF}|}{p_{MAF}} \leq \frac{|p_{Labeled\ Cases}-p_{Labeled\ Controls}|}{p_{MAF}}$, then we proved our result.
Equivalently, we need to show that $|p_{Labeled\ Controls} - p_{MAF}| \leq |p_{Labeled\ Cases} - p_{Labeled\ Controls}|$.
By definition of the minor allele frequency, the frequency of the allele is some weighted average between cases and controls, where there is some weight $\alpha \in [0, 1]$, such that
$p_{MAF} = \alpha p_{Labeled\ Controls} + (1 - \alpha)p_{Labeled\ Cases}$.
Hence, $|p_{Labeled\ Controls} - p_{MAF}| = |(1 - \alpha)p_{Labeled\ Controls} - (1 - \alpha)\, p_{Labeled\ Cases}| =$
$(1 - \alpha)\, |p_{Labeled\ Cases} - p_{Labeled\ Controls}| \leq |p_{Labeled\ Cases} - p_{Labeled\ Controls}|$. □

Here we extend our analysis from Lemma 1 to demonstrate what happens to the effect sizes (on average), when misclassification occurs with related traits.

First, we define the following notation. For three traits A, B and C (control), define $p_A$, $p_B$, $p_C$ to be the probability of observing the SNP given the person has the corresponding trait (A, B or C).

Furthermore, define $PPV_A$ to be the positive predictive value of trait A, or the percentage of labeled cases that have trait A. Similarly, define $NPV_A$ to be the percentage of labeled controls that have trait A. We define $PPV_B$, $PPV_C$, $NPV_B$, and $NPV_C$ in a similar fashion.

We will prove the following result:

**Lemma 4.** Suppose that $p_B - p_C \approx z(p_A - p_C)$, that is, we can relate the difference of probabilities between traits B and C, to a linear function of the difference of probabilities between traits A and C. Then

$$p_{Labeled\ Cases} - p_{Labeled\ Controls} \approx (PPV_A + zPPV_B - NPV_A - zNPV_B)(p_A - p_C).$$

**Remark 4:** Observe that when $PPV_B = NPV_B = 0$, this result reduces to our finding in Section A.1, as $NPV_A = 1 - NPV_B - NPV_C = 1 - NPV_C$.

**Remark 5:** From Lemma 2, the assumption that $p_B - p_C \approx z(p_A - p_C)$ will be true when there is high genetic correlation between the log odds ratios from traits A and traits B.

**Proof.** We now have that $p_{Labeled\ Cases} = PPV_A p_A + PPV_B p_B + PPV_C p_C$.

Similarly, $p_{Labeled\ Controls} = NPV_A p_A + NPV_B p_B + NPV_C p_C$.

Assuming the approximation holds, $p_B - p_C \approx z(p_A - p_C)$, then $p_B \approx z(p_A - p_C) + p_C$. Substituting this relationship in the above equations yields:

$$p_{Labeled\ Cases} \approx PPV_A p_A + PPV_B [z(p_A - p_C) + p_C] + PPV_C p_C$$
$$p_{Labeled\ Controls} \approx NPV_A p_A + NPV_B [z(p_A - p_C) + p_C] + NPV_C p_C.$$

Now using the fact that $PPV_B + PPV_C = 1 - PPV_A$, we get that

$$p_{Labeled\ Cases} \approx PPV_A p_A + PPV_B [z(p_A - p_C)] + PPV_B p_C + PPV_C p_C = PPV_A p_A + PPV_B [z(p_A - p_C)] + (1 - PPV_A) p_C$$

Similarly, $p_{Labeled\ Controls} \approx NPV_A p_A + NPV_B [z(p_A - p_C)] + NPV_B p_C + NPV_C p_C = NPV_A p_A + NPV_B [z(p_A - p_C)] + (1 - NPV_A) p_C$.

Now when we take the difference of probabilities between labeled cases and labeled controls, we get that

$p_{Labeled\ Cases} - p_{Labeled\ Controls} \approx PPV_A p_A + PPV_B [z(p_A - p_C)] + (1 - PPV_A) p_C - NPV_A p_A - NPV_B [z(p_A - p_C)] - (1 - NPV_A) p_C = PPV_A p_A + PPV_B [z(p_A - p_C)] - PPV_A p_C - NPV_A p_A - NPV_B [z(p_A - p_C)] + NPV_A p_C = PPV_A (p_A - p_C) + PPV_B z(p_A - p_C) - NPV_A (p_A - p_C) - NPV_B z(p_A - p_C).$

Consequently,
$p_{Labeled\ Cases} - p_{Labeled\ Controls} \approx (PPV_A + zPPV_B - NPV_A - zNPV_B)(p_A - p_C). \square$

#### Supplementary Derivation on Dilution Adjusted Effective Sample Size

For binary phenotypes, we often define the effective sample size as

$$N_{eff} = 4/(\frac{1}{N_{cases}} + \frac{1}{N_{controls}}),$$

although other effective sample size definitions are possible.

Since the effective sample size is proportional to the inverse variance for our β estimates, we can then define a dilution-adjusted effective sample size by considering how effective dilution affects our dilution-corrected estimates for the log-odds ratios. Denote the dilution-adjusted effective sample size as $N_{\varphi_{eff}}$. From the maximum likelihood framework from equation (1) in our main work, we need to multiply our log-odds ratio estimates by the effective dilution, so that estimates from different studies are measured on the same scale, $\varphi_{MED}\beta_{diluted} \sim N(\mu, \varphi^2_{MED}\sigma^2)$. We then have that

$$Var(\varphi_{MED}\beta_{diluted}) \propto \frac{\varphi^2_{MED}}{N_{eff}}$$

By considering the inverse of the variance, we now have an expression for the dilution adjusted effective sample size:

$$N_{\varphi_{eff}} = N_{eff}/\varphi^2_{MED}.$$

Note that this derivation implicitly assumes that all effective dilution values are at least 1. (In the event we have an effective dilution value less than 1, based on equation 3 in our manuscript, we can always choose that study to be our reference study, where our new dilution estimate will just be the reciprocal of our original dilution estimate.) Furthermore, if all of our studies are significantly diluted relative to ground truth, this dilution adjusted sample size correction will be insufficient in estimating the true dilution adjusted effective sample size, as $\varphi_{MED}$ only measures relative effective dilution. Nevertheless, this methodology will still yield accurate ratios of dilution adjusted effective sample sizes between studies, even if no gold standard study is available. (In practice, when comparing sample sizes across studies, we are often interested in the ratio of sample sizes between studies, as opposed to the difference, as this quantity tells us the factor of how much the standard errors will shrink when considering a larger study.)

#### Joint Estimate of Effective Dilution Increases Power

Perhaps surprisingly, we note that the uncertainty in estimating the effective dilution decreases when we jointly estimate the effective dilution across multiple studies, even though the studies are independent results of one another. When analyzing the critical points that optimize the log likelihood, we note that the effective dilution for a given study is a function of the true means scaled according to the reference study. Nevertheless, the true means for a given SNP are a function of all of the studies. Hence, decreasing our uncertainty in estimating the true mean for a given SNP, decreases our uncertainty in estimating $\varphi_k$, the scaling factor to make the means across studies comparable.

Consequently, when there is no sample overlap, we recommend jointly analyzing the effective dilution across multiple studies, as joint estimation of the effective dilution will yield more precise estimates.

#### Derivation for Estimating Power with Dilution

To estimate the power when we perform a one-sided Z test at 5% significance level, we can utilize the expression,

$1 - \Phi(1.64 - \beta/SE)$,

where $\Phi$ represents the cumulative distribution function of the normal distribution.
We wish to extend our power analysis to account for effective dilution, resulting in the following expression,

$1 - \Phi(1.64 - \beta/(\varphi_{MED} \times SE))$.

**Proof:** Suppose our effect size estimate for our SNP comes from a normal distribution, $X \sim N(\beta/\varphi_{MED}, SE)$, with specified effect size and standard error and suppose without loss of generality the true effect size is non-negative and we are performing a one-sided hypothesis test. Then under the null-hypothesis, the probability we reject the null is,

$Pr(X/SE > z_\alpha)$ where $z_\alpha$ represents the critical z-score to achieve a statistically significant result. Now define $Z = (X - \beta/\varphi_{MED})/SE \sim N(0, 1)$. Consequently, we have that

$Pr(X/SE > z_\alpha) = Pr(Z > z_\alpha - (\beta/\varphi_{MED}SE)) = 1 - Pr(Z < z_\alpha - \beta/(\varphi_{MED}SE)) =$

$1 - \Phi(z_\alpha - \beta/(\varphi_{MED}SE))$.

Substituting the critical value for reaching the 5% significance level will then yield the desired expression. We readily note that this methodology for adjusting our power computations for dilution driven effect-size heterogeneity across studies also extends to the more general case of transformed effects meta-analysis (as described later in this Supplement), such as when we wish to validate a SNP with a GWAS defined using continuous phenotype that was discovered using a GWAS with a related binary phenotype or vice versa.

#### Derivation Illustrating how Dilution Affects Performance Metrics in Polygenic Risk Score Validation

Here, we provide a mathematical derivation illustrating how dilution can affect performance metrics from polygenic risk score (PRS) validation. Denote $PPV_{PRS,Observed}$, $NPV_{PRS,Observed}$ as the observed positive predictive value and negative predictive value from a polygenic risk score model on a possibly diluted cohort and denote $PPV_{Pheno}$, $NPV_{Pheno}$ as the positive and negative predictive value of the labeled phenotype. We then have the following linear system of equations relating the true positive and negative predictive values from the positive risk score model under no dilution:

$$PPV_{PRS,Observed} = PPV_{PRS,Optimal} \times PPV_{Pheno} + (1 - NPV_{PRS,Optimal}) \times (1 - PPV_{Pheno})$$
$$NPV_{PRS,Observed} = NPV_{PRS,Optimal} \times NPV_{Pheno} + (1 - PPV_{PRS,Optimal}) \times (1 - NPV_{Pheno}),$$

where we can view our observed PPV and NPV from the PRS model as a weighted average of the optimal PPV and NPV on a validation cohort without phenotypic misclassification. Given the PPV and NPV of the observed PRS model and the labeled phenotype, we can then solve the system of linear equations to infer what the optimal PPV and NPV values are in a non-diluted validation cohort.

In practice, $\varphi_{MED}$ only provides information on the ratio of markedness values, $PPV_{Pheno} + NPV_{Pheno} - 1$. If desired, we can then perform a more thorough sensitivity analysis, where we consider all possible (or plausible) values for $PPV_{Pheno}$, $NPV_{Pheno}$ with a given markedness. We also have the option of further simplifying our analysis, if we assume that our best study is reasonably close to a gold-standard study.

#### Overview of Transformed Effects Meta-Analysis (TEMA), an Extension of Dilution Weights Meta-Analysis

When performing transformed effects meta-analysis, we posit that all studies share a common mean but the true study mean is transformed based on a hidden function specific to each study, which we infer from the data. We note that dilution adjusted weights (DAW) meta-analysis is a special case of TEMA, where the unknown transformation corresponds to the unknown amount of dilution of the phenotype. Mathematically, for approximately independent SNPs, our log-likelihood has the form:

$$l(D|\vec{\alpha}, \vec{\mu}, \vec{\sigma}) = \Pi_{i,j} \frac{1}{\sqrt{2\pi}\sigma_{ij}} exp(- \frac{(\beta_{ij} - f(\mu_i, \vec{\alpha}_j))^2}{2\sigma_{ij}^{\ 2}}),$$

where i denotes the SNP and j denotes the study.

We require our function $f(\mu, \vec{\alpha}): \mathbb{R}^{n+1} \rightarrow \mathbb{R}$ satisfy certain properties.

a. $f(\mu, \vec{1}) = \mu$, a value for $\vec{\alpha}$ to indicate that no transformation is required. Without loss of generality, we use $\vec{1}$, but note other values are possible.
b. $f(0, \vec{\alpha}) = 0$ that null effects are preserved regardless of transformation.
c. $\frac{\partial f}{\partial \mu}(\mu, \vec{\alpha}) > 0$, for all $\mu$, $\vec{\alpha}$. That is, if the true effect size increases, the corresponding transformed effect must increase in size as well for any choice of parameter $\vec{\alpha}$.

When performing transformed effects meta-analysis, we must first specify a family of functions for $f(\mu, \vec{\alpha})$. The focus of this paper considers the simplest case, where $\alpha = 1/\varphi_{MED}$ and

$f(\mu, \alpha): \mathbb{R}^2 \rightarrow \mathbb{R}$, $f(\mu, \alpha) = \alpha\mu$. We readily observe that the family of functions specified by $f(\mu, \alpha) = \alpha\mu$, satisfies properties (a)-(c) provided $\alpha > 0$.

Of course, we can choose more complicated non-linear examples such as multivariate polynomials of the form:

$f(\mu, \alpha_1, \alpha_2) = \alpha_1\mu + (1 - \alpha_1)^3\mu + (1 - \alpha_2)^2\mu + (1 - \alpha_1)(1 - \alpha_2)\mu$, where $0 < \alpha_1 \leq 1$ and $0 \leq \alpha_2 \leq 1$.

After specifying the family of functions for our transformed effects meta-analysis, (e.g. $f(\mu, \alpha) = \alpha\mu$), we estimate the unknown parameter $\alpha$ using maximum likelihood. In the special linear case, once we determine $\alpha$, we can perform a modified inverse variance weighted meta-analysis, described in the methods. Alternatively, we can perform a likelihood ratio test to compute p-values under the null hypothesis that the true-effect is null.

#### Comparison of DAW with other meta-analysis methods

In this section, we provide a mathematical overview illustrating why competing meta-analysis methodologies require invalid assumptions.

**For inverse variance weight (IVW) meta-analysis**, we maximize the log-likelihood function,

$$l(\mu,\ \beta,\ \sigma)\ =\ \sum_i log\ N(\beta_i|\mu,\ \sigma_i^2).$$

From this formulation, it becomes readily apparent that all estimated effect sizes across studies share the same mean $\mu$. Of course, when dealing with diluted phenotypes corresponding to studies of varying levels of quality, this assumption is difficult to defend. We now explain why this methodology is called “inverse variance weights”.

Computationally, we can derive closed-form expressions for the mean $\mu$ that maximizes the log-likelihood function. In particular, by leveraging the first-derivative test, we find that

$$0\ = \frac{\partial l(\mu, \beta, \sigma)}{\partial \mu} =\ \sum_i \frac{\beta_i - \mu}{\sigma_i^2}\ \Leftrightarrow$$

$$\mu\ (\sum_i 1/\sigma_i^2)\ \ = \sum_i \beta_i/(\sigma_i^2)\ ,$$

where $\mu$ is a weighted average of the estimated effect sizes across studies and the weights correspond to the inverse-variance of each study.

**For weighted Z meta-analysis,** we weight the Z-scores by the square root of effective sample sizes,

$$\sum_i \sqrt{N_{eff,i}}\, Z_i.$$

Since weighted Z uses effective sample sizes and not dilution adjusted effective sample sizes, we are likely to lose power when conducting a weighted Z meta-analysis. Furthermore, in the special case that $\sigma_i\ =\ 1\sqrt{N_{eff,i}}$, weighted Z meta-analysis is actually an IVW meta-analysis in disguise, as

$$\sum_i \sqrt{N_{eff,i}}\, Z_i\ =\ \sum_i \sqrt{N_{eff,i}} \frac{\beta_i}{\sigma_i}\ =\ \sum_i \frac{\beta_i}{\sigma_i^2}\ .$$

**For random effects meta-analysis,** we maximize log-likelihood function

$$l(\mu,\ \beta,\ \sigma,\ \tau)\ =\ \sum_i log\ N(\beta_i|\mu,\ \sigma_i^2\ +\ \tau^2).$$

Random effects meta-analysis allows for heterogeneity across studies by adding a parameter $\tau^2$ to our variance. Foremost, this formulation is prone to lose power as the degrees of freedom for estimating $\tau^2$ depends on the number of studies, which is often very small. In contrast, the degrees of freedom when estimating $\varphi_{MED}$ is very large as it is based on the number of SNPs being tested in the entire GWAS. In addition, the random effects meta-analysis indiscriminately applies the same shrinkage across all studies. In contrast, in a dilution adjusted weights (DAW) meta-analysis framework, we should only change the uncertainty of our estimates from studies that are marked as having poor data quality. Utilizing this methodology is also difficult to defend as in many cases, we know a priori which studies are prone to have lower data quality (e.g. health care disparities, self-reported data, etc.) Furthermore, DAW achieves high power by estimating a single heterogeneity parameter across all SNPs as opposed to random effects, which generates a separate measure of heterogeneity for every SNP.

**For MTAG:**

We also consider the performance of our proposed solution DAW against a multi-trait meta-analysis methodology, MTAG, since different studies for the same trait can be considered as different traits on account of measurement error. When leveraging a multi-trait meta-analytic framework, like MTAG, increasing the number of studies in the meta-analysis does not impact the relative weights of each study, because transitive inference is impossible when each study corresponds to a separate trait. In contrast, DAW assumes all studies address the same trait and can achieve increasingly precise weights for estimating the dilution with more studies in the meta-analysis. Furthermore, DAW affords more flexibility by enabling the utilization of summary statistics with low heritability and/or sample sizes because it is not built on top of LDSC. Consequently, we demonstrate both on real-world data and through simulation that DAW meta-analysis consistently achieves higher power for detecting true effects compared to all of the aforementioned methodologies, including IVW and MTAG without increasing false positive rate in simulations. Therefore, we maintain that the DAW meta-analysis framework is a much needed step towards making our analyses more robust against the aforementioned implicit biases that compromise data quality across studies.

#### Supplementary Methods

##### Multi-trait dilution simulation

To investigate the impact of multi-trait misclassification on the effective dilution, we implemented the following simulation. In this simulation, we have three groups: Group 1 (people with trait A), Group 2 (people with trait B) and Group 3 (controls). For simplicity, to distinguish between trait A and trait B, we have a hidden variable so that no patient can have both trait A and B. We establish one study as a reference with no misclassification, where the objective is to perform a GWAS on trait A. We define the other study with a prescribed positive predictive value along with a trait B mislabeling percentage, that indicates what percent of the misclassified cases have trait B. We do the same for negative predictive value, but for simplicity, we assume all misclassified controls have trait B.

We use the following parameter values, where:
PPV can range from .7, .8, .9, 1.
NPV can range from .9, .95, 1.
The percentage of incorrectly labeled cases with trait B can range from 0, .1, .2, .3, .4, .5.
The genetic correlation for trait B ranges from .1, .3, .5, .7, .9.
The heritability on the liability scale for trait B ranges from .0625, .125, .25.
The prevalence for trait B and trait A is fixed at 25%.
The heritability on the liability scale for trait A is fixed at 25%.

Note that the values selected for genetic correlation, heritability and NPV are based on biologically plausible values inferred from studies in the literature. (NPV is generally higher than PPV due to case-control imbalance).

**Simulating SNP level data.**
We simulated 1,000 potentially causal independent SNPs for traits A and B, across 75,000 individuals. (Only an expected 50% of those potentially causal SNPs could actually be causal for a given trait). The minor allele frequency for SNPs came from a uniform distribution ranging from 5% to 50%. SNPs were then normalized to have unit variance.

In addition, we simulated an additional 50,000 independent null effect SNPs where standard errors were matched with the potentially causal simulated SNPs. However for computational efficiency, null effects were simulated as part of the summary level data. We then constructed the two GWAS study populations from the pool of 75,000 simulated individuals, where the first 50,000 individuals can have either trait A or trait C (control) and the last 25,000 individuals can have either trait B or trait C (control).

Based on the mathematical analysis in the Supplementary Results section, we used a linear approximation for relating the trait A effect sizes to the trait B effect sizes, where we set the slope of the linear approximation to be the genetic correlation.

##### Impact of ascertainment bias/sampling prevalence on dilution, simulation

We simulated SNP data in a similar fashion to our multi-trait dilution simulation for 75,000 patients, see the above section for details. From the 75,000 simulated individuals, the prevalence of the phenotype was 25%. We allocated the first 25,000 individuals to the first GWAS study (where the sample prevalence is consistent with the population prevalence). We then constructed the second GWAS from the remaining 50,000 individuals, randomly filtering out controls, such that the number of cases in the sample matched the number of controls. We also considered the impact of dilution on the ascertained study, where the positive predictive value of the labeled phenotype could range from [.6,.7,.8,.9,1].

##### Measuring impact of population stratification on dilution with schizophrenia GWAS's

We measure the dilution when comparing two Schizophrenia GWAS's from the Million Veteran Program (MVP), where we randomly allocate cases and controls to a different group and perform GWAS's on the different groups. For one of the groups, we adjust for a fixed number of PC's (20). For the other group, we vary the number of PC's that we adjust (n = 5, 10, 15, 20). We leveraged release 4 of the MVP, which contains genotyping data from blood samples for more than 650,000 individuals. For these simulations, we leveraged imputed genotypes from TOPMed [16]. For QC methods for the genotyping data, we refer the reader to the earlier section in the Methods, *Genotyping, quality control and imputation.*

##### Measuring impact of population stratification on simulated traits with real-world genomic data

To assess the potential for population stratification to bias our effective dilution estimates, we simulated a polygenic trait with 25% prevalence and 25% heritability (on the liability scale). We selected 5% of the SNPs to be causal SNPs. Construction of these simulated polygenic traits for 40,000 randomly selected European ancestry individuals followed the approach taken by [17] . We adjusted the percentage of phenotypic variance explained by the 11th principal component. We specifically selected the 11th principal component, as GWAS's often adjust for the first 10 principal components, see for example [7–9]**.** Finally, after splitting the cohort into two groups, we compared the effective dilution between one analysis that correctly adjusts for all 11 principal components with another analysis that only corrects for 10 principal components.

##### Measuring impact of cryptic relatedness on simulated traits with real-world genomic data

We repeat the same analysis as described above, except here, in addition to the 40,000 individuals, we potentially retain an additional 4,000 pairs (8,000 individuals total) with kinship scores exceeding .088. We measure the dilution between one group where the kinship scores are all below .088, and another group where we retain n related pairs of individuals, where the n pairs range from 1,000 to 4,000.

#### Assessing performance of MTAG and PheMED/dilution adjusted weights meta-analysis on real-world genomic data

In this simulation, we meta-analyze three traits. Based on our findings, we focus on the impact of population stratification, as population stratification yielded greater (but still very modest) bias on the PheMED estimates than cryptic relatedness. In this set up, Trait 1 has no dilution and is adjusted for population stratification. Trait 2 may have dilution (1, 1.5, 2) but is adjusted for population stratification. And finally Trait 3, may have dilution(1, 1.5, 2) but is not adjusted for population stratification. We simulate population stratification by setting the variance explained of the 11th principal component to be 2.5%. The heritability of the trait is set to 25% (under no misclassification bias) on the liability scale, the prevalence is set to 25% and each analysis. Each GWAS contains approximately 16,000 patients. We estimate the dilution from MTAG by taking the ratio of the omega values, based on equations (3-5) in the supplement of their paper [11]. To compute p values for the dilution, we constructed 10,000 bootstrap samples from the simulation, and performed a one-sided test to determine whether the dilution from PheMED was closer to ground truth than MTAG. Similarly, we also performed a one-sided test to determine if the interquartile range from PheMED for the dilution was smaller than MTAG. When constructing the polygenic phenotype, only 10% of the SNPs from chromosomes 1-10 were causal and the rest of the SNPs were null. The heritability of the polygenic trait on the liability scale was set to 25% and the prevalence was also set to 25%. For details on constructing the polygenic trait, we refer the reader to the prior section in the supplementary methods section, *Measuring impact of population stratification on simulated traits with real-world genomic data.*

Bibliography


1. Lam, M. *et al.* Comparative genetic architectures of schizophrenia in East Asian and European populations. *Nat. Genet.* **51**, 1670–1678 (2019).
2. Pardiñas, A. F. *et al.* Common schizophrenia alleles are enriched in mutation-intolerant genes and in regions under strong background selection. *Nat. Genet.* **50**, 381–389 (2018).
3. Bigdeli, T. B. *et al.* Genome-Wide Association Studies of Schizophrenia and Bipolar Disorder in a Diverse Cohort of US Veterans. *Schizophr. Bull.* **47**, 517–529 (2021).
4. Duncan, L. *et al.* Significant Locus and Metabolic Genetic Correlations Revealed in Genome-Wide Association Study of Anorexia Nervosa. *Am. J. Psychiatry* **174**, 850–858 (2017).
5. Watson, H. J. *et al.* Genome-wide association study identifies eight risk loci and implicates metabo-psychiatric origins for anorexia nervosa. *Nat. Genet.* **51**, 1207–1214 (2019).
6. Mullins, N. *et al.* Genome-wide association study of more than 40,000 bipolar disorder cases provides new insights into the underlying biology. *Nat. Genet.* **53**, 817–829 (2021).
7. Levey, D. F. *et al.* Bi-ancestral depression GWAS in the Million Veteran Program and meta-analysis in >1.2 million individuals highlight new therapeutic directions. *Nat. Neurosci.* **24**, 954–963 (2021).
8. Demontis, D. *et al.* Genome-wide analyses of ADHD identify 27 risk loci, refine the genetic architecture and implicate several cognitive domains. *Nat. Genet.* **55**, 198–208 (2023).
9. Kurki, M. I. *et al.* FinnGen provides genetic insights from a well-phenotyped isolated population. *Nature* **613**, 508–518 (2023).
10. Gross, A., Tönjes, A. & Scholz, M. On the impact of relatedness on SNP association analysis. *BMC Genet.* **18**, 104 (2017).
11. Turley, P. *et al.* Multi-trait analysis of genome-wide association summary statistics using MTAG. *Nat. Genet.* **50**, 229–237 (2018).
12. Duffy, S. W. *et al.* A simple model for potential use with a misclassified binary outcome in

epidemiology. *J. Epidemiol. Community Health* **58**, 712–717 (2004).

13. Beesley, L. J., Fritsche, L. G. & Mukherjee, B. An analytic framework for exploring sampling and observation process biases in genome and phenome-wide association studies using electronic health records. *Stat. Med.* **39**, 1965–1979 (2020).
14. Powers, D. Evaluation: From precision, recall and fmeasure to roc, informedness, markedness and correlation. (2007).
15. Beesley, L. J. & Mukherjee, B. Statistical inference for association studies using electronic health records: handling both selection bias and outcome misclassification. *Biometrics* **78**, 214–226 (2022).
16. Taliun, D. *et al.* Sequencing of 53,831 diverse genomes from the NHLBI TOPMed Program. *BioRxiv* (2019) doi:10.1101/563866.
17. Hou, K. *et al.* Accurate estimation of SNP-heritability from biobank-scale data irrespective of genetic architecture. *Nat. Genet.* **51**, 1244–1251 (2019).
